# Safety, Immunogenicity and Efficacy of NVX-CoV2373 in Adolescents in PREVENT-19: A Randomized, Phase 3 Trial

**DOI:** 10.1101/2022.09.20.22279903

**Authors:** Germán Áñez, Lisa M. Dunkle, Cynthia L. Gay, Karen L. Kotloff, Jeffrey M. Adelglass, Brandon Essink, James D. Campbell, Shane Cloney-Clark, Mingzhu Zhu, Joyce S. Plested, Pavitra Roychoudhury, Alexander L. Greninger, Nita Patel, Alice McGarry, Wayne Woo, Iksung Cho, Gregory M. Glenn, Filip Dubovsky, the 2019nCoV-301 – Pediatric Expansion Study Group

## Abstract

**BACKGROUND:** Over 20% of cases and 0.4% of deaths from Covid-19 occur in children. Following demonstration of safety and efficacy of the adjuvanted, recombinant spike protein vaccine NVX-CoV2373 in adults, the PREVENT-19 trial enrolled adolescents.

**METHODS:** Safety, immunogenicity, and efficacy of NVX-CoV2373 were evaluated in adolescents aged 12 to <18 years in an expansion of PREVENT-19, a phase 3, randomized, observer-blinded, placebo-controlled trial in the United States. Participants were randomized 2:1 to two doses of NVX-CoV2373 or placebo 21 days apart, and followed for a median of 2 months after second vaccination. Primary end points were serologic non-inferiority of neutralizing antibody (NA) responses compared with young adults (18 to <26 years) in PREVENT-19, protective efficacy against laboratory-confirmed Covid-19, and assessment of reactogenicity/safety.

**RESULTS:** Among 2,247 participants randomized between April–June 2021, 1,491 were allocated to NVX-CoV2373 and 756 to placebo. Post-vaccination, the ratio of NA geometric mean titers in adolescents compared to young adults was 1.5 (95% confidence interval [CI] 1.3 to 1.7). Twenty Covid-19 cases (all mild) occurred: 6 among NVX-CoV2373 and 14 among placebo recipients (vaccine efficacy [VE]: 79.5%, 95% CI, 46.8 to 92.1). All sequenced viral genomes (11/20) were identified as Delta variant (Delta variant VE: 82.0% [95% CI: 32.4 to 95.2]). Reactogenicity was largely mild-to-moderate, transient, and more frequent in NVX-CoV2373 recipients and after the second dose. Serious adverse events were rare and evenly distributed between treatments.

**CONCLUSIONS:** NVX-CoV2373 was safe, immunogenic, and efficacious in the prevention of Covid-19 and those cases caused by the Delta variant in adolescents.

(Funded by the Office of the Assistant Secretary for Preparedness and Response, Biomedical Advanced Research and Development Authority and the National Institute of Allergy and Infectious Diseases (NIAID), National Institutes of Health; PREVENT-19 ClinicalTrials.gov number, NCT04611802).

---

Optimal control of coronavirus disease (Covid-19) as it moves into an endemic status requires that vaccination be extended to all ages to minimize disease overall and, especially, to reduce the social and mental health impact on children and adolescents.^1^ Severe acute respiratory syndrome coronavirus 2 (SARS-CoV-2) spike (S) protein-based vaccines using messenger RNA (mRNA) technology are authorized or approved for use in adolescents and younger children in the United States and elsewhere.^2^ NVX-CoV2373 (Novavax), a recombinant S (rS) protein vaccine co-formulated with a saponin-based adjuvant (Matrix-M™), is also authorized for emergency use in adults ≥18 years of age and recently for adolescents 12–17 years of age in the United States and numerous other countries and regions,^3-11^ based on safety, immunogenicity and protective efficacy against symptomatic Covid-19.^12-14^

We describe herein the safety, immunogenicity, and efficacy results in adolescents 12 through 17 years of age in the pediatric expansion of the PREVENT-19 trial in the United States conducted during 2021, during a period of predominance of the SARS-CoV-2 Delta variant.

## Methods

### TRIAL DESIGN, PARTICIPANTS, PROCEDURES, AND OVERSIGHT

PREVENT-19 is a phase 3, randomized, observer-blinded, placebo-controlled trial conducted in adults in the United States and Mexico evaluating safety, immunogenicity, and efficacy of NVX-CoV2373.^13^ After the primary objective for adults was achieved,^13^ this pediatric expansion enrolled adolescents at 73 clinical sites in the United States from April to June 2021. Healthy adolescents 12 through 17 years of age or those with stable chronic medical conditions, including chronic pulmonary, renal, or cardiovascular disease, diabetes mellitus, or well-controlled human immunodeficiency virus (HIV) infection, were eligible for participation. Key exclusion criteria included known previous laboratory-confirmed SARS-CoV-2 infection or known immunosuppression. Additional details regarding trial design, conduct, oversight, and analyses are provided in the Supplementary Appendix, the protocol, and statistical analysis plan.

Parents or guardians of participants provided written informed consent while participants provided assent before enrollment and randomization, without age stratification, using a web-based interactive system. Participants were allocated in a 2:1 ratio to receive two 0.5-mL intramuscular injections of either NVX-CoV2373 (5 *μg* rS SARS-CoV-2 + 50 *μg* Matrix-M™ adjuvant) or placebo 21 days apart. Site personnel who managed study vaccine logistics/preparation had no subsequent role in participant assessment.

Novavax was the trial sponsor and responsible for study design, development, and manufacture of clinical trial material. Trial data were available to all authors, who vouched for its accuracy and completeness and for fidelity to the trial protocol. The trial is ongoing, and investigators, Novavax clinical team, and the participants, remain blinded to participant-level treatment assignments. The protocol, amendments and overall oversight were approved by institutional review boards/ethics committees according to local regulations. Safety and efficacy were monitored through the placebo-controlled portion of the trial with regular reviews of unblinded data by the NIAID/NIH-sponsored DSMB.^15^

### SAFETY ASSESSMENTS

Solicited local and systemic adverse events (AEs) were collected via electronic diary for 7 days following each injection. Participants were assessed for all unsolicited AEs from the first dose through 28 days after the second dose (Day 49); serious AEs (SAEs), AEs of special interest (AESIs), and medically attended AEs (MAAEs) related to vaccination were collected from the first dose until the end of the study.

### IMMUNOGENICITY ASSESSMENTS

Neutralizing antibodies (NA) specific to SARS-CoV-2 were measured using a validated microneutralization assay (MN) with an inhibitory concentration of 50% (MN50), using wild-type virus strain SARS-CoV-2 hCoV-19/Australia/VIC01/2020 (GenBank MT007544.1) (360biolabs, Australia),^16^ with a lower limit of quantitation (LLOQ) of 20. Additional immunogenicity endpoints included serum anti-SARS-CoV-2 S protein IgG antibody levels,^17^ and human angiotensin-converting enzyme 2 (hACE2) receptor binding inhibition (RBI) antibodies to SARS-CoV-2 S protein.^18^ Both validated enzyme-linked immunosorbent assays (anti-S IgG and hACE2) were conducted at Novavax Clinical Immunology, Gaithersburg, MD, using reagents based on the prototype Wuhan strain. Additionally, in *post hoc* analyses, anti-S IgG and hACE2 RBI antibodies were measured against viral variants using reagents based on the SARS-CoV-2 Alpha, Beta, Delta, Gamma, Mu, and Omicron variants (fit-for-purpose assays conducted at Novavax Vaccine Immunology Laboratory, Gaithersburg, MD). Details of these assays and results are provided in the Supplementary Appendix.

Prior exposure to SARS-CoV-2 was determined by the presence of serum anti-nucleoprotein (NP) antibodies (University of Washington, Seattle, Washington, US), using the Roche Elecsys® Anti-SARS-CoV-2 assay,^19^ and/or a positive SARS-CoV-2 Reverse Transcriptase Polymerase Chain Reaction (RT-PCR) in nasal swab collected at baseline (University of Washington, Seattle, WA, US) using the Abbott RealTime Quantitative SARS-CoV-2 Assay.^4^

### EFFICACY ASSESSMENTS

The efficacy of NVX-CoV2373 in preventing the first episode of RT-PCR-confirmed symptomatic mild, moderate, or severe Covid-19 (according to FDA criteria,^20^ Table S3) with onset ≥7 days after the second injection in the per-protocol population of efficacy (PP-EFF), was summarized descriptively. Symptoms of suspected Covid-19 (Table S2) were reported by participants’ parents or guardians as soon as possible after onset or during weekly calls. When prespecified symptoms were reported, participants were instructed to undergo in-clinic medical evaluation, which included collection of nasal swabs for RT-PCR. End point Covid-19 cases were confirmed by positive nasal swab RT-PCR at the central laboratory. Whole-genome sequencing (WGS) and clade/lineage assignment were performed on RT-PCR-positive samples with sufficient viral RNA load (Supplementary Appendix). Severity of Covid-19 protocol-defined end points was assessed by investigators and study physicians according to protocol-specified criteria, and severe cases were confirmed through review by the external independent End Point Review Committee blinded to treatment assignment.

### STATISTICAL ANALYSIS

#### Safety analysis

Safety data from all participants who received at least one dose of study treatment were summarized descriptively. Severity and duration of solicited local and systemic AEs, reported daily for seven days by participants’ guardians in electronic diaries, were assessed (according to FDA criteria for severity^21^) after each injection. Unsolicited AEs were coded by preferred term and system organ class using the Medical Dictionary for Regulatory Activities (MedDRA, version 24.0), and summarized by severity and investigator-assessed relationship to study vaccine.

#### Immunogenicity analysis

The per-protocol immunogenicity set (PP-IMM) included participants without prior exposure to SARS-CoV-2 who had at least a baseline and one serum sample result available after full primary vaccination series and no major protocol violations considered likely to impact immune response at the corresponding study visit (e.g., RT-PCR-positive swabs or SARS-CoV-2 seropositivity prior to the visit in question). For the primary assessment of effectiveness, i.e., non-inferiority (NI) of the NA response at Day 35 compared with young adults, a formal non-randomized analysis was performed using NA responses from a subset of adolescent participants in the PP-IMM group compared with that observed in a subset of similarly unexposed adult participants 18 to < 26 years of age in the PREVENT-19 study.^4^

For all assays, the geometric mean titer (GMT) at each study visit, and the geometric mean fold rise (GMFR) compared to the baseline (Day 0) at each post-vaccination study visit, with 95% confidence interval (CI) were summarized by treatment group. The 95% CI was calculated based on the t-distribution of the log-transformed values for geometric means or GMFR, then back transformed to the original scale for presentation. Serologic response (SCR) was defined as a proportion of participants with a ≥ 4-fold rise between Days 0 and 35. The 95% CI was calculated using the exact Clopper-Pearson method.

The primary non-inferiority effectiveness objective required meeting three criteria: 1) lower bound (LB) of two-sided 95% CI for the ratio of GMTs (GMT_12-<18yo_/GMT_18-<26yo_) >0.67, 2) point estimate of the ratio of GMTs ≥ 0.82 (estimated as square root of 2/3), and 3) LB of the two-sided 95% CI for difference of SCR (SCR_12-<18yo_ /SCR_18-<26yo_) > -10%.

#### Efficacy analysis

The PP-EFF used for all protective efficacy analyses included participants who 1) had no evidence of prior SARS-CoV-2 infection at baseline or up to ≥7 days after the second injection, 2) received both injections of assigned treatment, and 3) had no major protocol deviations. Vaccine efficacy (VE%) was defined as (1 – RR) × 100, where RR is the relative risk of incidence rates between the two treatment groups. The estimated RR and two-sided 95% CI were derived using Poisson regression with robust error variance.

As implemented earlier for adult participants in PREVENT-19,^13^ a blinded crossover (participants originally randomized to placebo were offered NVX-CoV2373 and vice versa) was implemented for adolescent participants after a median 2-month safety follow-up had been attained. The intention was to offer all participants active vaccine as soon as possible without compromising FDA-required placebo-controlled safety follow-up.

## Results

### PARTICIPANTS

A total of 2,304 participants were screened and 2,247 were randomized between April 26 and June 05, 2021 (Figure 1). The safety analysis set (SafAS) included 2,232 participants who received at least one dose of NVX-CoV2373 (1,487) or placebo (745). 1,799 participants (80% of all randomized) were included in the PP-EFF population and 1,654 (74%) in the PP-IMM population based on evidence of previous SARS-CoV-2 infection (15.7% vs. 16.8% in active vs. placebo groups, respectively) and/or other protocol-defined exclusionary criteria (Table 1).

**Table 1.**
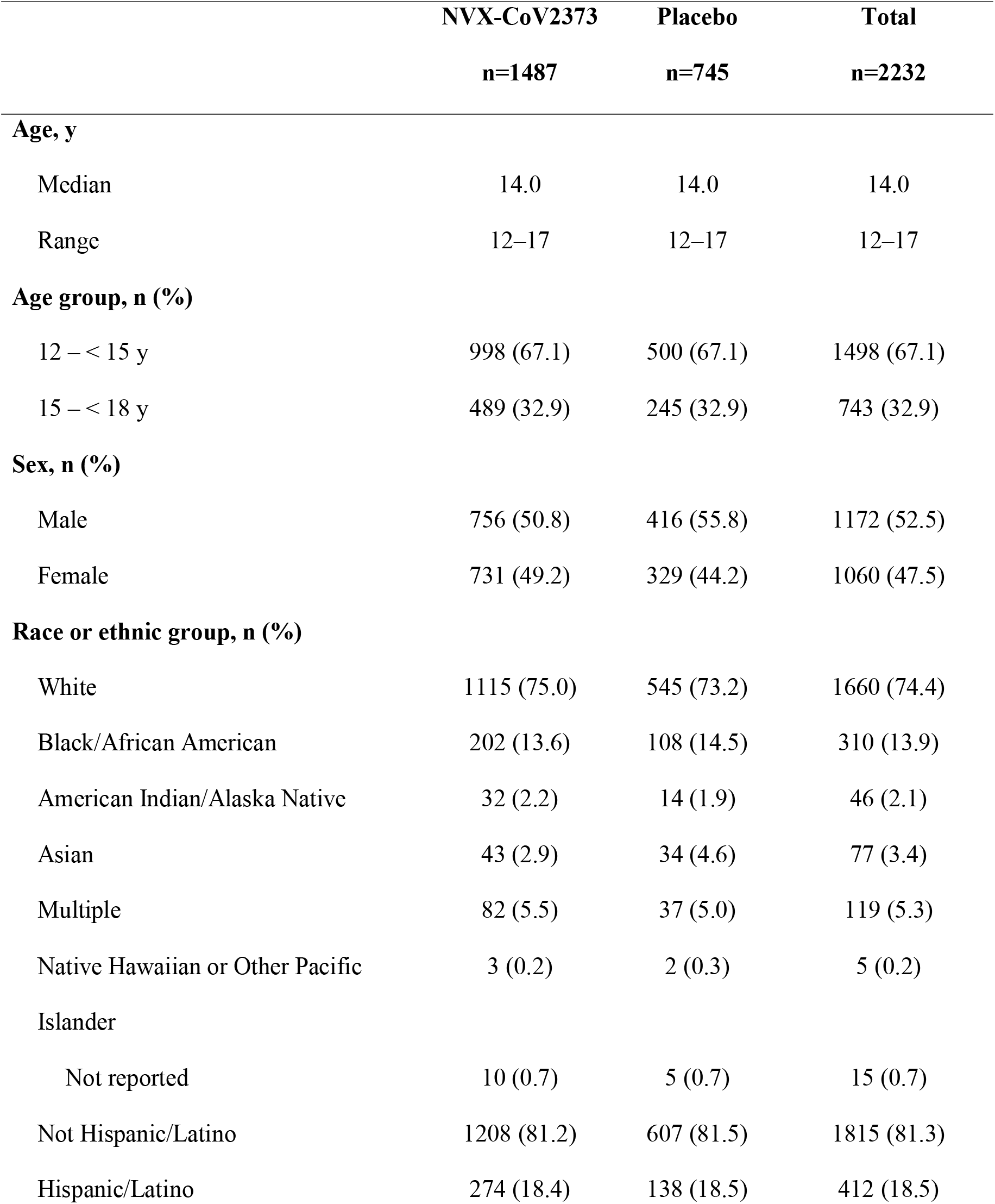
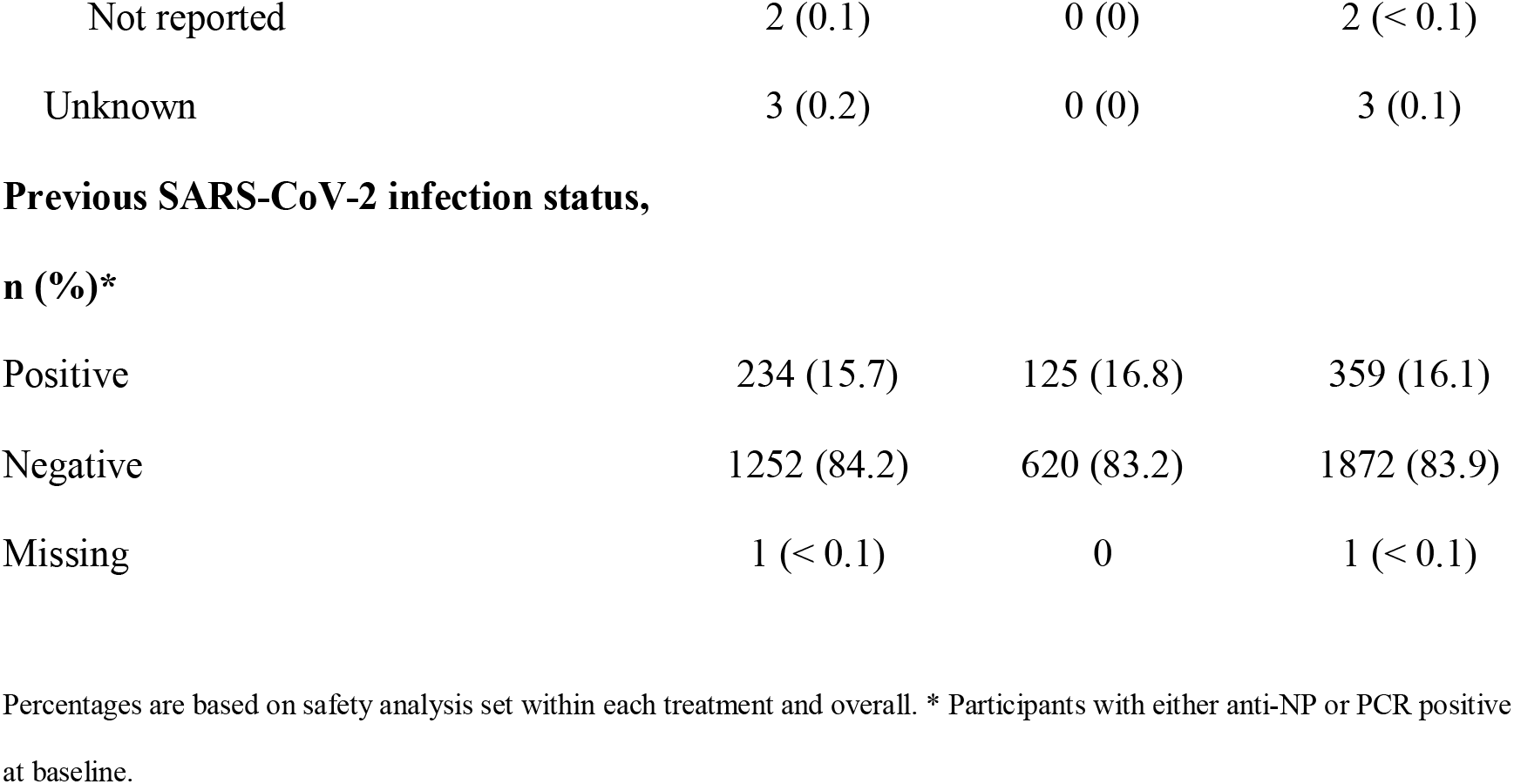
Demographics and Baseline Characteristics (Safety Analysis Set Population).

**Figure 1.**
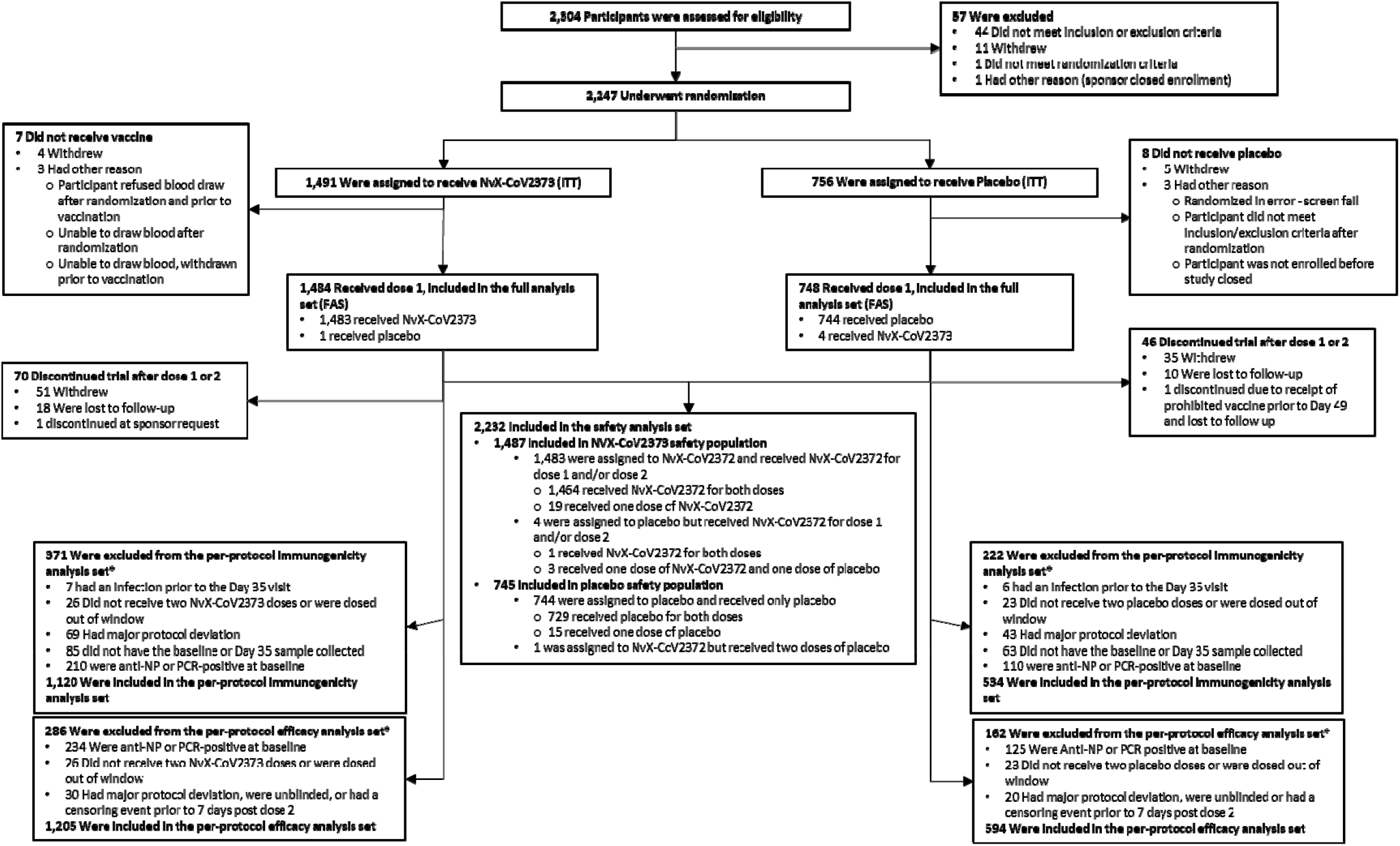
Trial Disposition. The full analysis set (FAS) included all participants who were randomly assigned to treatment and received at least one dose, regardless of protocol violations or missing data, and are analyzed according to the trial vaccine group as randomized. *Participants could have more than one reason for exclusion.

Median duration of safety follow-up after second vaccination was 71 days. Baseline demographics of the SafAS were well balanced between treatment groups; 47.5% self-identified as female, 74.4% as White, 13.9% as Black/African American, 2.1% as American Indian/Alaska Native, and 18.5% as Hispanic/Latino. The median age was 14 years; 67% were 12 to <15 years (Table 1).

### SAFETY

#### Reactogenicity

Solicited local and systemic AEs were predominantly mild-to-moderate in severity and self-limiting, although more frequent in NVX-CoV2373 recipients, and more common after the second injection. After each dose, the most frequently reported solicited local AEs were injection site pain and tenderness. The median duration of these events was ≤2 days (range 1–7 days) (Table S9). Severe (≥Grade 3) local reactions were 1.5% vs. <1% after dose 1, and 8.5% vs. <1% after dose 2 in the NVX-CoV2373 and placebo groups, respectively (Figure 2 and Table S8).

**Figure 2.**
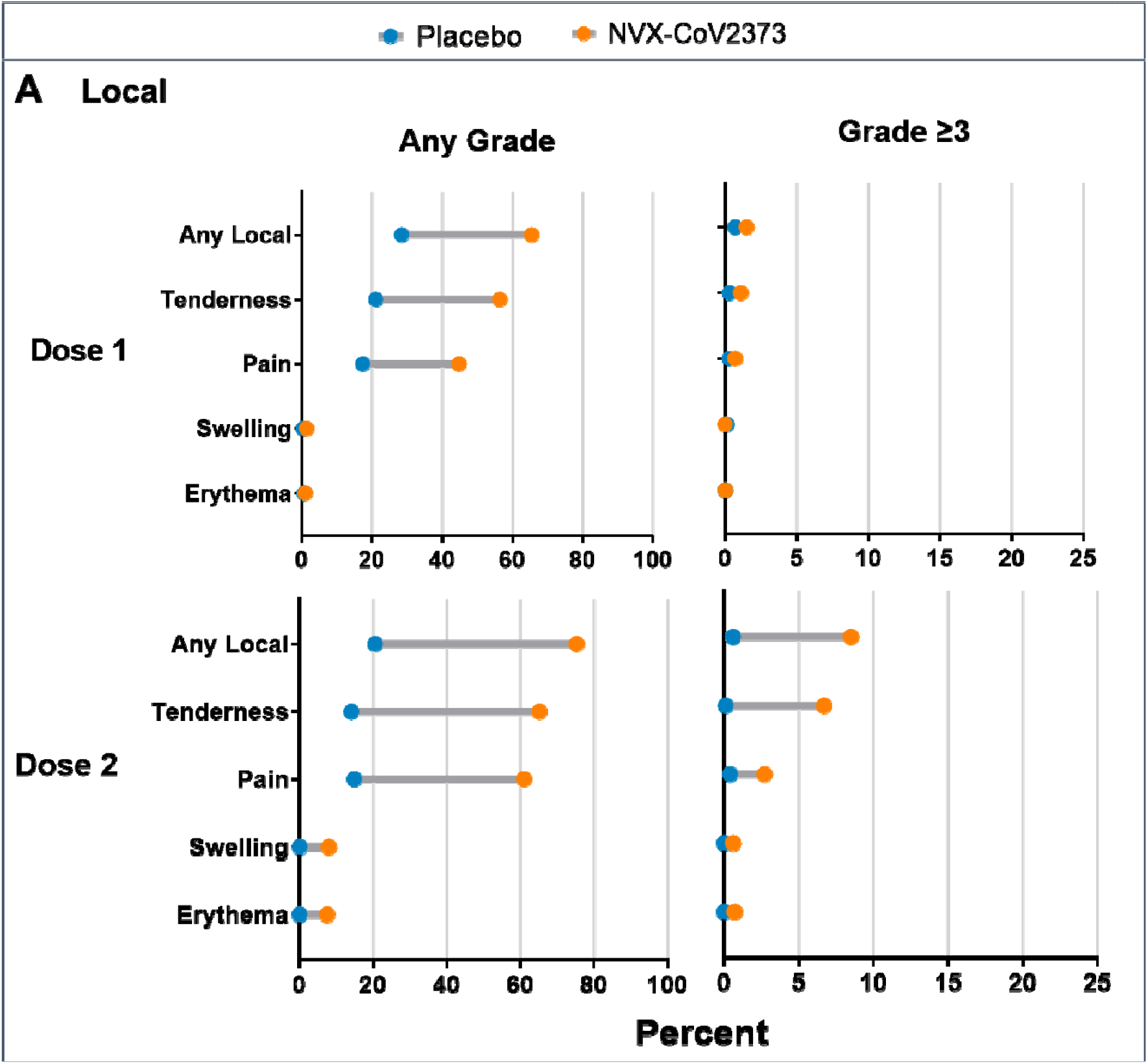
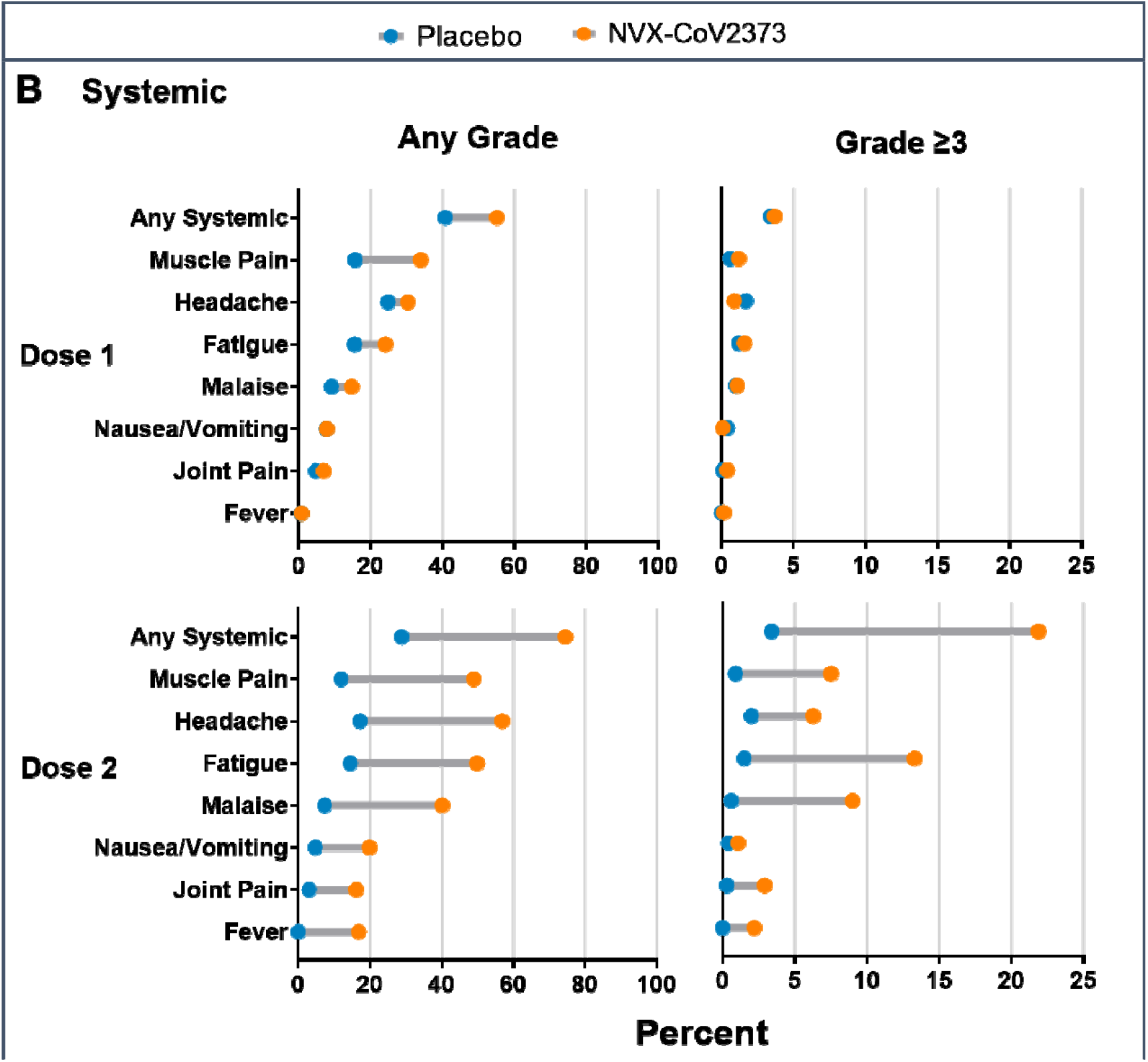
Solicited Local and Systemic Adverse Events. The percentage of participants in each treatment group with solicited local (A) and systemic (B) adverse events during the 7 days after each vaccination is plotted by FDA toxicity grade, either as any grade (mild, moderate, severe, or potentially life-threatening) or as Grade 3+ (severe or potentially life-threatening).^20^

The most common solicited systemic AEs were headache, fatigue, myalgia, and malaise. These also were detected more frequently among NVX-CoV2373 recipients and after the second injection with a median duration of ≤2 days (range 1–7 days) (Table S11). Fever of any severity occurred in 16.9% of participants, largely NVX-CoV2373 recipients, especially after the second dose. Severe systemic reactions (≥Grade 3), most commonly fatigue, occurred in 3.7% vs. 3.4% after dose 1, and 22.0% vs. 3.4% after dose 2 in the NVX-CoV2373 and placebo groups, respectively (Figure 2 and Table S10). Similar reactogenicity rates occurred in the age subgroups (Figure S6).

#### Unsolicited AEs

Unsolicited AEs occurred with similar frequency in vaccine and placebo recipients (15.9% vs. 15.6%). Reports of MAAEs, SAEs, and severe AEs were balanced across treatment groups (Table S7). There were no safety events that triggered prespecified pause rules. No episodes of anaphylaxis, vaccine-enhanced Covid-19, Guillain Barré syndrome^22^, thrombosis with thrombocytopenia syndrome (TTS)^23^, or myocarditis/pericarditis^24^ were observed (Tables S12-S13). There were no deaths or AESI/PIMMC among adolescent trial participants.

### IMMUNOGENICITY

The ratio of NA response against SARS-CoV-2 wild-type virus at Day 35 for previously unexposed adolescents compared with that observed in similarly unexposed adult PREVENT-19 participants 18 to < 26 years of age met all criteria for non-inferiority: GMT ratio (GMTR) point estimate and 95% CI of LB: 1.5, 95% CI, 1.3 to 1.7, and LB of serologic response difference: - 1.0, 95% CI, -2.8 to 0.2 (Table 2).

**Table 2.** Neutralizing Antibody Response in Adolescents Compared to Young Adults in the PREVENT-19 Trial.*

| Age, y | No.<br>Participants | Geometric mean<br>titer at Day 35<br>(95% CI)† | Geometric<br>mean titer ratio<br>(95% CI)† | Serologic<br>response at Day<br>35, % (95% CI)‡ | Difference in<br>serologic<br>response (95%<br>CI)‡ |
| --- | --- | --- | --- | --- | --- |
| 12 - < 18 | 390 | 3860<br>(3423 to 4352) | 1.5 (1.3 to 1.7) | 98.7<br>(97.0 to 99.6) | -1.0 (-2.8 to 0.2) |
| 18 - < 26 | 416 | 2634<br>(2398 to 2904) |  | 99.8<br>(98.7 to 100) |  |

\* Neutralizing antibody titers of adult participants (18-<26 y) were compared to those from adolescents (12-<18 y). All participants in either age group were part of the per-protocol analysis set (i.e., SARS-CoV-2 unexposed participants who had at least a baseline and 1 serum sample result available after full primary vaccination and had no major protocol violations that were considered clinically relevant to impact immune response at the corresponding study visit, e.g., RT-PCR-positive swabs or seropositivity for SARS-CoV-2 prior to the visit in question).
† Geometric mean titers, geometric mean ratios and two-sided 95% confidence intervals were calculated based on the t-distribution of the log-transformed values for geometric means or GMFR, then back transformed to the original scale for presentation. Assay results below the lower limit of quantitation (200 EU/mL) were documented as 100 EU/mL (0.5 times the lower limit of quantitation). The non-inferiority criterion was met since the lower boundary of the two-sided 95% confidence interval for the geometric mean ratio was greater than 0.67, and the point estimate was equal or greater than 0.82.
‡ Serologic response was defined as a percentage of participants with a value ≥ 4-fold difference between Day 35 and Day 0. The 95% CI was calculated using the exact Clopper-Pearson method. The noninferiority criterion was met since the lower boundary of the two-sided 95% confidence interval for the difference of serologic response was greater than -10%.
Data source: validated microneutralization assay conducted by 360biolabs.<sup>16</sup>

NA GMTs and serologic response were markedly higher in vaccine than placebo groups across age subgroups (Figure S1). Day 35 serum IgG levels specific to wild-type and more recently emerged variants’ spike proteins and hACE2 RBI antibodies tested *post hoc* also demonstrated high antibody levels against all tested variants, including the Omicron subvariants BA.1, BA.2, and BA.5 (Figures S2–S5).

### EFFICACY

In the full analysis set (FAS), the incidence of Covid-19 in the placebo group was 9.86 cases per 100 person-years (95% CI 6.22 to 15.61), with cumulative incidence curves separating after day 21 (Figure 3). Among the 1,799 PP-EFF participants followed through September 27, 2021 (median surveillance time 64.0 days, range 1-135), 20 Covid-19 cases occurred (incidence 14.20 cases per 100 person-years [95% CI, 8.42 to 23.93] in placebo and 2.90 cases per 100 person-years [95% CI 1.31 to 6.46] in vaccine recipients). The 6 cases in NVX-CoV2373 recipients and 14 in placebo recipients yielded VE of 79.5% (95% CI, 46.8 to 92.1) (Table S5). All cases were mild in severity, thus VE against moderate-to-severe Covid-19 could not be established. Nasal swabs from 11/20 (55%) end point cases, 3 in vaccine and 7 in placebo recipients, yielded WGS. All (11, 100%) were identified as the Delta variant, yielding VE of 82.0% (95% CI, 32.4 to 95.2) (Table S6).

**Figure 3.**
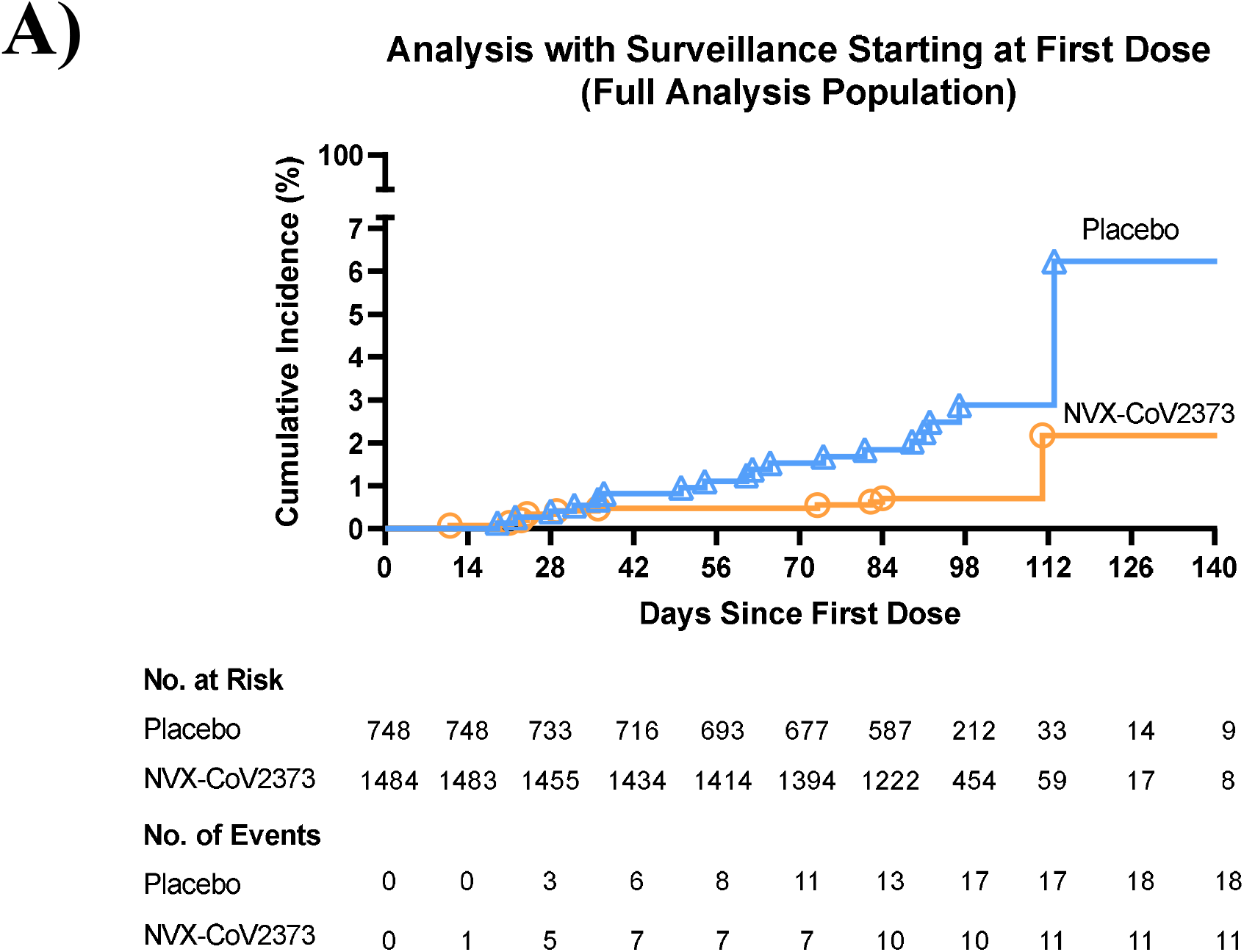
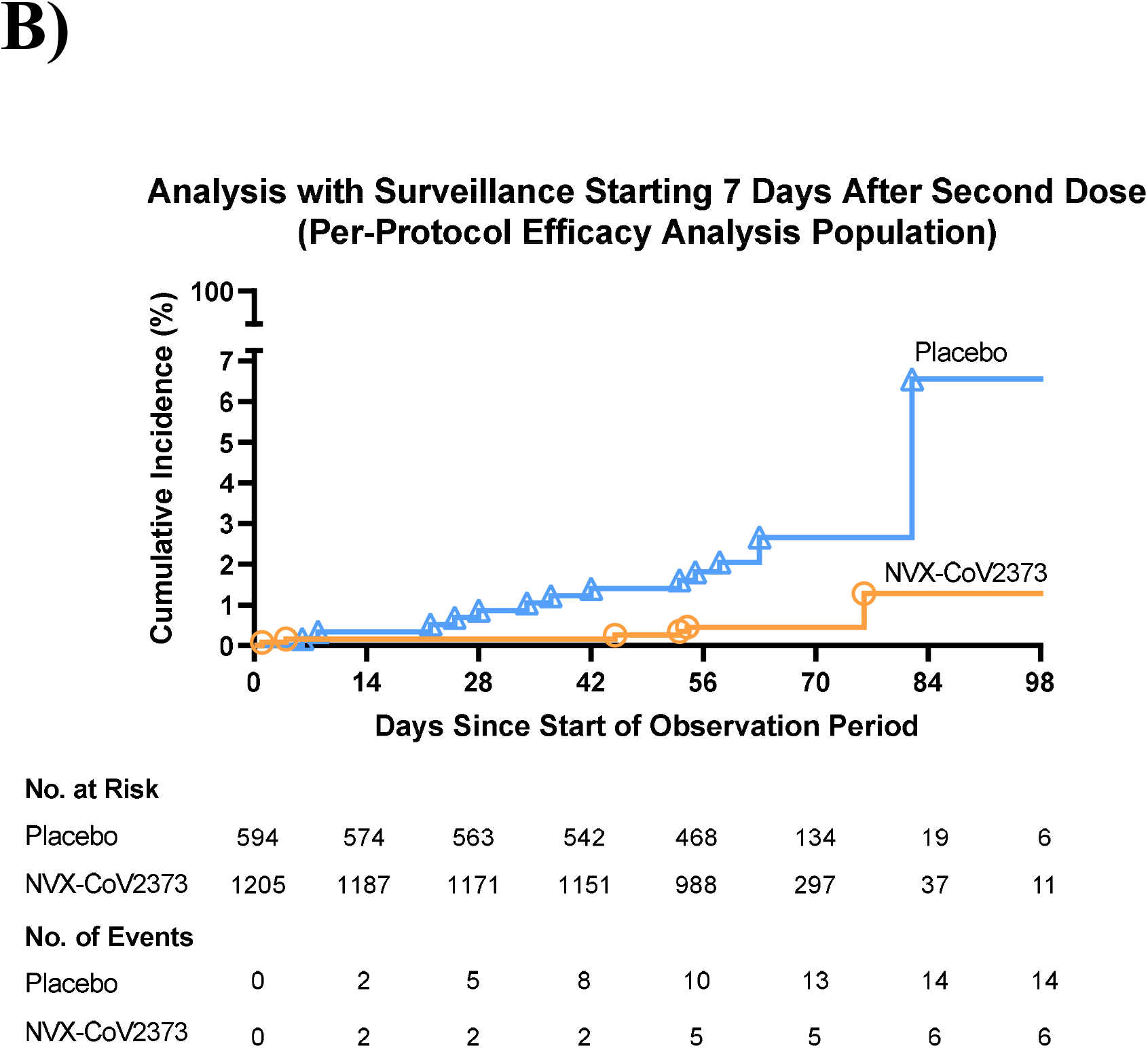
Cumulative Incidence Plot of Overall Efficacy of NVX-CoV2373 Against Symptomatic Covid-19. The time period of prospective surveillance of Covid-19 illness in the full analysis population was from the first dose of NVX-CoV2373 or placebo; per-protocol symptomatic Covid-19 cases were defined as beginning at least 7 days after the second dose (i.e., Day 28) through approximately 4 months of follow-up, or unblinding or receipt of Emergency Use Authorization vaccine. A) All participants, full analysis set; B) All participants, baseline seronegative/virologically negative, per-protocol population.

## Discussion

The pediatric expansion of the ongoing PREVENT-19 trial into over 2,200 racially and ethnically diverse adolescents in the United States demonstrated that NVX-CoV2373 appeared safe and effective (as determined by pre-defined immunogenicity criteria). NA responses on Day 35 post-vaccination were comparable for both GMFR and serologic response to those observed in young adults from PREVENT-19, in whom a high degree of protective efficacy had been demonstrated.^13^ Furthermore, protective efficacy of 79.5% (95% CI, 46.8 to 92.1) was demonstrated in the adolescents against the predominant circulating Delta variant.

The high short-term VE of NVX-CoV2373 for the prevention of Covid-19 in adolescents 12 through 17 years of age corroborated the earlier results from the adult portion of the study.^13^ These VE results were also consistent with those observed for mRNA vaccines in this age group. However, unlike phase 3 trials characterizing the efficacy of mRNA vaccines in adolescents,^25,26^ VE in this pediatric expansion was established during a period of almost exclusive circulation of the Delta variant, the only variant detected in all cases that yielded sequencing results (VE against Delta 82.0%, 95% CI, 32.4 to 95.2). Even though VE was specified as a descriptive analysis in the pediatric expansion, the results recapitulate the overall high VE observed for NVX-CoV2373 during earlier Phase 3 trials in adults,^12,13^ and provide additional evidence that the vaccine may elicit more broadly protective immunity to both contemporary and emerging SARS-CoV-2 variants.

No safety concerns were identified during the safety follow-up period reported (median 71 days post dose 2, with >84% of participants followed for at least 60 days for this analysis). Reactogenicity was mild-to-moderate in severity and, as expected, higher in frequency and severity after the second vaccination. By contrast, similar rates of unsolicited AEs (including SAE or severe AEs) between vaccine and placebo recipients were observed.

NAs and anti-S binding IgG antibodies at Day 35 (i.e., 14 days after the second vaccine dose) have been correlated with VE for NVX-CoV2373.^27^ High levels of humoral responses at Day 35 were observed in adolescents (as determined by both, anti-S IgG binding and functional MN and hACE2 RBI assays) against prototype SARS-CoV-2, as well as against more recent variants: Alpha, Beta, Delta, Gamma, Mu and Omicron, including subvariants BA.1, BA.2, and BA.5 (Figures S1–S5), which were 2-4 times higher than those observed in PREVENT-19 adult participants (Áñez et al, in preparation).

A limitation of this report is the short period of time (median surveillance time: 93 days) during which VE of the primary series of two doses of NVX-CoV2373 given 21 days apart could be evaluated. Placebo-controlled follow-up was limited by early implementation of a blinded crossover to ensure retention of this age group by providing active vaccination for all participants when other vaccines became available under emergency use authorizations.^13^ Notably, the protective efficacy was assessed during the predominant circulation of the Delta variant, which was later replaced by the Omicron variant. However, the *post hoc* analyses of immune responses against the more recent circulating Omicron subvariants supports likely effectiveness against a broad distribution of future emerging variants (Supplementary Appendix). The effect of a NVX-CoV2373 booster dose given 5–6 months after the primary series is being assessed for all PREVENT-19 participants, including those exposed to Omicron.^28,29^

NVX-CoV2373 has recently been authorized for emergency use by the FDA in adults and adolescents 12 years and older.^11,14^ The vaccine is expected to increase uptake in adolescents, >24% of whom have not yet received a full vaccination regime with mRNA vaccines.^30^ A favorable safety profile, storage and transportation requirements, and induction of broad, “universal”-like immune responses to provide protection against emerging variants, suggest that NVX-CoV2373 is an important tool in the fight against the current Covid-19 pandemic worldwide.

## Supporting information

CONSORT Checklist

## Data Availability

A data-sharing statement provided by the authors will be available with the full text of this article upon publication (https://clinicaltrials.gov/ct2/show/NCT04611802).

## [Disclosures]

Supported by Novavax, Inc., the Office of the Assistant Secretary for Preparedness and Response, Biomedical Advanced Research and Development Authority (BARDA) (contract number: OWS: Novavax’s Project Agreement No. 1 under its Medical CBRN Defense Consortium (MCDC) Base Agreement No. 2020-530; Department of Defense (DoD) No. W911QY20C0077); and the National Institute of Allergy and Infectious Diseases (NIAID), National Institutes of Health. The NIAID provides grant funding to the HIV Vaccine Trials Network (HVTN) Leadership and Operations Center (UM1 AI68614), the HVTN Statistics and Data Management Center (UM1 AI68635), the HVTN Laboratory Center (UM1 AI68618), the HIV Prevention Trials Network Leadership and Operations Center (UM1 AI68619), the AIDS Clinical Trials Group Leadership and Operations Center (UM1 AI68636), and the Infectious Diseases Clinical Research Consortium leadership group (UM1 AI148684).

No other potential conflict of interest relevant to this article was reported.

Disclosure forms provided by the authors will be available with the full text of this article upon publication.

## [Acknowledgments]

We especially thank all the study participants who volunteered for the study and who have contributed their clinical experience to the establishment of safety and efficacy of NVX-CoV-2373, as well as to the 2019nCoV-301 – Pediatric Expansion Study Group members (listed in the Supplementary Appendix); to the funders (BARDA, NIAID/NIH), the members of the NIAID Data and Safety Monitoring Board (DSMB), whose diligent monitoring of all trial data contributed to ensure the safety and well-being of the trial participants; community leadership groups throughout the country who assisted with community engagement and recruitment; and unnamed colleagues at each of the sites who generously contributed to the trials in many ways, and all unnamed colleagues at Novavax, Inc., who worked tirelessly and gave unlimited efforts to the development, testing, and support of this trial. Editorial assistance on the preparation of this manuscript was provided by Kelly Cameron, PhD, and Rebecca Harris, PhD, of Ashfield MedComms, an Inizio company, supported by Novavax, Inc.

