## Supplementary Appendix for "Safety, Immunogenicity and Efficacy of NVX-CoV2373 in Adolescents in PREVENT-19: A Randomized, Phase 3 Trial"

**Table of Contents**

2019nCoV-301 Pediatric Expansion Principal Investigators and Study Team ………….………………	7

Data and Safety Monitoring Board (DSMB) Members List ..…………………………………………..	11

Objectives and End Points ..……………………………………………………………………………..	12

Table S1. Primary and Secondary Objectives and End Points Addressed in This Manuscript. Protocol Version 8.0. ……………………………………………………………………………………………..	12

Supplemental Methods ……………………………….……………………………….….….………......	15

Safety Assessments ……………………………………………………………………………..	15

Safety Monitoring ……………………………………………………………………………….	15

Prospective Surveillance of Covid-19 .……………………………………………………….....	15

Covid-19 End Point Assessment .……………………………………………………………….	16

Nasal Swabs for Viral Detection ………………………………………………………………..	17

SARS-CoV-2 RT-PCR Testing ………………………………………………………………...	17

Whole-Genome Sequencing (WGS) and Clade/Lineage Assignment ………………………….	18

Anti-S IgG Antibody Titer Determination (ELISA) [fit-for-purpose assay] …………………...	18

hACE2 Receptor Inhibiting Antibody Titer Determination (ELISA) [fit-for-purpose assay] ….	19

Analysis Populations .…………………………………………………………………………...	20

Statistical Method for Efficacy End Points ……………………………………………………..	22

Statistical Method for Effectiveness ……...…………………………………………………….	23

Supplemental Tables and Figures ………………………………………….……………………………	24

Table S2. Symptoms Suggestive of Covid-19 …………………………………………………………...	24

Table S3. End Point Definitions of Covid-19 Severity ………………………………………………….	24

Table S4. Demographics and Baseline Characteristics (Safety Analysis Set) .………………………….	26

Figure S1. A) Box Plot of Neutralizing Antibody Titers for SARS-CoV-2 Wild-Type Virus at Specified Time Points in Baseline Serologically Negative/PCR-negative Adolescent Participants 12 to < 18 Years of Age (PP-IMM Analysis Set), B) Participants 12 to < 15 Years of Age (PP-IMM Analysis Set), and C) Participants 15 to < 18 Years of Age (PP-IMM Analysis Set) ……………………..…………………...	27

Figure S2. Box Plot of Serum IgG Antibody Concentrations to SARS-CoV-2 S Protein in Baseline Serologically Negative/PCR-negative Adolescent Participants 12 to < 18 Years of Age (PP-IMM Analysis Set) …………………………………….....……………………………………………………	29

Figure S3. Box Plot of hACE2 Inhibition Antibodies to SARS-CoV-2 S Protein in Baseline Serologically Negative/PCR-negative Adolescent Participants 12 to < 18 Years of Age (PP-IMM Analysis Set) …...	30

Figure S4. Serum IgG Antibody Concentrations to SARS-CoV-2 S Protein From Different Variants in Baseline Serologically Negative/PCR-negative Adolescent Participants 12 to < 18 Years of Age (N=40, PP-IMM Analysis Set, *Post Hoc* Analysis, Non-Validated Fit-for-Purpose Assay) ……………..……..	31

Figure S5. hACE2 Inhibition Antibodies to SARS-CoV-2 S Protein From Different Variants in Baseline Serologically Negative/PCR-negative Adolescent Participants 12 to < 18 Years of Age (N=40, PP-IMM Analysis Set, *Post Hoc* Analysis, Non-Validated Fit-for-Purpose Assay) ……………………………...	32

Table S5. Vaccine Efficacy Against RT-PCR-Confirmed Symptomatic Mild, Moderate, or Severe Covid-19 at Least 7 Days After Second Vaccination Due to Any SARS-CoV-2 Variant in Adolescent Participants Not Previously Exposed to SARS-CoV-2 (PP-EFF Analysis Set) ……………………….	33

Table S6. Vaccine Efficacy Against RT-PCR-Confirmed Symptomatic Mild, Moderate or Severe Covid-19 at Least 7 Days After Second Vaccination Due to the SARS-CoV-2 Delta Variant in Adolescent Participants Not Previously Exposed to SARS-CoV-2 (PP-EFF Analysis Set) ………………………...	34

Table S7. Overall Summary of Treatment-Emergent Adverse Events Reported Between Start of First Vaccination and Blinded Crossover or Early Withdrawal (Safety Analysis Set) ………………………	35

Figure S6. Solicited Local and Systemic Adverse Events by Age Subgroup………………………….	36

Table S8. Summary of Solicited Local Adverse Events Within 7 Days After Dose 1 and Dose 2 in All Participants (Safety Analysis Set) …………………………………….….….…………………………..	37

Table S9. Duration (Days) of Solicited Local Adverse Events Within 7 Days After Dose 1 and Dose 2 in All Participants (Safety Analysis Set) …………………………………….……..………………………	38

Table S10. Summary of Solicited Systemic Adverse Events Within 7 Days After Dose 1 and Dose 2 by Age Group (Safety Analysis Set) …………...……………………….….……………….….…………...	39

Table S11. Duration (Days) of Solicited Systemic Adverse Events Within 7 Days After Dose 1 and Dose 2 in All Participants (Safety Analysis Set) ………………………………..…..….……………………...	41

Table S12. Overall Summary of Unsolicited Adverse Events by System Organ Class and Preferred Term Reported Within 49 Days After First Vaccination in at Least 0.5% of All Adolescent Participants in Any Study Vaccine Group by Age Strata (Safety Analysis Set)..…………………………………………….	43

Table S13. Overall Summary of Unsolicited Serious Adverse Events From Start of First Vaccination to Blinded Crossover Dose in All Adolescent Participants in Any Study Vaccine Group by Age Strata (Safety Analysis Set)……..……………………………………….…...…………………………………	45

References .……………………………………………………………………………..………………..	46

**2019nCoV-301 Pediatric Expansion Study Group (Pubmed Listed, in Alphabetical Order of Institution Affiliation)**

| **Affiliation/Funding*** | **Study Group** | **Location** |
| --- | --- | --- |
| **United States and Puerto Rico** | | |
| Accel Research Sites | James Andersen, MD, Szheckera Fearon, MSN, FNP-C, Rosa Negron, MD, Amy Medina, ADN, BS, Colleen Figueroa, Courtney Smith | Lakeland, FL |
| Accel Research Sites | Bruce Rankin, DO, John M. Hill, MD, Steven Shinn, MD, Vivek Rajasekhar, DO, Marshall Nash, MD, Ashraf Affan, MD | DeLand, FL |
| Acevedo Clinical Research Associates | Armando Acevedo, MD, Alina Monteagudo Can, MD, Hector Rodriguez, MD, Israel Zagales, Christine Prieto, Lizz Hernandez Diaz | Miami, FL |
| Alabama Clinical Therapeutics | Max Hale, MD, Patrick Farr, MD, Liesel French, MD, Teresa Goldsmith, MD, James Warmack, MD, Bailey Murphy | Birmingham, AL |
| Alliance for Multispecialty Research | Robyn Hartvickson, MD, Brooke Dunlavy, MD, Richard Glover, MD, Amber Grant, MD, Troy Holderman, MD, Stacy Slechta, MD | Newton, KS |
| Alliance for Multispecialty Research | Terry Poling, MD, Terry Klein, Thomas Klein, Tracy Klein, Sarah Pinkham, Shannen Lassiter | Wichita, KS |
| Asclepes Research Centers | Imad Jandali, MD, Maryam Belavilas, MD, David Daniels, Katie Leonard, Abigail Vetter, Toni Rich | Spring Hill, FL |
| Aventiv Research | Samir Arora, MD, Grazia Cannon, MD, Sridhar Guduri, MD, Alandra Lingel, MD, Veronica Moore, MD, Sarah Wilcox, MD | Columbus, OH |
| Biomedical Advanced Research and  Development Authority (BARDA) | Richard Gorman, MD, Gary Horwith, MD, Robin Mason, MS, MBA | Washington, DC |
| Benchmark Research | Laurence Chu, MD, Michelle Listz, Lamar Box, Cindy Duran, Isaiah Knight, Katherine Davis | Austin, TX |
| Benchmark Research | Greg Hachigian, MD, Deborah Murray, FNP, Michael Cancilla, PA, Logan Ledbetter, PA, Masaru Oshita, MD | Sacramento, CA |
| Benchmark Research | William Seger, MD,  John Villegas, Ben Seger, Virginia Loudermilk, Ruth Reyes, Anthony Kim | Fort Worth, TX |
| Brownsboro Park Pediatrics | Wendy C Daly, MD, Rebecca Becherer, MD, Denver Cornett, MD, Karen Dick, MD, Kimberly Downs, MD, Pamela Hall, MD | Louisville, KY |
| California Research Foundation | Donald M. Brandon, MD, William B. Davis, MD, Daniel T. Lawler, MD, Cindy Stevens, MD, Karl Walter, MD, Michelle Rios | San Diego, CA |
| Cenexel RCA | Nelia Sanchez-Crespo, Thelma Beltron, Jennifer Schwartz, Patricia Balebona, Beatriz Rivera, Barbara Garcia, | Hollywood, FL |
| Charlotte-Mecklenburg Hospital Authority d/b/a Atrium Health / NIAID (UM1AI068614) | Christine B. Turley, MD, Andrew McWilliams, MD, Tiffany Esinhart, PA-C, Natasha Montoya, APRN, Shamika Huskey, FNP, Leena Paul, FNP | Charlotte, NC |
| Clinical Neuroscience Solutions | Michael E. Dever, MD, Mitul Shah, MD, Michael Delgado, MD, Tameika Scott, DrPH, Patricia Brown, PhD, Americo, Padilla, MD | Orlando, FL |
| Clinical Neuroscience Solutions | Lisa S. Usdan, MD,  Lora J. McGill, MD , Valerie K. Arnold, MD , Carolyn Scatamacchia, MSN, NP-C , Codi Anthony, DNP, APRN, PMHNP-BC , Robyn Presley | Memphis, TN |
| Clinical Research Associates | Stephan Sharp, MD, Michael Caldwell, MD, Linda Schipani, RN, Allison Ulrich, RN, Wendy Tidwell, Stacy Lynn Cox, APN, RN | Nashville, TN |
| Clinical Research Center of Nevada | Michael Levin, MD, Julia Gass, Marcy Kulic, MD, Eduardo Rodriguez, Jessica Corea, Sierra Dansbee | Las Vegas, NV |
| Clinical Research Institute of Southern Oregon | John Delgado, MD, Jaleh Ostovar, NP, Audrey Kuehl, Sarah Smiley, Danuel Hamlin, Ben Taucher | Medford, OR |
| Coastal Carolina Research Center | Cayce Tangeman, MD, Yvonne Davis, MD, Vanessa Armetta, Mary Love, David Summers, RN, J. Bruce Etheridge, MD | North Charleston, SC |
| Coast Clinical Research | Teresita Salazar, MD, Femina David, MD, Filipinas Vitug, Amiel Guevarra, Noemi Ramirez | Bellflower, CA |
| Comprehensive Clinical Research | Ronald Ackerman, MD, Jamie Ackerman, Florida Aristy, APRN | West Palm Beach, FL |
| Covid-19 Prevention Network (CoVPN) | Lawrence Corey, MD, Kathleen M Neuzil, MD, MPH, Huub G Gelderblom, MD, PhD, Nzeera Ketter, MD, Carrie Sopher | Seattle, WA |
| DM Clinical Research | Vicki E. Miller, MD, Amy Starr, Sonia Guerrero, Madiha Baig, Maryam Jamil, Husain Motiwala | Tomball, TX |
| DM Clinical Research - Pediatric Healthcare of NW Houston | Khozema Palanpurwala, MD, Monica Murry, MD, Amy Starr, MD, Meghan Tonti, MD, Rebecca Wischnewsky, MD, Earl Martin, MD | Houston, TX |
| Empire Clinical Research | Yogesh K. Paliwal, MD, Amit Paliwal, MD, Sarah Gordon, MS, Krystle Edwards, Cynthia Montano-Pereira, Michael Campos, MD | Pomona, CA |
| Health Research of Hampton Roads | George H. Freeman, MD , Esther Laverne Harmon, ANP , Marshall A. Cross, MD , Kacie Sales, BSN, RN,  Catherine Q. Gular, PharmD | Newport News, VA |
| Holston Medical Group | Joseph Ley, MD, Amanda Donoho, MD, Kimberley Hunt, MD, Donald Lewis MD, Stephanie Tipton, MD, Emily Whitaker, MD | Kingsport, TN |
| Jacksonville Center for Clinical Research | Jeffry Jacqmein, MD , Maggie Bowers, PA-C , Dawn Robison, APRN-C,  Victoria Mosteller, MD , Janet Garvey, DNP | Jacksonville, FL |
| Johnson County Clin-Trials | Carlos Fierro, MD, Mary Easley, BSN, RN , Amy Thompson, MD, Heather Barker, Mazen Zari, Karol Moore | Lenexa, KS |
| Kentucky Pediatric/Adult Research | Daniel Finn, MD, Lindsay Blackman, MD, Stanley, Block, MD, Christal Denton, MD, Martha Osborn, MD, Robert Smith, MD | Bardstown, KY |
| Lynn Health Science Institute | Carl P. Griffin, MD, William Schnitz, MD, Raymond Cornelison, MD, Linda Lopez, Kim Hamilton, Kim Calloway | Oklahoma City, OK |
| Triangle Orthopaedic Associates, PA | David B. Musante, MD , William P. Silver, MD,  Linda R. Belhorn, MD,  Nicholas A. Viens, MD,  David Dellaero, MD, Elizabeth Wilkers, MD | Durham, NC |
| MedPharmics | Robert Jeanfreau, MD, Nicki Johnson, Estafania Bazan, Davilyn Roys, Steven Darden, Susan Jeanfreau, MD | Metairie, LA |
| MedPharmics | Paul G. Matherne, MD, Amy Caldwell, RN, Jessica Stahl, RN, Nicole Guttierrez, RN, Cassandra Beeks LPN | Gulfport, MS |
| Meridian Clinical Research | Frank Eder, MD, Ryan Little, MD., Susan Owen, MD, Heather Shaw, MD, John Tarbox, MD, Victoia Engler | Binghamton, NY |
| Meridian Clinical Research | Brandon Essink, MD, Roni Gray, APRN , Fritz Raiser, DO, Christine Wilson , Tiffany Nemecek, Hannah Harrington, MPH | Omaha, NE |
| Meridian Clinical Research | Charles Harper, MD, Torie Johnson, Chelsie Nutsch, NP, Sally Eppenbach, NP, Wendell Lewis, NP, Katlyn Mace | Norfolk, NE |
| Meridian Clinical Research | Brannon C Perilloux, MD, Christopher Dedon, MD, Lori Cook, Brittany Zedlitz, Vasavi Srinivasan, Paige Melner | Baton Rouge, LA |
| Meridian Clinical Research | Joanna Sextor, MD, Jessica Long, MD, Francis Palumbo, MD, Devon Myers, Caroline Vleck, MD, Peter Warfield, Dr | Washington, DC |
| National Institute of Allergy and Infectious Diseases (NIAID) / National Institutes of Health (NIH) | Tatiana Beresnev, MD, Maryam Jahromi, MD, Mary A. Marovich, MD, Julia Hutter, MD, Martha Nason, PhD | Bethesda, MD |
| National Research Institute | Mark Leibowitz, MD,  Fernanda Morales,  Mike Delgado, Rosario Sanchez, Norma Vega | Los Angeles, CA |
| Novavax, Inc. | Germán Áñez, MD, Lisa M. Dunkle, MD, Gary Albert, Erin Coston, Chinar Desai, Haoua Dunbar, Mark Eickhoff, Renee Entzminger-Sneed, Jenina Garcia, Margaret Kautz, Angela Lee, Maggie Lewis, Alice McGarry, Patrick Newingham, Patty Price-Abbott, Patty Reed, Kimberly Cerenze Short, Diana Vegas, Bethanie Wilkinson, PhD, Katherine Smith, MD, Wayne Woo, MS, Iksung Cho, MS, Gregory M. Glenn, MD, Filip Dubovsky, MD, MPH | Gaithersburg, MD |
| Velocity Clinical Research | David L. Fried, MD, Lynne A. Haughey, MSN, FNP, Ariana C. Stanton, PA-C, Monica Freeman, Jacqueline DiFazio, Courtney Langlois | Warwick, RI |
| Orange County Research Institute | Rosario Retino, MD, Alexis Deniz, MD, Wendy Paiva, MD, Leonel Pajarillaga, MD, Lisa Pyio, MD, LendyTorres, Dr | Ontario, CA |
| Pediatric Research of Charlottesville | Paul Wisman, MD, Carlos Armengol, MD, Gemia Bouber, MD, Peggy Bressler, MD, Alaina Brown, MD, Candyce Dorsey, Dr | Charlottesville, VA |
| PMG Research of Bristol | Bernard Grunstra, MD, Amy Dye, Shelby Gilmer Olds, Joshua Bullen, Miranda Roark, Jennie Eller | Bristol, TN |
| Ponce School of Medicine / NIAID (UM1AI148685) | Elizabeth Barranco-Santana, MD, Jessica Rodriguez, MD, Rafael Mendoza, MD, Karen Ruperto, MD, Odette Olivieri, MD, Enrique Ocaña, MD | Ponce, Puerto Rico |
| Preferred Primary Care Physicians | Bryce Palchick, MD, Nathan Bennett, MD, Sarah Sobrosky, MD, Michael Gates, MD, Suzanne Klutch, MD, Jessica Martier, Dr | Pittsburgh, PA |
| Premier Health Research Center | Marilou Cruz, MD, Hoang-Chuing Vu MD, Janet Serrano, Valerie Martinez, Amiel Guevarra, Marc Cruz | Downey, CA |
| Providence Clinical Research | Teresa S. Sligh, MD, Parul Desai, NP, Vincent Huynh, BSc, Carlos Lopez, MD, Erika Mendoza, BA | North Hollywood, CA |
| Research Your Health | Jeffrey Adelglass, MD, Waseem Chughtai, BS, MBBS, Anuja Sathe, MS, Pamela Fox | Plano, TX |
| Rochester Clinical Research | Matthew G. Davis, MD, Jennifer Foley,  Shelly Kane, MD, Cassidy Glod, Marissa Wuilliez, Abigail Purcell | Rochester, NY |
| Senders Pediatrics | Shelly Senders, MD, Ashley Jeffrey, Christopher Fackelmann, Nicholas Jezerinc, Caitlin Fillioe, Timothy Hudec | Cleveland, OH |
| Spartanburg Medical Research | Charles Fogarty, MD, Nicole Crockford, MD, Jami Jones, Connie Mccauley, Sherry Yeisley, Alison Fogarty | Spartanburg, SC |
| Sterling Research Group | Bruce C. Gebhardt, MD,  Padma N. Mangu, MD, Debra Beck Schroeck, MS, PA-C,  Rajesh Kumar Davit, MD,  Gayle D. Hennekes, PA-C, MPAS | Cincinnati, OH |
| Sundance Clinical Research | Larkin Tyler Wadsworth III, MD, Horacio Marafioti, MD, Lyly Dang, DNP-BC, Lauren Clement, NP-C, Jennifer Berry, FNP-BC | St. Louis, MO |
| Tekton Research | Adebayo Akinsola, MD, Gabriela Baetista, MD, Baba Arimah, Sheree Dover, Leslie Hernandez, Susan Herrera, | Chamblee, GA |
| Tekton Research | Kenneth Etokhana, MD, Nathaniel De La Cruz, Xavier Fajardo, Veronica Galaviz, Meranda Ruiz, Tony Diaz | San Antonio, TX |
| Texas Center for Drug Development | Veronica Garcia-Fragoso, MD,  Maria Gabriela Becerra, MD,  Cecilia Mckeown , Lisa Holloway,  Stacey Montero, Tracy Kowalsi | Houston, TX |
| University of California Davis Health / NIAID (UM1AI068614) | Stuart H. Cohen, MD, Monica Ruiz, Dean M. Boswell, BS, Elizabeth E. Robison, BS, Trina L. Reynolds, BS, Sonja Neumeister, MPH | Sacramento, CA |
| University of Colorado Hospital CRS / NIAID (UM1AI068636) / NCATS (UL1TR002535, UM1AI069432) | Thomas B. Campbell, MD, Suzanne Fiorillo, MSPH, Rebecca Pitotti, RNP, Victoria Riedel Anderson, MS, Jose Castillo Mancilla, MD, Nga Le, PharmD | Aurora, CO |
| University of Maryland School of Medicine / NIAID (UM1AI148689) | James D. Campbell, M.D., M.S., Milagritos Tapia, MD, Karen L. Kotloff, MD, Kathleen Neuzil, MD, Andrea Berry, MD, E. Adrienne Hammershaimb, MD, MS, Rosa MacBryde, RN | Baltimore, MD |
| University of Nebraska Medical Center / NIAID (UM1AI068614) | Diana F Florescu, MD, Richard Starlin, MD, David Kline, MD, Andrea Zimmer, MD, Anum Abbas, MD, Natasha Wilson, APRN | Omaha, NE |
| University of North Carolina / NIAID (UM1AI068619) / University of North Carolina at Chapel Hill Center for AIDS Research (P30AI050410) / NC TraCS Institute (UL1TR002489) | Cynthia L. Gay, MD, MPH, Erin Hoffman, Carolina Pastrana Medina, Susan Pedersen, Mandy Tipton, Alison Burbank, MD, Michelle Hernandez MD, Peyton Thompson MD, Zachary Willis MD, Joseph Eron, MD. | Chapel Hill, NC |
| University of Washington | Robert W. Coombs, MD, PhD, Alex Greninger, MD, PhD, MS, MPhil, Pavitra Roychoudhury, PhD, Erin A. Goecker, MS, Yunda Huang, PhD, Youyi Fong, PhD | Seattle, WA |
| Velocity Clinical Research | Robert J. Buynak, MD, Angella Webb, APRN, Rena Rivas, Stephanie Andree, FNP, Rachel McNeal, Megan Smith, MD | Valparaiso, IN |
| Velocity Clinical Research | Julie Kasarjian, MD, Judith Kirstein, MD, Krista Foster, Nicole Abels, Brandy Lopez, Crystle Rajania, Dr | Banning, CA |
| Velocity Clinical Research | Margaret Rhee, MD, Gabrielle Jones, Alanna Billups, Jane Boggan, Denise Roadman, PAC, Celeste Blazy | Cleveland, OH |
| Velocity Clinical Research | Marian E. Shaw, MD, Mark A. Turner, MD, Cory J. Huffine, FNP-C, Esther S. Huffine, FNP-C, Raymond Coon, Jacqueline Hanson | Meridian, ID |
| Velocity Clinical Research | Michael Waters, MD, Karla Zepeda, NP, Scott Overcash, MD, Jordan Coslet, NP, Dalia Tovar, MA, Kia Lee | Chula Vista, CA |
| Ventavia Research Group | Mark Koch, MD, Norma Escamilla, MD, Lydia Luna, MD, Erin Mcleod, Kathryn Dykes, Katy Dykes | Fort Worth, TX |
| Walter Reed Army Institute of Research | Julie A. Ake, MD, MSc | Silver Spring, MD |
| Wee Care Pediatrics | Michael Husseman, MD, Janes Fennell, MD, Jennifer Gilsoul MD, Robert Hoki, MD, Ashley MacDonald, MD, Mia Lobato | Layton, UT |
| Wee Care Pediatrics | Peter Silas, MD, Cody Hawkes, MD, Jennifer Nelson, MD, Jennifer Cooper, Jennifer Nelson, MD, Jerica Twitchell, RN | Syracuse, UT |
| Foothills Research Center | Kenneth Steil, MD, Mildred DeJesus, MD, LaShonda Gilbert, MD, Carey Goldsmith, MD, Maria Gustilo, MD, Jaimi Jones, Dr | Phoenix, AZ |
| WR Clinsearch | Mark McKenzie, MD,  Teresa Deese,  Mitsi Earwood , Vickie Leathers , Diane Sproles, Obadias Marques, Dr | Chattanooga, TN |

* Funding of institutions by the National Institute of Allergy and Infectious Diseases (NIAID) and/or research support by the National Center for Advancing Translational Science (NCATS), as indicated. All other institutions were funded by Office of the Assistant Secretary for Preparedness and Response, Biomedical Advanced Research and Development Authority. The content of this publication is solely the responsibility of the authors and does not necessarily represent the official views of the funding sources.

**2019nCoV-301 Pediatric Expansion Principal Investigators and Study Team (in alphabetical order)**

| **Principal Investigator** | **Study Team** | **Institution** | **Location** |
| --- | --- | --- | --- |
| Armando Acevedo, MD | Angelica Alvarez, Martha Falson, Blake Fernandez, Elizabeth Garcia Vallina, Robert Grant, Lizz Hernandea Diaz, Irus Izquierdo, Alina Monteagudo, Christine Prieto, Hector Rodriguez, Carmen Vargas, Isreal Zagales, | Acevedo Clinical Research Associates | Miami, FL |
| Ronald Ackerman, MD | Jamie Ackerman, Joahua Ackerman, Ronald Ackerman, Florida Aristy, Michael Bruck, Tomeko Heard, Diana Mann, Maureen Stewart, Cheryl Demczyk, Rohan Barron, Ashley Torres, Jennifer Gomez, Tiffany Potter | Comprehensive Clinical Research | West Palm Beach, FL |
| Jeffrey Adelglass, MD | Waseem Chughtai, Anuja Sathe, Cameron Galownia, Ramiro Lopez, Erica Parker-Martinez, Helene Harrison, Chiedza Mutindori, Sabrina Flowers, Tamara Betters, Carolyn Ackley, Pamela Fox, Noelia Tejada James, Dorothy Saylor, Hallen Dao, Jon Etta Randolph, Jason Tentativa, Malaika Chughtai, Shanzae Chughtai, Maheen Shah, Hayyan Chughtai, Tyler Love, Ti'arah Love, Daylia Hollins | Research Your Health | Plano, TX |
| Adebayo Akinsola, MD | Gabriela Bautista, Babatunde Arimah, Jazmine Roman, Arlene Torres, Sydnei Simpson, Analy Calvillo, | Tekton Research | Chamblee, GA |
| James Andersen, MD | Szheckera Fearon, Rosa Negron, Amy Medina, Diana Holmes, Colleen Figueroa, Cristal Ruiz, Nancy Masseus Tare Floyd, Kenta Oliver, Candice Gerber, Mae Ann Francisco, Gilbert de la Cruz, Ginny McClanahan, Veronica Walker, David Irwin, Gloria Adejobi | Accel Research Sites | Lakeland, FL |
| Samir Arora, MD | Kristina Susi, Maragaret Rood, Alex Lingel, Roger Faubel, Jordan Carpenter, Allison Forsythe, Sarah Wilcoox, Grazia Cannon, Ridhar Guduri, Veronica Moore | Aventiv Research | Columbus, OH |
| Elizabeth Barranco-Santana, MD | Michele Irizarry, Alice Grace Rodriguez, Irmaris Arroyo, Sara Cancel, Alejandra Román, Juan D. Lugo, Armando X. Torres, Marianne Hernandez, Brenda Garcia, Nancy Jiménez, Orlando Torres | Ponce School of Medicine | Ponce, Puerto Rico |
| Donald M. Brandon, MD | William B. Davis, Daniel T. Lawler, Maria Aceves, Kathleen B. Anderson, Janice E. Brandon, Patricia A. Brandon, Lorraine Boggs, Charlene Cruz, Mairead Hawkins, Clarice Hranicky, Karen G. McCrea, Cindy F. Stevens, Hannah J. Zapata, Karl Walter, Lorraine Boggs, Janice Brandon, Jeffery Brandon, Claire Hranicky, Tierney O’Connor, Michelle Rios, | California Research Foundation | San Diego, CA |
| Robert J. Buynak, MD | Mark Yarosz, Rachel McNeal, Megan Smith, Patricia Volom, Erica Lewis, Miranda Lee, Destiny Williams, Jessicah Johnson, Priscilla Dodson, Stephanie Andree, Angella Web, Yesenia Chavez, Hannade Ibrahim | Velocity Clinical Research | Valparaiso, IN |
| James D. Campell, MD, MS | Rosa MacBryde, Myounghee Lee, Andrea Berry, Milagritos Tapia, Karen Kotloff, DeAnna Friedman-Klabanoff, Mark Travassos, Meagan Deming, E. Adrienne Hammershaimb, Mary Rackson, Megan McGilvray, Cheryl Young, Colleen Boyce, Shirley George, Nancy Greenberg, Ginny Cummings, Rosa MacBryde, Kaitlin Mason, Susan Holian, Laura Liberman, Debra Houck, Jennifer Winkler, Rebecca Hudson, Rachel Riser, Leida Flores-Pineda, Delle Vazquez, Megan Pawtowski, Samantha Maynard, Michelle Ramirez, Constance Thomas, Mary Griffin, Audreda Flythe, Jennifer Marron, Kelly Brooks, Lisa Turek, Patricia Farley, Shantel Frels, Sherry McCammon, Panagiota Komninou, Daryl Grays, Chidozie Obichere, Julieta Nacario, Barbara Sinagra, Arren Gapasin, Leona Gittens, Shernel Barrett, Shannon Bittner, Joanne Snow, Jadzia Floyd, Josh Hend, Melissa Marini, Ifayet Mayo, Kathy Strauss, Sudhaunshu Joshi, Cheilon Bolanos, Kristen Krebs, Moishe Lerner, Sarah Hart, Brenda Dorsey, Alyson Kwon, Sophie Harper, Myounghee Lee, Phuong Tran Nguyen, Deanna Brandford, Jessica Moore, Leslie Howe, Barbara Albert, Yi Esparza, Maria Schanz, Jennifer German, Natelaine Fripp, Alexander Herrera, Dae Hyoun Jeong, Natasha Harris, Anne Thurston, Elizabeth Johnson, Nancy Wymer, Sandra Molina | University of Maryland School of Medicine | Baltimore, MD |
| Thomas B. Campbell, MD | Donna McGregor, Laurel Ware, Myron Levin, Steven Johnson, Sophia Quesada, Martin Krsak, Kristine Erlandson, Nicholas Sarchet, Vanessa Sutton, Lawrence Moran, Tracey Stevenson, Alaina Dougherty, Julianne Randlemon | University of Colorado Hospital CRS | Aurora, CO |
| Laurence Chu, MD | Marie Barrientes, Lamar Box, Michael Chouteau, Christal Lauren, Katherine Davis, Ronesha Davis, Cindy Duran, Ruth Fitch, Brooke Harris, Lisa Johnson, Isaiah Knight, Jennifer Leyva, Michelle Listz, Brandon Newsom, Marisol Ramos, Jessica Ruff, Victoria Sweet, Francesca Vigil, Taylor Wilson | Benchmark Research | Austin, TX |
| Stuart H. Cohen, MD | Curtis Blankenship, Katelyn Trigg, Courtney Lymuel, Gursimran Mann, Zayan Musa, Hana Minsky, Eliseo Vasquez, Nicole Garza, Kaitlyn Low, Mehrab Hussain, William Li, Rahul Araza, Monique Conover, George Thompson, Hien Nguyen, Scott Crabtree, Bennett Penn, Minh-Vu Nguyen, Archana Reddy, Derek Bays, Kaitlyn Hardin, Matthew Boutros, Alan Koff, Natascha Tuznik, Angel Desai, Naomi Hauser, Sarah Waldman, Gauri Barlingay, Dean Blumberg | UC Davis Health | Sacramento, CA |
| Marilou Cruz, MD | Hoang-Chuong Vu, Janet Serrano, Valerie Martinez, Amiel Guevarra, Marc Cruz, Letitcia Rodriguez | Premier Health Research Center | Downey, CA |
| Wendy C Daly, MD | Rebecca Becherer, Denver Cornett, Karen Dick, Kimberly Downs, Pamela Hall, Renee Heustis, Renee Lord, Deborah Massey-Eyre | Brownsboro Park Pediatrics | Louisville, KY |
| Matthew G. Davis, MD | Helen Bell-Upsher, Alyssa Birkbeck, Ann Casey, Keli Collazo, Lynsey Collins, Edward Coomber, Therese Dayton, Carolyn Dipoala, Brandy Douglass, Kathleen Ebeling, Jessica Fisher, Jennifer Foley, Cassidy Glod, Cherie Hallinan, Gillian Hess-Caple, Deborah Hodges, Maurice Holmes, Sarah Ingalsbe-Geno, Shelly Kane, Jean Kelly, Patricia Larabee, Linda Link, Chelsey LoMonaco, Michelle Lurvey, Jordan Lysons, Stefanie Mangefrida, Joseph Mann, Alison Meyers, Julie Mooney, Marie Musolino, Alicia Olena, Ashley Osinski, Abigail Purcell, Robin Spadafora, Marissa Wuilliez, Jaclyn Zona Zerina Zornic | Rochester Clinical Research | Rochester, NY |
| John Delgado, MD | Jaleh Ostovar, Alison Alvord, Levi Gadbois, Danuel Hamlin, Audrey Kuehl, Ryan Rackey, Sarah Smiley, Ben Taucher, Francisca Villa-Rodriguez, Jay Weisbart | Clinical Research Institute of Southern Oregon | Medford, OR |
| Michael E. Dever, MD | Michael Delgado, Tameika Scott, Laverne Denise Davila, Nelisa Frias, Anissa Hilton, Patricia Brown, Shana Caldwell, Martha Hendrix, Edmund Delgado, Mitul Shah, Gracemarie Rosario, Kaneitra Williamson, Taylor Lucier, Jaime Hawat, Matthew Stephens, Monica Cooper, Dante Canidate, Denise Pagan, Sierra Robinson, Pascal Nelson-Quiles, Anthony Perez, Chanel Adams, Keisha Foster, Scott Salmon, Andrew Lockwood, Priya Moorhouse, Paul Yi | Clinical Neuroscience Solutions | Orlando, FL |
| Frank Eder, MD | Heather Shaw, Ryan Little, Victoria Engler, Ashlehy Conover, Abigail Wine, Jennifer Molstead, Carolyn Grausgruber, Tammy Kohn, Traci Hull, Tarin Gordon, Kerry Hobarth, Nicole Croft, John Tarbox, Stacy Zudoffsky | Meridian Clinical Research | Binghamton, NY |
| Brandon Essink, MD | Hannah Harrington, Samantha Soule, Hannah Panec, Tiffany Nemecek, Christine Wilson, Jessica Satorie, Roni Gray, Fritz Raiser, Chelsea Steinmetz, Troy Humphries, Samantha Nocita, Brooke Dworak, Carissa Schjebal, Kayla Flege, Avery Dunn, Ashtynn Jarosz, Azra Bauman, Francis Pitts, Jason Powell, Paige Kojdecki, Micki Ortez, Camelia Vidlak, Cody Price, Kayla Downey, Hieu Nguyen, Alan Wilson, Patty Hough, Cortney Roach, Lauren Scrivner, Rylie Herrenbruck, Jessie Mutum | Meridian Clinical Research | Omaha, NE |
| Kenneth Etokhana, MD | Candace Faye, April Reyes, Jeremy Allen, Nathaniel De La Cruz, Tony Diaz, Xavier Fajardo, Veronica Galaviz, Susan Herrera, Meranda Ruiz. | Tekton Research | San Antonio, TX |
| Carlos Fierro, MD | Sam Braijian, Kayla Charles, Fallon Collene, Matthew Davis, Mary Easley, Angela Eichler, Amy Eriksen, Denise Essix, Christa Estrada, Ann Geier, Nicole Godard, Randal Goldstein, Dina Hammine, Kaelyn Howell, Rachel Jackson, Masani Jones, Betsy Kennedy, Eric Lance, Natalia Lesistner, Tish Maun, Mitchel McAfee, Kaley Miller, Kelly Moen, Karol Moor, Lana Morrison, Stephen Naing, Kenny Nguyen, Leslie Owen, Hope Pacheco, Michelle Plumber, Zylyn Richardsen, Maryrohn Rinehart, Latoria Rios, Terri Ruder, Elizabeth Theis, Amy Thompson, Corey Turpin, Amber Wolf, Mazen Zari | Johnson County Clin-Trials | Lenexa, KS |
| Daniel Finn, MD | Ashley Bennett, Nicole Berry, Lindsay Blackmon, Stanley Block, Christal Denton, Lisa Dones, Lisa Good, Kayla Hamilton, Kali Lewis, Shannon Mann, Nicole Mattingly, Wendy Mattingly, Cortney Taylor, Diane Miller, Michelle Newton, Martha Osbourn, Robert Smith, Haley Spalding, Sarah Sparks, Monica Strenecky, Ronald Tyler, Shannon Woods | Kentucky Pediatric/Adult Research | Bardstown, KY |
| Diana F Florescu, MD | Mark Rupp, Daniel Brailita, Adia Sikyta, Erica Stohs, Sara Hurtado Bares, Nada Fadul, Matthew Lunning, Elizabeth Schnaubelt, Molly Ferris, Andrew Buettner, Matthew Palmer, Bailee Lichter, Alison Lewis, Chase Kimberling, Jonathan Beck, Erin Iselin, Kimmai McClain, Andrew Schnaubelt | University of Nebraska Medical Center | Omaha, NE |
| Charles Fogarty, MD | Nicole Crockford, Jami Jones, Anita Morris, Alison Fogarty, Lynne Sarkisian, Angie Williams, Sherry Yeisley, Charlie Fogarty, Connie McCauley | Spartanburg Medical Research | Spartanburg, SC |
| Veronica Garcia-Fragoso, MD | Maria Gabriela Becerra, Lisa Holloway, Bonnie Colville, Shakira Barr, Chen Ho Yang, Tracy Kowalski, Danitra Glasper, Diana Chehab, Joanna Quezon, Maryam Rabbani, Sadaf Batla, Berenice Ferrero, Dean Jang, Enya Rentas-Sherman, Olga Konshina, Teodoro Seminario, Akram Assaf, Frances Saubon, Jenny Torres, Felicia Ardoin, Bernardo Martinez Leal, Faryal Mahmood, Mary Rogers, Vicki Miller, Chance Caddell, Dean Jang, Salma Mustafa, Kathia Hurtado, Syeda Sarwat, Juan Martinez, Alondra Martinez, Sally Hussein, Allison Sorto, Pauline Ngban, Saba Irfan, William Fernandez, Stacy Villareal, Misbah Baloch, Mohammad Rizvi, Shamarrian Hampton, Karina Sainz, Amy Anderson, Ashraf Jafri, Dtuti Sharma, Khola Khan, Kara Sikes, Quiana Wilson, Nina Nuncio, Leticia De La Cruz, Joshua Morre, Imani Troutman, Natasha West, Abdeali Dalal, Hansol Jang, Andrea Juanillo, Omowunmi Fenuyi, Crystal Reese, Dheeraj Narla, Faraya Bokhari, Zurzar Wahaj, Mariya Potapenko, Lucia Almaguer, Muhammad Saleem, Norma Gonzalez, Mirella Melendez, Allie Valentine | Texas Center for Drug Development | Houston, TX |
| George H. Freeman, MD | Esther Laverne Harmon, Marshall A. Cross, Kacie Sales, Catherine Q. Gular, Amanda Fronzaglio, Timothy O’Malley, Zaahin Huq, Jenna Johnson, Jessica Fuggett, Danielle Merian, Rita Quinn | Health Research of Hampton Roads | Newport News, VA |
| David L. Fried, MD | Lynne A. Haughey, Ariana C. Stanton, Jacqueline DiFazio, Filomena Teixeira, Helena L. Godley, Johnna Pezzullo, Monica Freeman, Stephen Gadbois, Emily Anderson | Omega Medical Research | Warwick, RI |
| Cynthia L. Gay, MD, MPH | Maria Bullis, Kirby Caraballo, William Yun Zhao, Kristen Gray, Sarah Law, Jola Mehmeti, Debra Pence, Jane Salm, Allyson Adamo, Catherine Kronk, Alexander Bradley, Miriam Chicurel-Bayard, Nicole Maponga, Julie Nelson, Gloria Oyediran, Charlotte McGehee, Andrew Powell, William Wolf, Ali Castillo, Centhla Washington, Soujanya Anuganti, Kara Barnes, Catherine Pelay, Carmmen Garcia, Paul Alabanza, Kevin Pham, Makayla Nicely, Leah Hahaj, Joseph Trevor Brick, Robin Crawley-Cox, Denise Malone, Amber Barbour, Juan David Chavarriaga, Oesa Vinesett | University of North Carolina | Chapel Hill, NC |
| Carl P. Griffin, MD | Kim Calloway, William Cornelison, Lacey Dietz, Angela Genovese, Shanda Gower, April Green, Brianna Green, Kim Hamilton, Destiny Heingzig-Cartwrtight, Chris Hyatt, Linda Lopez, Ryan Morgan, Idaly Porras, Chalimar Rojo, William Schnitz, Pam Stevenson, Dalia Tovar, Sharee Wright | Lynn Health Science Institute | Oklahoma City, OK |
| Bernard Grunstra, MD | Amy Dye, Shelby Gilmer Olds, Harry Phillip Claybrook, Joshua Bullen, Shai Perry, Miranda Roark, Wendy Hall-Cheers, Kaya Mann, Alasandra Daggs, Jennie Eller, Kayla Mann | PMG Research of Bristol | Bristol, TN |
| Greg Hachigian, MD | Deborah Murray, Michael Cancilla, Logan Ledbetter, Masaru Oshita | Benchmark Research | Sacramento, CA |
| Max Hale, MD | Jill Andringa, Selena Ashworth, Jennifer Campbell, Patrick Farr, Liesel French, Teresa Goldsmith, Karen Martin, Bailey Murphy, Lisa Palmedo, Tracy Sanders, Robert Sellers, James Wamack | Alabama Clinical Therapeutics | Birmingham, AL |
| Charles Harper, MD | Torie Johnson, Courtney Green, Misty Appeldorn, Katlyn Mace, Samantha Wieseler, Chelsea Spilinek, Chelsie Nutsch, Kayla Andal, Kelsey Kelley, Alisha Kiepke, Taysha Hingst, Jennifer Grebe, Wendell Lewis, Sally Eppenbach, Diahn Pekny | Meridian Clinical Research | Norfolk, NE |
| Robyn Hartvickson, MD | Samantha Vanlew, Denae Villines, Justin Phillips, Dylan Thomas, Marissa Mueller, Caressa Presley, Richard Glover II, Amber Grant, Stacy SLechta, Troy Holdeman, Sherry Henning, Colton King, Alyssa Rogers, Jennifer Bennett, Jennifer Richardson, Sidney Ridder, Brooke Dunlavy, Cheryl Sauerwein, Lisa Hemmelgarn, Jana Iniguez, Jared Stremel | Alliance for Multispecialty Research (AMR) | Newton, KS |
| Michael Husseman, MD | Mia Lobato, Ashley MacDonald, Nacy Cleverley, Robert Hoki, James Fennell, Jennifer Gilsoul, Colton Ragsdale, Roderick Maxwell, Bradley Pace, Sherri Wood, Shalyce Schroeder, Tiffani Clayson, Jennifer Wilson, Kimberly Cook, Shelly Searle, Alicia Peterson, Emmy McDaniel, Jenesica Smith, Crystal Amerine, Diane Goodrich, Derek Keeney | Wee Care Pediatrics | Layton, UT |
| Jeffry Jacqmein, MD | Cassie Lawler, Amber DeVries, Cara Seifart, Sonia Gerardo, Cassie Lawler, Angela Morris, Emery Noles, Madison Martinez, Laura Little, Chris Ganzhorn, Brenda Anderson, Amanda Elwood, Ramil Castillo, Lateshia Taylor, Michelle Mackie, Carolina Stewart, Debbie Huxford, Sharon Smith, Erin Schellhorn, Burnadette Moineau, Lisa Joseph, Monica Satorzoda | Jacksonville Center for Clinical Research | Jacksonville, FL |
| Imad Jandali, MD | Katie Leonard, Toni Rich, Abigail Vetter, Maryam Belavilas, David Daniels, Haley Rizzuto | Asclepes Research Centers | Spring Hill, FL |
| Robert Jeanfreau, MD | Nicki Johnson, Estafania Bazan, Shonna James, Katelyn Jackson, RaeShanta McKendall, Matthew Pitts, Melissa Spedale, Brittney Stewart, Susan Tortorich, Steven Darden, Susan Jenafreau, Davilyn Roys, Paul Pati | MedPharmics | Metairie, LA |
| Julie Kasarjian, MD | Judith Kirstein, Sean Harrison, Susan Potter, NP, Erica Coleman, Hanna He, Brenda Delgado, Gladiz Ponce, Brandy Lopez, Nuvia Espinoza, Tonya Gieser, Jazmin Gallegos, Bonnie Goodale, Samantha Carrillo, Nikki Abels, LaTasha Bluder, Sheridan Selby, Katia Talamantez | Velocity Clinical Research | Banning, CA |
| Mark Koch, MD | Norma Escamilla, Lydia Luna, Erin McLeod, Kathryn Dykes, Linda Langdon, Leticia Falcon, Oriella Gonzalez, Anna Marie Parker, Raquel Watkins, Brandon Planchard, Brannon Rolen, Natalie Johnson, Ana Rubio, Jorge Juarez, Chrystal Cook, Roxanne Gomez, Gilda Espinoza, Khadija Arain, Erica Reyes, Jeremiah Sneed, Katie Gaston, Rama Viswatmula, Patricia Bradley, Shalini Raghuraman | Ventavia Research Group | Fort Worth, TX |
| Mark Leibowitz, MD | Fernanda Morales, Rosario Sanchez, Mike Delgado, Norma Vega, Nelly Ayala, Iliana Gallaga, Cassandra Celis, Jennifer Muniz, Mariela Quiroz, Juan Frias, Rea Abaniel, John Nelson, Maricor Grio, Alejandro Moreno, Coralia Soto, Jose Espino, Daniel Vargas, Stephanie Lopez, Angelica Frransico, Eulalia Francisco, Sharon Kelly, Crystal Martinez, Rosie Jimenez, Vivian Garcia, | National Research Institute | Los Angeles, CA |
| Michael Levin, MD | Jessica Corea, Sierra Dansbee, Ashten Davies, Julia Gass, Marcy Kulic, Eduardo Rodreiguz | Clinical Research Center of Nevada | Las Vegas, NV |
| Joseph Ley, MD | Amand Donoho, Ashley Helton, Kimberley Hunt, Anna Johnson, Donald Lewis, Christopher Morelock, David Morin, Lori Ray, Stephanie Tipton, Emily Whitaker | Holston Medical Group | Kingsport, TN |
| Douglas Logan, MD | Amanda Anderson, Michael Anderson, Margaret Baumann, Jennifer Brockhuis, Kelsey Chambers, Natalie Crawford, Lauren Ditmyer-Keith, Tawnyn Ebner, Emily Eichelbrenner, Bruse Gebhardt, Katrina Hale, Gayle Hennekes, Debbie Hilgeman, Victoria Hughes, Tatum Keith, Alexandra Lingel, Douglas Logan, Joanne Major, Brinda Patel, Elizabeth Schaeper, Debra Schroeck, Rhonda Sefton, Jessica Shafer, Samantha Simmons, Ashley Smith, Sarah Walsh, Cindy Wright | Sterling Research Group | Cincinnati, OH |
| Paul G. Matherne, MD | Cassie Beeks, Sarah Bowen, Deven Fejka, Nicole Guttierrez, Lakeyla Bates, Pam Taylor, Gigi Benoit, Jennifer Gates, Jessica Stahl, Lakeyla Bates, Sean Laird, Steven Darden, Tasha Parker, Amber Billiot, Latasha Scott, Amy Caldwell, Amanda Vowell, | Medpharmics | Gulfport, MS |
| Mark McKenzie, MD | Teresa Deese, Vickie Leathers, Misti Earwood, Erica Osmundse, Gisela Heintz, Michelle Forgey, Diane Sproles, Stefanie Mullins, Marquez Obadias, Elizabeth Michael, Shelly Brooks, Angela Harris, Sarah Barrett, Natasha Ballard | WR Clinsearch | Chattanooga, TN |
| Vicki E. Miller, MD, MPH | Amy Star, Blanca Gomez, Monica Murry, Fredric Santiago, Shiela Varghese, Muhammad Irfan, Sonia Guerrero, Madiha Baig, Dustin Watson, Nayab Goher, Maryam Jamil, Heather Leary, Humera Siddiqui, Shelby Danforth, Ragen Powell, Husain Motiwala, Blair Allday, Amy Anderson, Shanika Chatman, Danielle Thompson, Marissa Cruz, Jazmine Spears, Amber Tippit, Md Touhidul Islam, Mirella Melendez, Manisha Joshi, Allie Valentine | DM Clinical Research | Tomball, TX |
| David Musante, MD | William P. Silver, Linda R. Belhorn, Nicholas A. Viens, David Dellaero, Shandelle Parker, Andrew Zimmerman, Roger Ordronneau, Bryan Stanislaus, Megan Dice, Sarah Wilkerson | M3-Emerging Medical Research | Durham, NC |
| Khozema Palanpurwala, MD | Mirza Baig, Lauren Brown, Mercedes Cabrera, Elisa Castro, Esther Cho, Jallesse Flores, Valerie Flores, Vanessa Gonzales, Tania Hernandez, Tetiana Huff, Muhammad Irfan, D’Ambra James, Maria Jeffers, Tenea Johnson, Samjhnana Kafle, Amna Khan, Hermelinda Marroquin, Earl Martin, Rosibel Martinez, Shahad Mohammed, Monica Murray, Thuy Pham, Ragen Powell, Yajaira Rodriguez, Atish Sangahvi, Christina Schmitt, Ambreen Shah, Majid Shah, Amy Starr, Patti Tate, Meghan Tonti, Rebecca Wischnewsky, Aroob Zaheer, | DM Clinical Research - Pediatric Healthcare of NW Houston | Houston, TX |
| Bryce Palchick, MD | Alan Abraham, Nathan Bennett, Sarah Dobrosky, Michael Gates, Suzanne Klutch, Jessica Martier, Laura Pellegrini, Mary Pinyot, Shari Rozen, Kyla Shultz, Victoria Trickett, Jill Waldo | Preferred Primary Care Physicians | Pittsburgh, PA |
| Yogesh K. Paliwal, MD | Amit Paliwal, Krystle Edwards, Sarah Gordon, Connie Navarrete, Alexandria Vasquez, Maria Gomez, Vanessa Monsiviaz, Tony Chi Do, Diana Cardenas, Michael Campos | Empire Clinical Research | Pomona, CA |
| Brannon C Perilloux, MD | Vasavi Srinivasan, Brittany Zedlitz, Stewart Perilloux, Lindsey Lewis, Chris Dedon, Paige Menier, Kelly Castle, Reuben Lilly | Meridian Clinical Research Baton Rouge | Baton Rouge, LA |
| Terry Poling, MD | Terry Klein, Tracy Klein, Sarah Pinkham, Rebecca Andersen, Jennifer McKinzie, Demetria Underwood, Kelly Bailey, Kaley Duu, Kecia Lee, Mariana Parsons, Stephania Lopez, Thomas Klein, Shannen Lassiter, Kaley Dull, Mariana Parsons, Madison Monroe | AMR Wichita East | Wichita, KS |
| Bruce Rankin, DO | John M. Hill, Steven Shinn, Vivek Rajasekhar, Marshall Nash, Michelle Tutt, Kimberlee Del Campo, Douglas F. Winter, Leandro Fernandez, Melissa Hodges, Michelle Jones, Sean Lemoine, Veronica Walker, Roy D. Richardson, Angeline Petracca. Katina Marchione, Michelle Morgan, Ashley McCaffrey, Amber Vasquez, Amy Houck-Dominy, Angela Hammerle, Antonio Rivera, Claxton Copeland, Crystal Paccione, Diana Toney, Fadhel Alyunis, Jennifer Dittman, Kriston Applewhite, Lora Parahovnik, Over Seijas, Ryan Hobbick, Samantha Watts, Shatonia Fields, Stacie Evans, Teresa Logsdon, Thais Truffa, Tifany Huertas, Vienna Bauer,William Serrano, Daisy Sawyer, Giovanni Urquilla, Tonya Toby, Albert Garcia, Alicia J. Cevera,Jeffery Hood, Hannah Hodges, Melissa Willard | Accel Research Sites | DeLand, FL |
| Rosario Retino, MD | Wendy Paiva, Alex Coronel, Florence Harder, Leonillo Pajarillaga, Alex Deniz, Emagacia Reyes, Lendy Torres, Lisa Pyio, | Orange County Research Institute | Ontario, CA |
| Margaret Rhee, MD | Gabrielle Jones, Kelli Meissner, Alanna Billups, Joanne Mortimer, Jane Boggan, Denise Roadman, Celeste Blazy, Melinda DeLong, Cassandra Uminski | Velocity Clinical Cleveland | Cleveland, OH |
| Teresita Salazar, MD | Feminia David, Amiel Guevarra, Noemi Ramirez, Filipinas Vitug | Coast Clinical Research | Bellflower, CA |
| Howard Schwartz, MD | Nelia Sanchez-Crespo, Terry Piedra, Barbara Corral, Jennifer Schwartz | Cenexel RCA | Hollywood, FL |
| William M Seger, MD | John Villegas, Ben Seger, Virginia Loudermilk, Alma Guel, Tisha Davis, Jailyn Reyes, Logan Kimball Crystal Starr, Kathrine Hollie, Ruth Reyes, Elizabeth Boydston, Maria Seger, Tasha Todd, Anthony Kim, Kimberly Pullen, Jean Seignon, Mohammed Antwi, Beverly Ewing | Benchmark Research | Fort Worth, TX |
| Shelly Senders, MD | Christopher Fackelmann, Caitlin Fillioe, Nicholas Jezerinac, Ashley Jeffrey, Emily Horoszoko, Timothy Hudec, Cortney Carr, Alexandra Keresztesy, Emily Gorjanc, | Senders Pediatrics | Cleveland, OH |
| Joanna Sexter, MD | Morgan Cark, Devan Myers, Marian Oshobukola, Elizabeth Weber, | Meridian Clinical Research | Washington, DC |
| Stephan Sharp, MD | Michael Caldwell, Linda Schipani, Allison Ulrich, Wendy Tidwell, Audrey Quickel, Stacy Lynn Cox, Blair Wilkerson, Leanne Martin, Robert Stokes, Lois McClure, Karen Mullen, Ashley Colby, Lynnette Hall, Lisa Booy, Pamela Pinkston, Bailee Saturday | Clinical Research Associates | Nashville, TN |
| Marian E. Shaw, MD | Paula Dockstader, Erika Ayon, Kate Lyon, Patricia Williams, Chris Coon, Ray Coon, Laura Crenshaw, Tracy Moreno, Crislyn Rausch, Erika Mullins, Katie Lacey, Melinda Smith, Jason Calhoun, Tyler Scoggins, Audra Weslowski, Brenda Meza, Shannon Veach, Melanie Wilkerson, Tabitha Scott, Antonio Navarrete, Ronnie Moore, Stacy Martinez, Daniel Korsgaard, Jacqueline Hanson, Nicholas Tuttle | Velocity Clinical Research | Meridian, ID |
| Peter Silas, MD | Jerica Twitchell, Lexie Smith, Jennifer Cooper, Emmy McDaniel, Nancy Cleverley, Cody Hawkes, Brigitte Howard, Tania Peterson, Tiffani Clayson, Jessica Smith, Jennifer Wilson, Kimberly Cook, Shelly Searle, Alicia Peterson, Diane Goodrich, Derek Kenney, Mandi Tanner | Wee Care Pediatrics | Syracuse, UT |
| Teresa S. Sligh, MD | Scott Sligh, Parul Desai, Vincent Huynh, Carlos Lopez, Erika Mendoza, Dennis Perez, Samuel Ceballos, Jennifer Gomez, Janneth Becerra, Tiffany Martinez, Erika Navarro Fausto | Providence Clinical Research | North Hollywood, CA |
| Kenneth Steil, MD | Mandi Ward, Scarlett Hammett, Alyssa McAramara, Maria Gustilo, Carey Goldsmith, Amy Brace, Mildred DeJesus, Jaimi Jones, Desiree Tobin, Laronda Lee, Brian Martin, Andrew Wall | Wee Care Pediatrics | Syracuse, UT |
| Cayce Tangeman, MD | Cayce Tangeman, Yvonne Davis, Rica Santiago, George Minson, John Bowman, Vanessa Armetta, Tiffany Hill, David Summers, Emily Caudle, Mary Love, Jessica Brindel, Mark Henriksen, Taylor McCormick, Wesley Stubbs, Melba Freedy, Tara Kelly, Susan English, Gino Pineda, Derrick Brown, Patrick Greene, Genda Williams, Theresa Highsmith, Patrik Das | Coastal Carolina Research Center | North Charleston, SC |
| Christine B. Turley, MD | Lewis McCurdy, Tonisha Brown, Martha Pawlicki, Jennifer Reeves, Jona Bauer, Cedrick Griner, Cameron Russell, Veena Sampathkumar, Zeynep Alimchandani, Robin Muller, Tracey Coakley, Mary Sours, Saifelnasr Mohamed, Sone Alanoh, Amy Yeh, Sahra Khan, Eleojo Abutu, Genena Buck, Sarah Hicks, Andreana Alexander, Tammy Patterson, Maria Martilnsalaco, Amy Clontz, Marina Leonidas, Zainab Shahid, Jay I. Patel, Ryan Bender | The Charlotte-Mecklenburg Hospital Authority | Charlotte, NC |
| Lisa S. Usdan, MD | LaKeshia Pipkin, Vanessa Wheeler, Julia Sinatra, Irene Woods Powell, Leslie Lazar, Debra O’Brien, Kelly Iskiwitz, Carol Marsh, Tavia Flagg, Erin Wells, Brandi Gruber, Lora J. McGill, Valerie K. Arnold, Carolyn J. Scatamacchia, Codi M. Anthony, Robyn M. Presley, Melissa N. Flowers, Charles L. Grandberry, Cathy T. Houpt, Dominique L.Ross | Clinical Neuroscience Solutions | Memphis, TN |
| Larkin Tyler Wadsworth III, MD | Horacio Marafioti, Lyly Dang, Lauren Clement, Kristen, Anya Penly, Elizabeth Garner, Angie Kean, Sophia Bolakas, Cerece Miles, Christy Shultz, George Cherniawski, Stephanie Tesson, Breanna Galibert, Karen Knapp, Jennifer Berry, Alexander Chistensen, Monica Matkins | Sundance Clinical Research | St. Louis, MO |
| Michael Waters, MD | Dalia Tover, Scott Overcash, Jordan Coslet, Michael Voskanian, Giuliano Zolin, Matthew Petro, Gina Weaver, Kia Lee, Hanh Chu, Karla Zepeda, Crystle Rajania, John Rodriguez, Tracey Fabrega, Kaitlyn Sandler, Alex Tapia, Cecilia Barbabosa, Renee Pasion, Jacob Pineda, Rosalynn Landazuri, Angelica Franco, Estee Garcia, Marilynn Rodriguez, Joanna Ocampo | Velocity Clinical Research | Chula Vista, CA |
| Paul Wisman, MD | Carlos Amengol, Gemila Bouber, Peggy Bressler, Alaina Brown, Josh Bruce, Candyce Dorsey, Maire Lee, MIcehlle Sarah Knight, Amy Malek, Shana Murphy, Adriane Neumeister, L. Page Perriello, Marie Vetter, Kelly Vincel, Claudia Wisman | Pediatric Research of Charlottesville | Charlottesville, VA |

**Data and Safety Monitoring Board (DSMB) Members List**

| **Member** | **Institution** |
| --- | --- |
| Richard J. Whitley, MD (Chair) | School of Medicine, University of Alabama at Birmingham |
| Abdel Babiker, PhD | MRC Clinical Trials Unit at University College, London |
| Lisa A. Cooper, MD, MPH | Johns Hopkins University School of Medicine and Bloomberg School of Public Health |
| Susan S. Ellenberg, PhD | University of Pennsylvania School of Medicine,  Center for Clinical Epidemiology and Biostatistics,  Division of Biostatistics |
| Alan Fix, MD, MS | Vaccine Development Global Program, Center for Vaccine Innovation and Access, PATH |
| Marie Griffin, MD, MPH | Vanderbilt University Medical Center |
| Steven Joffe, MD, MPH | University of Pennsylvania, Perelman School of Medicine |
| Jorge Kalil, MD, MPH | Heart Institute, Hospital das Clínicas da Faculdade de Medicina da Universidade de São Paulo, Brazil |
| Myron M. Levine, MD, DTPH | University of Maryland School of Medicine |
| Malegapuru W. Makgoba, MB, ChB, DPhil, FRCP | University of KwaZulu-Natal, South Africa |
| Reneé H. Moore, PhD | Drexel University |
| Anastasios A. Tsiatis, PhD | North Carolina State University |
| Sally Hunsberger, PhD (Executive Secretary) | NIAID, NIH |

**Objectives and End Points.**

**Table S1. Primary and Secondary Objectives and End Points Addressed in This Manuscript. Protocol Version 8.0.**

| Objectives | End Points |
| --- | --- |
| **Primary:** | **Primary:** |
| - To evaluate the efficacy of a 2-dose regimen of SARS-CoV-2 rS adjuvanted with Matrix-M1 compared to placebo against PCR-confirmed symptomatic COVID-19 illness diagnosed ≥ 7 days after completion of the second injection in the initial set of vaccinations of adolescent participants 12 to < 18 years of age. - To describe the safety experience for the vaccine versus placebo in adolescent participants (12 to < 18 years of age) based on solicited short-term reactogenicity by toxicity grade for 7 days following each vaccination (Days 0 and 21) after the initial set of vaccinations. - To assess overall safety through 49 days (28 days after second injection of each set of vaccinations [initial and crossover]) by comparing vaccine versus placebo for all unsolicited AEs and MAAEs. - To assess the frequency and severity of MAAEs attributed to vaccine, AESIs, or SAEs through the EoS and to compare vaccine versus placebo after each set of vaccinations (initial and crossover). - To assess all-cause mortality in vaccine versus placebo recipients after each set of vaccinations (initial and crossover). - To assess non-inferiority of the neutralizing antibody response for all adolescent participants seronegative to anti-SARS-CoV-2 NP antibodies at baseline, compared with that observed in seronegative adult participants 18 to 25 years of age from the Adult Main Study (Immunogenicity Population participants before crossover). | **Primary Endpoints:**   - First episode of PCR-positive mild, moderate, or severe COVID-19, where severity is defined as:   **Mild COVID-19 (≥ 1 of the following):**   - Fever (defined by subjective or objective measure, regardless of use of anti‑pyretic medications) - New onset cough - ≥ 2 additional COVID-19 symptoms:   - New onset or worsening of shortness of breath or difficulty breathing compared to baseline.   - New onset fatigue.   - New onset generalized muscle or body aches.   - New onset headache.   - New loss of taste or smell.   - Acute onset of sore throat, congestion or runny nose.   - New onset nausea, vomiting or diarrhea.   **OR Moderate COVID-19 (≥ 1 of the following):**   - High fever (≥ 38.4°C) for ≥ 3 days (regardless of use of anti-pyretic medications, need not be contiguous days). - Any evidence of significant LRTI:   - Shortness of breath (or breathlessness or difficulty breathing) with or without exertion (greater than baseline).   - Tachypnea: 24 to 29 breaths per minute at rest.   - SpO_2_: 94% to 95% on room air.   - Abnormal chest X-ray or chest CT consistent with pneumonia or LRTI. - Adventitious sounds on lung auscultation (eg, crackles/rales, wheeze, rhonchi, pleural rub, stridor).   **OR Severe COVID-19 (≥ 1 of the following):**   - Tachypnea: ≥ 30 breaths per minute at rest. - Resting heart rate ≥ 125 beats per minute. - SpO_2_: ≤ 93% on room air or PaO_2_/FiO_2_ < 300 mmHg. - High flow oxygen (O_2_) therapy or NIV/NIPPV (eg, CPAP or BiPAP). - Mechanical ventilation or ECMO. - One or more major organ system dysfunction or failure to be defined by diagnostic testing/clinical syndrome/interventions, including any of the following:   - Acute respiratory failure, including ARDS.   - Acute renal failure.   - Acute hepatic failure.   - Acute right or left heart failure.   - Septic or cardiogenic shock (with shock defined as SBP < 90 mm Hg OR DBP < 60 mm Hg).   - Acute stroke (ischemic or hemorrhagic).   - Acute thrombotic event: AMI, DVT, PE.   - Requirement for: vasopressors, systemic corticosteroids, or hemodialysis. - MIS-C, as per the CDC definition:   - An individual aged < 21 years presenting with fever (>38.0°C for ≥24 hours, or report of subjective fever lasting ≥24 hours), laboratory evidence of inflammation (including, but not limited to, one or more of the following: an elevated C-reactive protein (CRP), erythrocyte sedimentation rate (ESR), fibrinogen, procalcitonin, d-dimer, ferritin, lactic acid dehydrogenase (LDH), or interleukin 6 (IL-6), elevated neutrophils, reduced lymphocytes and low albumin), and evidence of clinically severe illness requiring hospitalization, with multisystem (>2) organ involvement (cardiac, renal, respiratory, hematologic, gastrointestinal, dermatologic or neurological); AND   - No alternative plausible diagnoses; AND   - Positive for current or recent SARS-CoV-2 infection by RT-PCR, serology, or antigen test; or COVID-19 exposure within the 4 weeks prior to the onset of symptoms. - Admission to an ICU. - Death   **Safety Endpoints:**   - Reactogenicity incidence, duration and severity (mild, moderate or severe) recorded by parent(s)/caregiver(s) on their electronic patient-reported outcome diary application (eDiary) on days of vaccination and subsequent 6 days (total 7 days after each vaccine injection in the initial set of vaccinations). - Reactogenicity endpoints include injection site reactions: - Pain. - Tenderness. - Erythema. - Swelling/induration. - Systemic reactions: - Fever. - Malaise. - Fatigue. - Arthralgia. - Myalgia. - Headache. - Nausea/vomiting. - Incidence and severity of MAAEs through 49 days, ie, 28 days after second injection of each set of vaccinations (initial and crossover). - Incidence and severity of unsolicited AEs through 49 days, ie, 28 days after second injection of each set of vaccinations (initial and crossover). - Incidence and severity of MAAEs attributed to study vaccine, SAEs and AESIs through Month 12. COVID-19 diagnoses will be included as “medically important” events that are reported as SAEs. - Incidence and severity of SAEs (including COVID-19 diagnoses), MAAEs attributed to study vaccine and AESIs during Month 12 through Month 24 or the EoS. - Death due to any cause.   **Effectiveness Endpoint:**   - Neutralizing antibody response at Day 35 for all adolescent participants seronegative to anti-SARS-CoV-2 NP antibodies at baseline, compared with that observed in seronegative adult participants 18 to 25 years of age from the **Adult Main Study** (Immunogenicity Population participants before crossover). |
| **Secondary Objectives:** | **Secondary End Points:** |
| - To evaluate the efficacy of a two-dose regimen of SARS-CoV-2 rS adjuvanted with Matrix-M™ compared to placebo against RT-PCR-confirmed moderate-to-severely symptomatic Covid-19 illness diagnosed ≥7 days after completion of the second vaccination in the initial set of vaccinations of adult participants ≥18 years of age. - To assess VE against ANY symptomatic SARS‑CoV-2 infection. - To assess VE according to race and ethnicity. | - First episode of RT-PCR-positive moderate or severe Covid-19, as defined under the primary end point. - ANY symptomatic SARS-CoV-2 infection, defined as: RT-PCR-positive nasal swab and ≥1 of any of the following symptoms: - Fever. - New onset cough. - New onset or worsening of shortness of breath or difficulty breathing compared to baseline. - New onset fatigue. - New onset generalized muscle or body aches. - New onset headache. - New loss of taste or smell. - Acute onset of sore throat, congestion, or runny nose. - New onset nausea, vomiting, or diarrhea. - Description of course, treatment, and severity of Covid‑19 reported after a RT-PCR-confirmed case via the Endpoint Form. |

Abbreviations: AESI = adverse event of special interest; AMI = acute myocardial infarction; ARDS = acute respiratory distress syndrome; BiPAP = bilevel positive airway pressure; BMI = body mass index; CDC = Centers for Disease Control and Prevention; Covid-19 = coronavirus disease 2019; CoVPN = Covid-19 Prevention Network; CPAP = continuous positive airway pressure; CT = computed tomography; DBP = diastolic blood pressure; DVT = deep vein thrombosis; ECMO = extracorporeal membrane oxygenation; eDiary = electronic patient-reported outcome diary; EoS = end of study; FiO_2_ = fraction of inspired oxygen; hACE2 = human angiotensin-converting enzyme 2; ICU = intensive care unit; IgG = immunoglobulin G; IFN-γ = interferon gamma; IL = interleukin; LRTI = lower respiratory tract infection; MAAE = medically attended adverse event; MN = microneutralization; NIAID = National Institute of Allergy and Infectious Diseases; NIH = National Institutes of Health; NP = nucleocapsid; NIPPV = non-invasive positive pressure ventilation; NIV = non-invasive ventilation; O_2_ = oxygen; OWS = Operation Warp Speed; PaO_2_ = partial pressure of oxygen; PBMC = peripheral blood mononuclear cell; RT-PCR = reverse transcriptase-polymerase chain reaction; PE = pulmonary embolism; SAE = serious adverse event; SARS-CoV-2 = severe acute respiratory syndrome coronavirus 2; SARS-CoV-2 rS = severe acute respiratory syndrome coronavirus 2 recombinant spike protein nanoparticle vaccine; SBP = systolic blood pressure; SpO_2_, oxygen saturation; Th1 = type 1 T helper; Th2 = type 2 T helper; TNF-α = tumor necrosis factor alpha; VE = vaccine efficacy.

**Supplemental Methods**

**Safety Assessments**

Following collection of sufficient safety data to support application for Emergency Use Authorization (EUA), i.e., median 2 months’ duration of safety follow-up, participants were offered administration of two injections of the alternate study material 21 days apart (“blinded crossover”). That is, initial recipients of placebo did receive SARS-CoV-2 rS with Matrix-M™ adjuvant and initial recipients of SARS-CoV-2 rS with Matrix-M™ adjuvant did receive placebo. The same procedure used for the initial set of vaccinations was followed at the time of the blinded crossover to ensure that the integrity of the blinded study was maintained.

Solicited AEs of reactogenicity after the initial series of vaccinations was collected via participant’s parents/representative reporting in the eDiary utilizing a smartphone application. All participants’ parents/representative were trained on the use of these applications, and smartphone devices were provided for those parents/representatives who needed them at the initiation of their participation in the study (Day 0).

Safety assessments included collection of participant-recorded solicited (local and systemic reactogenicity) events through 7 days following each injection in the initial set of vaccinations collected via eDiary. Unsolicited AEs and MAAEs were collected through 49 days, i.e., 28 days after second injection of the initial and crossover sets of vaccinations. MAAEs attributed to vaccine, AESIs, SAEs, will be collected through EoS.

**Safety Monitoring**

Safety was monitored routinely by the Sponsor physicians. A centralized Data and Safety Monitoring Board (DSMB) was established in collaboration with NIH, NIAID, Biomedical Advanced Research and Development Authority (BARDA), and Novavax according to the charter dictated by the participating groups. This group reviewed interim unblinded data periodically throughout the period of blinded follow-up and made recommendations with respect to safety.

**Prospective Surveillance of Covid-19**

For prospective surveillance, participants’ parents were requested to notify clinical sites as soon as fever or other specified symptoms were experienced by the participant or during the scheduled weekly remote contacts with the site. Sites then scheduled an in-person Acute Illness Visit for medical evaluation (to include oxygen [O_2_] saturation and respiratory rate) and medically attended nasal swab in symptomatic participants. Active surveillance for Covid-19 continued after the blinded crossover through the first 12 months of study. Passive surveillance of safety and efficacy via remote contacts or the scheduled visits continues during Months 12 to 24.

Study participants whose medically attended nasal swabs from the Acute Illness Visit were confirmed at the central laboratory to be RT-PCR-positive for SARS-CoV-2 were contacted by the study site to arrange a Convalescent Visit. The Convalescent Visit was to occur approximately 1 month (or as soon thereafter, as feasible) after the onset of the RT-PCR–confirmed case of Covid-19 to assess status of AEs, record the clinical course of the disease on the End Point Form, and obtain a blood sample for convalescent serologic testing.

**Covid-19 End Point Assessment**

To ensure the quality and accuracy of investigator-recorded end point assessments collected on the Endpoint Assessment electronic case report form (eCRF) page, programmatic checking was performed prior to the data extraction for analysis. The algorithms and the data sources to be used for programming were determined and documented prior to unblinding. Using data elements relevant to the end point definition and captured in the eCRF, programmatic determination of potential end points and associated start date and severity were performed. Data elements used included RT-PCR results by the central laboratory from the swab and pulse oximeter readings collected at the Acute Illness Visit. Disease episodes were constructed programmatically, including date of initial symptoms, date of positive RT-PCR test result, and preliminary severity based on symptoms reported and pulse oximeter readings. The programmatically determined end points were compared with the data collected on the Endpoint Assessment eCRF. Discrepancies such as missing or difference in date of start of illness or difference in severity rating prompted Data Management to issue queries to the investigators. The Endpoint Assessment eCRF data, along with the official study RT-PCR results from the University of Washington, Seattle, WA, were used for analysis of the efficacy end points.

Potentially severe cases of symptomatic RT-PCR–positive Covid-19 were reviewed by an external Independent Endpoint Review Committee (ERC) established by the sponsor. The ERC consisted of physicians with clinical and research experience (e.g., medical review and/or clinical trial experience) in infectious diseases. The committee’s structure, responsibilities, and operation were specified within a charter. Potentially severe cases included Covid-19 reported as SAEs, programmatically identified end points consisting of at least 1 pulse oximeter reading ≤93%, and episodes identified as severe on the Endpoint Assessment eCRF. Participant profiles (as outlined in the charter) were provided to the committee members for review according to the process outlined in the charter. These participant profiles included demographics, medical history, AEs, concomitant medications, and the Endpoint Assessment eCRF. The external reviewers documented the criteria used for their clinical review of the cases.

The results of the review were to confirm the case as severe or rule that the case was not severe. The outcome of the review for each case was stored in an electronic medical review system. A file was exported from the system and provided to the Biostatistics group for use in analysis. Cases that were ruled as not severe by the committee despite an investigator-entered severe grading were further reviewed and documented by Novavax clinician(s) prior to unblinding to determine the severity to use in analysis. Cases that were ruled as severe by the ERC but not severe by the investigator were analyzed as severe in the analysis.

**Nasal Swabs for Viral Detection**

Swabs of the anterior nares were obtained at the study site on Day 0 (prior to study vaccination), at the Acute Illness Visit, and at the first crossover vaccination visit.

Quantitative RT-PCR was performed on RT-PCR–positive swabs using the Abbott RealTime RT-PCR to assess viral load and sequencing of viral genetic material detected in nasal swab RT-PCR testing to evaluate viral mutations.

**SARS-CoV-2 RT-PCR Testing**

The RT-PCR test being used is the Abbott RealTime SARS-CoV-2 Assay, which was granted EUA by the US Food and Drug Administration (FDA) on March 18, 2020 (<https://www.fda.gov/media/136255/download>). The testing was performed at the University of Washington. The validation and verification of the Abbott RealTime SARS-CoV-2 Assay analytical and clinical performance has been published.^1^ Dry swabs were used and have been validated for this assay with storage at 2-8°C for up to 7 days and then frozen at -80°C. Once the sample is received at the University of Washington, the dry swabs are eluted into Roswell Park Memorial Institute (RPMI) +2% fetal bovine serum (FBS) or phosphate buffered saline (PBS) prior to testing. The University of Washington conducted an analysis validating and verifying the performance of the Abbott RealTime SARS-CoV-2 Assay with varying viral dilutions. The outcome of the analysis was published elsewhere.^2^

**Whole-Genome Sequencing (WGS) and Clade/Lineage Assignment**

Secondary end points of the trial included the first episode of RT-PCR–positive, symptomatic Covid-19 due to strain shown by gene sequencing to represent a variant not considered as a variant of concern (VOC) or variant of interest (VOI), as well as those due to VOC/VOI, according to the Centers for Disease Control and Prevention (CDC) SARS-CoV-2 Variant Classifications and Definitions,^3^ starting at least 7 days post-vaccination 2 in the initial vaccination period. However, due to the SARS-CoV-2 epidemiology at time of initiation of the Pediatric Expansion of PREVENT-19, there were no cases reported due to variants antigenically comparable to the prototype strain. In fact, all sequences from Covid-19 cases were identified as the Delta variant.

Baseline RT-PCR–positive samples as well as RT-PCR positive samples from the Acute Illness Visit with enough viral load were sent to the University of Washington Virology Laboratory for WGS, using methodology described elsewhere.^4,5^ In case of multiple RT-PCR-positive samples for a given symptomatic episode, that with the highest viral load was chosen for sequencing.

Sequencing analysis included viral clade/lineage assignment using both the Nextstrain clade label system (<https://nextstrain.org/blog/2021-01-06-updated-SARS-CoV-2-clade-naming>) and the PANGO lineages designation system (<https://cov-lineages.org/>), and identification as a VOC/VOI/High Consequence, per the CDC (<https://www.cdc.gov/coronavirus/2019-ncov/variants/variant-info.html>). Evaluation of SARS-CoV-2 infection or disease was available by site and/or geographic region. The classification of variants was conducted by the University of Washington Virology Laboratory and provided for analysis.

**Anti-S IgG Antibody Titer Determination (ELISA) [Fit-for-Purpose Assay] (Figure S4)**

The quantitation of anti-spike (S) protein immunoglobulin G (IgG) by enzyme-linked immunosorbent assay (ELISA) (EC_50_) against recently emerging variants was conducted in the Novavax Vaccine Immunology Laboratory to determine cross-reactivity in *post-hoc* testing. This procedure was fit-for-purpose and differs minimally from the validated version of the assay performed by Novavax Clinical Immunology Laboratory for the SARS-CoV-2 Wuhan prototype strain that was used for per protocol testing. The validated assay has been already described elsewhere.^6^ The validated method uses a reference standard to measure the total IgG in the serum samples to ensure the consistent measurement and the IgG is reported in ELISA units (EU/mL). Both methods are technically similar, except for the final readout.

For the fit-for-purpose assay, 96-well microtiter plates (Thermo Fisher Scientific, Rochester, NY, USA) were coated with 1.0 µg/mL of SARS-CoV-2 rS protein from Wuhan-Hu-1, Alpha, Beta, Delta, Delta Plus, Gamma, Mu, and Omicron BA.1. Plates were washed with PBS-T and nonspecific binding was blocked with TBS Startblock blocking buffer (Thermo Fisher Scientific). Serum samples were serially diluted 3-fold down starting with a 1:300 dilution (ie, 10^-2^ to 10^-8^) and added to the coated plates followed by incubation at room temperature for 2 hours. Following incubation, plates were washed with PBS-T and HRP-conjugated goat anti-human IgG (Southern Biotech, Birmingham, AL, USA) was added for 1 hour. Plates were washed with PBS-T and 3,3’,5,5’-tetramethylbenzidine (TMB) peroxidase substrate (Sigma, St Louis, MO, USA) was added. Reactions were stopped with TMB stop solution (ScyTek Laboratories, Inc. Logan, UT). Plates were read at optical density (OD) 450 nm with a SpectraMax Plus plate reader (Molecular Devices, Sunnyvale, CA, USA). EC_50_ titer values were calculated by 4-parameter fitting using SoftMax Pro 6.5.1 GxP software. Individual subject anti-S IgG titers, group geometric mean titers, and 95% confidence intervals (CIs) were plotted using GraphPad Prism 7.05 software. For serum titers below the assay lower limit of detection (LOD), a titer of < 300 (starting dilution) was reported and a value of “150” assigned to the sample to calculate the group mean titer.

**hACE2 Receptor Inhibiting Antibody Titer Determination (ELISA) [Fit-for-Purpose Assay] (Figure S5)**

Human ACE2 receptor blocking antibody ELISA against newer variants was performed in the Novavax Vaccine Immunology Laboratories as *post-hoc* testing. This procedure was fit-for-purpose and differs minimally technically from the validated version of the SARS-CoV-2 Wuhan prototype strain assay performed by Novavax Clinical Immunology Laboratory. The validated assay has been already described elsewhere.^7^

For the fit-for-purpose assay, ninety-six-well plates were coated with 1.0 μg/mL SARS-CoV-2 rS protein from the Wuhan-Hu-1, Alpha, Beta, Delta, Delta Plus, Gamma, Mu, and Omicron BA.1 variants, overnight at 4°C. Plates were washed with PBS-T, and nonspecific binding was blocked with TBS Startblock blocking buffer (Thermo Fisher Scientific). Sera were serially diluted 2-fold starting with a 1:20 dilution and added to coated wells for 1 hour at room temperature. After washing, 30 ng/mL of histidine-tagged hACE2 (Sino Biologicals, Beijing, CN) was added to wells for 1 hour at room temperature. HRP-conjugated anti-histidine IgG was added and incubated for 1 hour followed by addition of TMB substrate. Plates were read at OD 450 nm with a SpectraMax Plus plate reader (Molecular Devices, Sunnyvale, CA, USA) and data analyzed with SoftMax Pro software. The % Inhibition for each dilution for each sample was calculated using the following equation in the SoftMax Pro program: 100-[(MeanResults/ControlValue@PositiveControl)*100].

Serum dilution versus %Inhibition plot was generated, and curve fitting was done by 4 parameter logistic (4PL) curve fitting to data. Serum antibody titer at 50% binding inhibition (IC_50_) of hACE2 to SARS-CoV-2 rS protein was determined in the SoftMax Pro program. Individual subject hACE2 receptor inhibiting titers, group geometric mean titers, and 95% CI were plotted using GraphPad Prism 7.05 software. For a titer below the assay LOD, a titer of <20 (starting dilution) was reported and a value of “10” assigned to the sample to calculate the group mean titer.

**Analysis Populations**

There were 6 main analysis sets used in this trial:

- The **Intent-to-Treat (ITT) Analysis Set** included all participants who were randomized, regardless of protocol violations or missing data. The ITT analysis set was used for participant disposition summaries and were analyzed according to the treatment arm to which the participant was randomized.
- The **Full Analysis Set (FAS)** included all participants who were randomized and received at least one dose of study vaccine/placebo, regardless of protocol violations or missing data. Participants who were unblinded with an intention to receive other Covid-19 vaccines were censored at the time of unblinding. The FAS population was analyzed according to the treatment group to which participants were randomized. The FAS were used for supportive analyses. When the efficacy end points were analyzed using FAS, baseline SARS-CoV-2 seropositivity or nasal swab RT-PCR-positivity was ignored.
- The **Safety Analysis Set** included all randomized participants who received at least one dose of study vaccine/placebo. Participants in the Safety Analysis Set were analyzed according to the treatment actually received. In cases where information is available that indicated that a participant received both active vaccine and placebo during the initial vaccination series, the participant was analyzed as part of the active group.
- The **Per-Protocol Efficacy (PP-EFF) Analysis Set** included all participants who received the full prescribed regimen of trial vaccine and had no major protocol deviations that occurred before the first Covid-19 positive episode (i.e., participant was censored at the time of the protocol deviation) and were determined to affect the efficacy outcomes, including baseline SARS-CoV-2 seropositivity or nasal swab RT-PCR‑positivity. Participants who were unblinded with an intention to receive other Covid-19 vaccines were censored at the time of unblinding. Although the study enrolled participants regardless of SARS-CoV-2 serologic status at the time of initial vaccination, any participants with confirmed infection or prior infection due to SARS‑CoV-2 at baseline, by nasal swab RT-PCR or serology (assessed by anti-nucleocapsid [anti-NP]), were excluded from the PP‑EFF population. PP-EFF was the primary set for all efficacy end points.
- A second **PP-EFF (PP-EFF-2) Analysis Set** was defined to allow evaluation of baseline serostatus analysis’s impact on vaccine efficacy (VE). The PP-EFF-2 Analysis Set followed the same method described in the PP-EFF population, with the exception that it included all participants regardless of baseline serostatus (anti-NP serology) or baseline virological status (RT-PCR).
- The Day 35 **Per-Protocol Immunogenicity (PP-IMM) Analysis Set** included all participants who had at least a baseline and Day 35 sample result available after vaccination and had no major protocol deviations that occurred before the Day 35 visit (i.e., participant was censored at the time of the protocol deviation) that were determined to affect the effectiveness outcomes, including baseline SARS-CoV-2 seropositivity or nasal swab RT-PCR positivity. Participants who were unblinded with an intention to receive other Covid-19 vaccines were censored at the time of unblinding. Although the study enrolled participants regardless of SARS-CoV-2 serologic status at the time of initial vaccination, any participants with confirmed infection or prior infection due to SARSCoV-2 at baseline, by nasal swab RT-PCR or serology (assessed by anti-NP), were excluded from the PP-IMM population. Participants must have received the second vaccination to be included in the PP-IMM analysis set. PP-IMM was the primary set for all effectiveness end points.

**Statistical Method for Efficacy End Points**

The VE is defined as VE (%) = (1 – RR) × 100, where RR = relative risk of incidence rates between the two trial vaccine groups (SARS-CoV-2 rS / placebo). The RR was estimated by exponentiating the treatment group coefficient from a Poisson regression analysis with robust error variance.^8^ To assess incidence rates rather than absolute counts of cases, accounting for differences in follow-up times starting at 7 days after the second vaccination among participants, an offset was utilized in the Poisson regression. The generalized linear model with unstructured correlation matrix (robust error variances) was used. The explanatory variables in the model included the trial vaccine group. The dependent variable was the incidence rate of the end point of interest. The robust error variances were estimated using repeated statement and the participant identifier. To account for the censoring in the analysis, the offset was defined as the natural log of the time from the start of follow-up (7 days post-second vaccination) to the outcome of interest or to the end of study in addition to censoring described in the analysis set definitions. Poisson distribution was used with a logarithmic link function. In the case where there were zero end points for one of the vaccine groups or the total number of end points in both treatment groups combined is less than 5, a Poisson model was substituted with an exact conditional binomial method using the Clopper-Pearson method. This method conditions on the total number of events across the treatment groups where the number of events in the active group are generated from a single binomial distribution. The point estimate from this single binomial distribution and the corresponding confidence intervals constructed using the Clopper-Pearson method were transformed back to relative risks. A Cox proportional hazards model was also developed as a supportive analysis to the Poisson regression. The model followed the same explanatory and dependent variables as the Poisson model and censored participants based on their follow-up time available.

**Statistical Method for Effectiveness**

For the effectiveness endpoint (non-inferiority of the neutralizing antibody response compared with young adults from the PREVENT-19 trial, 18 to <26 years), successful demonstration of non-inferiority requires meeting the following 3 pre-specified criteria simultaneously:

1. lower bound of 2-sided 95% CI for the ratio of geometric mean titers (GMTs) (GMT_12-<18yo_/GMT_18-<26yo_) > 0.67
2. point estimate of the ratio of GMTs ≥ 0.82 (estimated as square root of 2/3)
3. lower bound of the 2-sided 95% CI for difference of serologic response (SCR_12-<18yo_/SRC_18-<26yo_) > - 10%

With 400 evaluable participants (500, accounting for 20% non-evaluability) in the active vaccine group randomly selected from each of the 18 to < 26 years of age subset of participants in the Adult Main Study and the Pediatric Expansion of PREVENT-19, there is over 85% power (through simulations) to demonstrate the first two non-inferiority criteria when assuming an underlying GMT for the 18 to <26 years of age group up to 1.1-fold higher than the 12 to <18 years of age group. In the absence of an established correlate of protection for SARS-CoV-2 vaccines, serologic response was defined as ≥ 4-fold increase in neutralization titers (MN50) at Day 35 relative to baseline titers. With this definition and assumed SCR of 95% in the 18 to <26 years of age group, there is over 80% power to demonstrate the third non-inferiority criterion for a difference as large as 4% lower in the 12 to <18 years of age group.

**Supplemental Tables and Figures**

**Table S2. Symptoms Suggestive of Covid-19**

- Fever (body temperature >38.0°C, in the absence of other symptoms) or chills

- New onset or worsening of cough compared with baseline

- New onset or worsening of shortness of breath or difficulty breathing over baseline

- New onset of fatigue

- New onset of generalized muscle or body aches

- New onset of headache

- New loss of taste or smell

- Acute onset of sore throat

- Acute onset of congestion or runny nose

- New onset of nausea or vomiting

- New onset of diarrhea

Abbreviations: Covid-19 = coronavirus disease 2019.

**Table S3. End Point Definitions of Covid-19 Severity**

| **Covid-19 Severity** | **End Point Definitions** |
| --- | --- |
|  | **First episode of RT-PCR-positive mild, moderate, or severe Covid-19:** |
| **Mild** | **≥1 of the following:**   - Fever (defined by subjective or objective measure, regardless of use of anti‑pyretic medications) - New onset of cough - ≥2 additional Covid-19 symptoms:   - New onset or worsening of shortness of breath or difficulty breathing compared to baseline.   - New onset of fatigue.   - New onset of generalized muscle or body aches.   - New onset of headache.   - New loss of taste or smell.   - Acute onset of sore throat, congestion, or runny nose.   - New onset of nausea, vomiting, or diarrhea. |
| **Moderate*** | **≥1 of the following:**   - High fever (≥38.4°C) for ≥3 days (regardless of use of anti-pyretic medications, need not be contiguous days). - Any evidence of significant lower respiratory tract infection (LRTI):   - Shortness of breath (or breathlessness or difficulty breathing) with or without exertion (greater than baseline).   - Tachypnea: 24 to 29 breaths per minute at rest.   - SpO_2_: 94% to 95% on room air.   - Abnormal chest X-ray or chest computerized tomography (CT) consistent with pneumonia or LRTI. - Adventitious sounds on lung auscultation (e.g., crackles/rales, wheeze, rhonchi, pleural rub, stridor). |
| **Severe*** | **≥1 of the following:**   - Tachypnea: ≥30 breaths per minute at rest. - Resting heart rate ≥125 beats per minute. - SpO_2_: ≤93% on room air or PaO_2_/FiO_2_ <300 mmHg. - High flow oxygen (O_2_) therapy or non-invasive ventilation (NIV)/non-invasive positive pressure ventilation (NIPPV) (e.g., continuous positive airway pressure [CPAP] or bilevel positive airway pressure [BiPAP]). - Mechanical ventilation or extracorporeal membrane oxygenation (ECMO). - One or more major organ system dysfunction or failure to be defined by diagnostic testing/clinical syndrome/interventions, including any of the following:   - Acute respiratory failure, including acute respiratory distress syndrome (ARDS).   - Acute renal failure.   - Acute hepatic failure.   - Acute right or left heart failure.   - Septic or cardiogenic shock (with shock defined as systolic blood pressure [SBP] <90 mm Hg OR diastolic blood pressure [DBP] <60 mm Hg).   - Acute stroke (ischemic or hemorrhagic).   - Acute thrombotic event: acute myocardial infarction (AMI), deep vein thrombosis (DVT), pulmonary embolism (PE).   - Requirement for: vasopressors, systemic corticosteroids, or hemodialysis. - MIS-C, as per the CDC definition:   - An individual aged < 21 years presenting with fever (>38.0°C for ≥24 hours, or report of subjective fever lasting ≥24 hours), laboratory evidence of inflammation (including, but not limited to, one or more of the following: an elevated C-reactive protein (CRP), erythrocyte sedimentation rate (ESR), fibrinogen, procalcitonin, d-dimer, ferritin, lactic acid dehydrogenase (LDH), or interleukin 6 (IL-6), elevated neutrophils, reduced lymphocytes and low albumin), and evidence of clinically severe illness requiring hospitalization, with multisystem (>2) organ involvement (cardiac, renal, respiratory, hematologic, gastrointestinal, dermatologic or neurological); AND   - No alternative plausible diagnoses; AND   - Positive for current or recent SARS-CoV-2 infection by RT-PCR, serology, or antigen test; or COVID-19 exposure within the 4 weeks prior to the onset of symptoms. - Admission to an intensive care unit (ICU). - Death. |

Abbreviations: AMI = acute myocardial infarction; ARDS = acute respiratory distress syndrome; BiPAP = bi-level positive airway pressure; Covid-19 = coronavirus disease 2019; CPAP = continuous positive air pressure; CT = computerized tomography; DBP = diastolic blood pressure; DVT = deep vein thrombosis; ECMO = extracorporeal membrane oxygenation; FiO_2_ = fraction of inspired oxygen; ICU = intensive care unit; LRTI = lower respiratory tract infection; NIV = non-invasive ventilation; NIPPV = non-invasive positive pressure ventilation; PaO_2_ = partial pressure of oxygen in the alveolus; PE = pulmonary embolism; SBP = systolic blood pressure; SpO_2_ = oxygen saturation.

*	Participants with a single vital sign abnormality placing them in the moderate or severe categories must also meet the criteria for mild Covid-19.

**Table S4. Demographics and Baseline Characteristics (Safety Analysis Set)**

| **Parameter** | **NVX-CoV2373**  **N = 1487** | **Placebo**  **N = 745** | **Total**  **N = 2232** |
| --- | --- | --- | --- |
| **Age (years)** | | | |
| Mean (SD)  AGE | 13.9 (1.4) | 13.8(1.4) | 13.8 (1.4) |
| Median | 14.0 | 14.0 | 14.0 |
| Min, max | 12, 17 | 12, 17 | 12, 17 |
| **Age group, n (%)** | | | |
| 12 to < 15 years | 998 (67.1) | 500 (67.1) | 1498 (67.1) |
| 15 to < 18 years  AGEGR1 | 489 (32.9) | 245 (32.9) | 734 (32.9) |
| **Sex, n (%)**  SEX | | | |
| Male | 756 (50.8) | 416 (55.8) | 1172 (52.5) |
| Female | 731 (49.2) | 329 (44.2) | 1060 (47.5) |
| **Race, n (%)** | | | |
| White | 1115 (75.0) | 545 (73.2) | 1660 (74.4) |
| Black or African American | 202 (13.6) | 108 (14.5) | 310 (13.9) |
| American Indian or Alaska Native | 32 (2.2) | 14 (1.9) | 46 (2.1) |
| Asian  RACEL | 43 (2.9) | 34 (4.6) | 77 (3.4) |
| Multiple | 82 (5.5) | 37 (5.0) | 119 (5.3) |
| Native Hawaiian or Other Pacific Islander | 3 (0.2) | 2 (0.3) | 5 (0.2) |
| Not reported | 10 (0.7) | 5 (0.7) | 15 (0.7) |
| **Ethnicity, n (%)**  ETHNIC | | | |
| Hispanic or Latino | 1208 (81.2) | 607 (81.5) | 1815 (81.3) |
| Not Hispanic or Latino | 274 (18.4) | 138 (18.5) | 412 (18.5) |
| Not reported | 2 (0.1) | 0 | 2 (< 0.1) |
| Unknown | 3 (0.2) | 0 | 3 (0.1) |
| **BMI category, n (%)** | | | |
| Underweight (<18.0 kg/m^2^) | 40 (2.7) | 28 (3.8) | 68 (3.0) |
| Normal (18.0–24.9 kg/m^2^) | 771 (51.8) | 417 (56.0) | 1188 (53.2) |
| Overweight (25.0–29.9 kg/m^2^) | 270 (18.2) | 107 (14.4) | 377 (16.9) |
| Obese (≥30.0 kg/m^2^) | 406 (27.3) | 193 (25.9) | 599 (26.8) |
| **Baseline serostatus, n (%)** | | | |
| **Anti-NP/ PCR*** | | | |
| Positive | 234 (15.7) | 125 (16.8) | 359 (16.1) |
| Negative | 1252 (84.2) | 620 (83.2) | 1872 (83.9) |
| Missing | 1 (< 0.1) | 0 | 1 (< 0.1) |

Abbreviations: BMI = body mass index; max = maximum; min = minimum; NP = nucleocapsid; NVX-CoV2373 = 5 μg SARS-CoV-2 rS with 50 μg Matrix-M™ adjuvant; PCR = polymerase chain reaction; SARS-CoV-2 rS = severe acute respiratory syndrome coronavirus 2 recombinant spike protein nanoparticle vaccine; SD = standard deviation.

BMI was classified as follows (using gender and age specific percentiles): Underweight = subjects less than the 5th percentile; Healthy weight = subjects within the 5th percentile and up to the 85th percentile; Overweight = subjects within the 85^th^ percentile to less than the 95th percentile;
 Obesity = subjects equal to or greater than the 95th percentile.

* Participants with either anti-NP or PCR were reported.

Figure S1. A) Box Plot of Neutralizing Antibody Titers for SARS-CoV-2 Wild-Type Virus at Specified Time Points in Baseline Serologically Negative/PCR-negative Adolescent Participants 12 to < 18 Years of Age (PP-IMM Analysis Set), B) Participants 12 to < 15 Years of Age (PP-IMM Analysis Set), and C) Participants 15 to < 18 Years of Age (PP-IMM Analysis Set)

**A)**

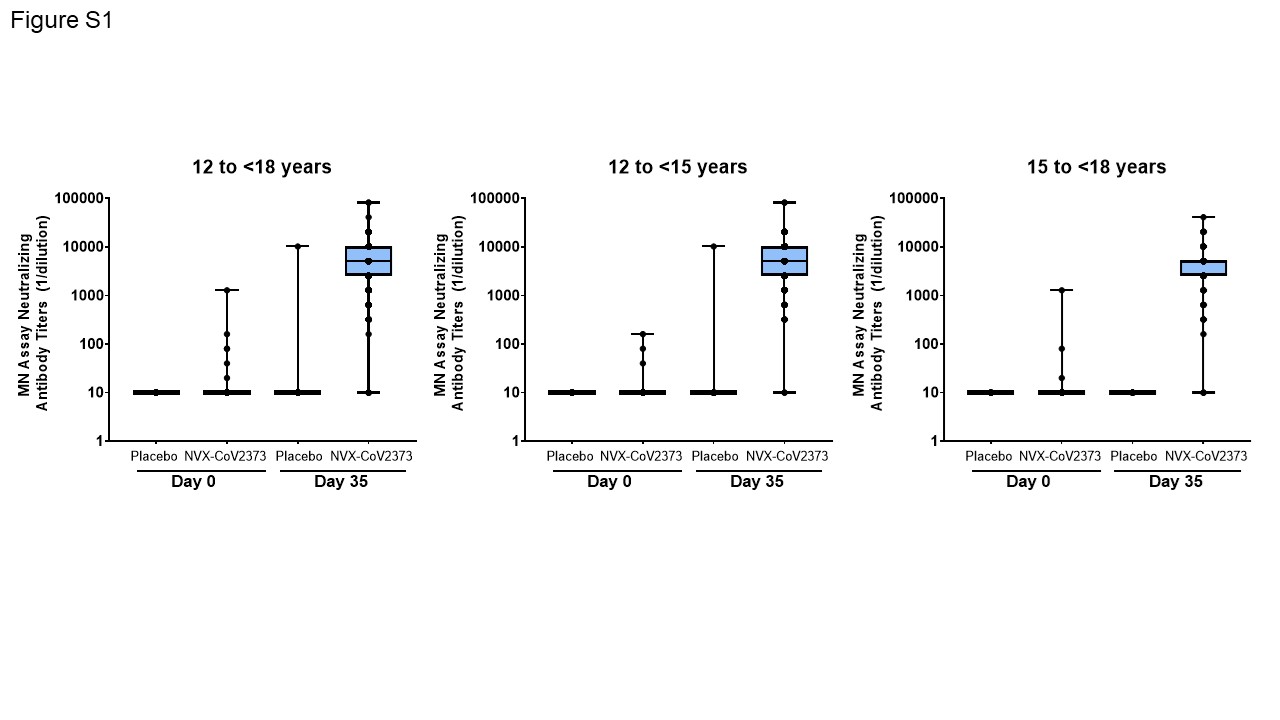

B)

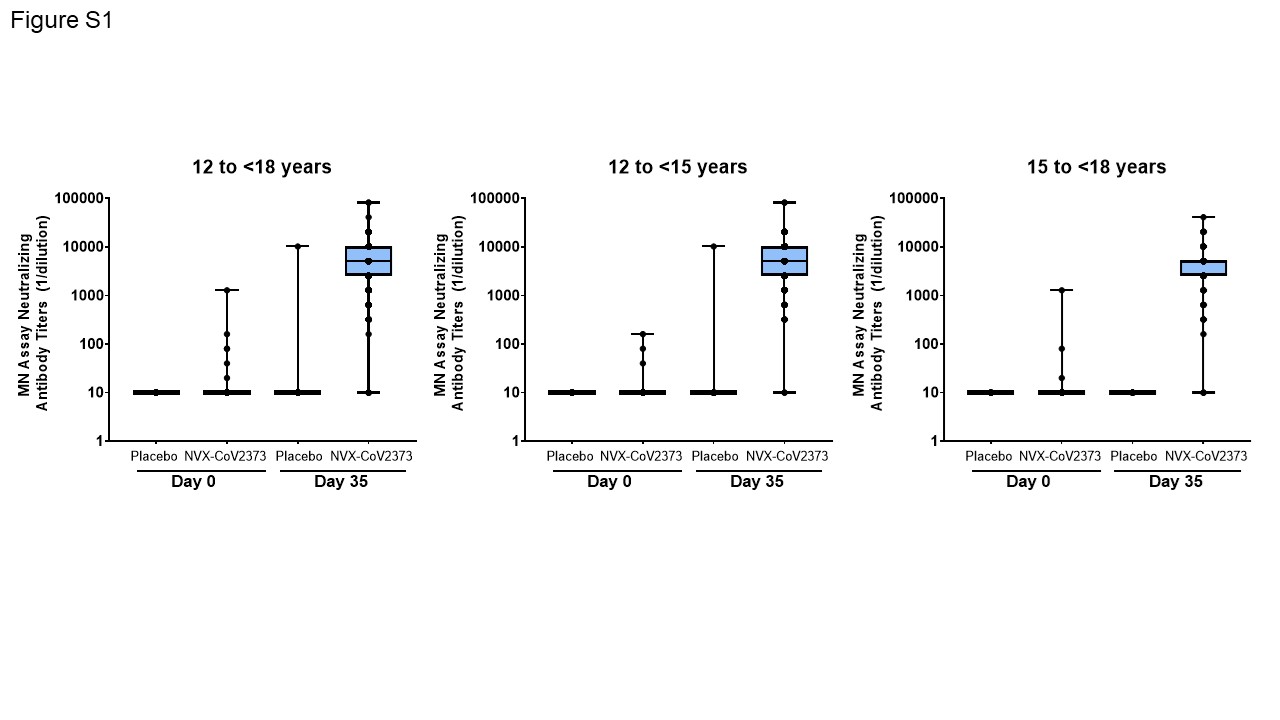

**C)**

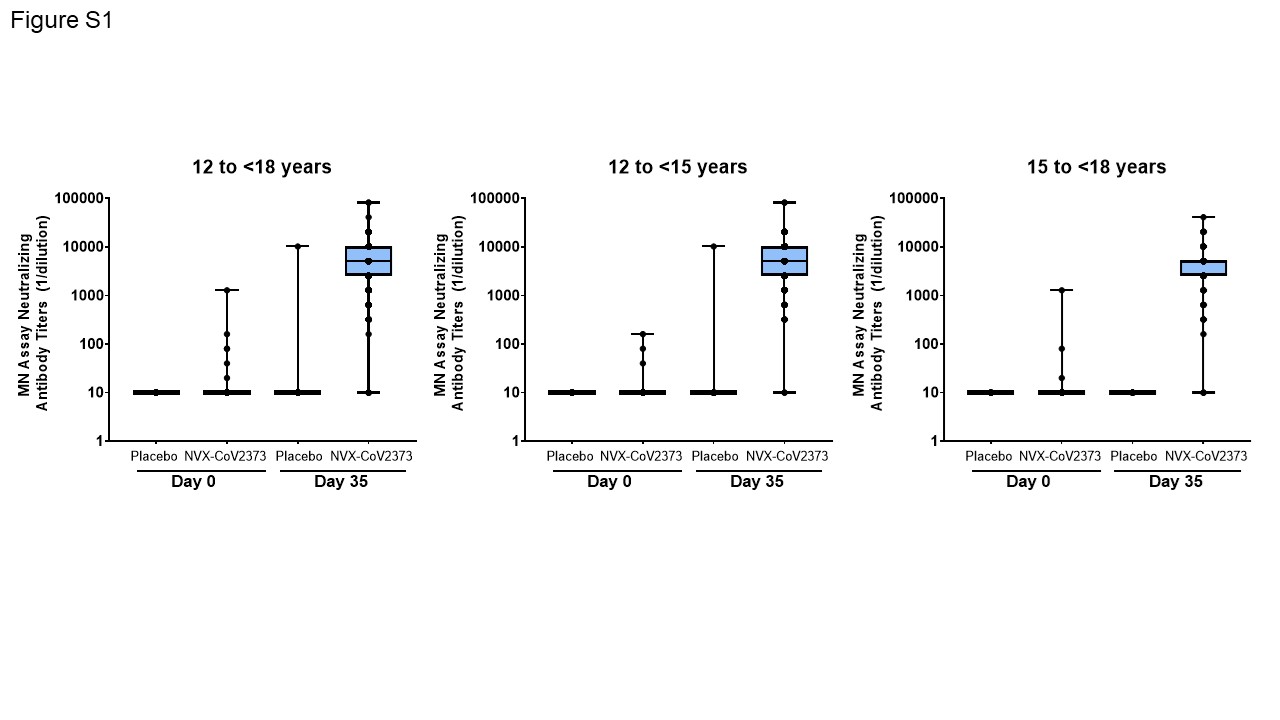

Abbreviations: LLOQ = lower limit of quantification; MN = microneutralization; NVX-CoV2373 = 5 µg SARS-CoV-2 rS + 50 µg Matrix-M adjuvant; PP‑IMM = Per-Protocol Immunogenicity; SARS-CoV-2 = severe acute respiratory syndrome coronavirus 2; SARS-CoV-2rS = NVX-CoV2373.

Note, titer values less than LLOQ (20) were replaced by 0.5 × LLOQ.

Data source: validated microneutralization assay conducted by 360biolabs.^9^

**Figure S2. Box Plot of Serum IgG Antibody Concentrations to SARS-CoV-2 S Protein in Baseline Serologically Negative/PCR-negative Adolescent Participants 12 to < 18 Years of Age (PP-IMM Analysis Set)**

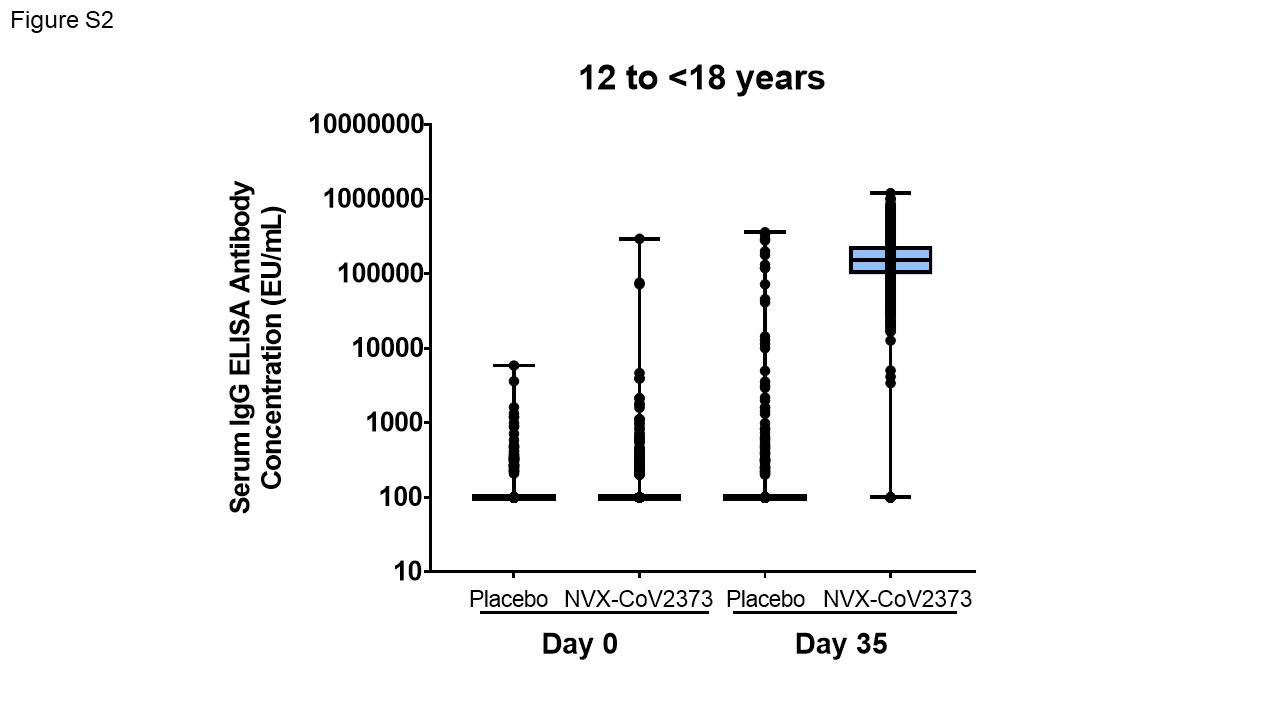

Abbreviations: ELISA = enzyme-linked immunosorbent assay; IgG = immunoglobulin G; LLOQ = lower limit of quantification; NVX-CoV2373 = 5 µg SARS-CoV-2 rS + 50 µg Matrix-M adjuvant; PP‑IMM-2 = Per-Protocol Immunogenicity 2; SARS-CoV-2 = severe acute respiratory syndrome coronavirus 2; SARS-CoV-2rS = NVX-CoV2373.

Note, titer values less than LLOQ (20) were replaced by 0.5 × LLOQ.

Data source: validated serum IgG ELISA assay conducted by Clinical Immunology Laboratory, Novavax.^6^

**Figure S3. Box Plot of hACE2 Inhibition Antibodies to SARS-CoV-2 S Protein in Baseline Serologically Negative/PCR-negative Adolescent Participants 12 to < 18 Years of Age (PP-IMM Analysis Set)**

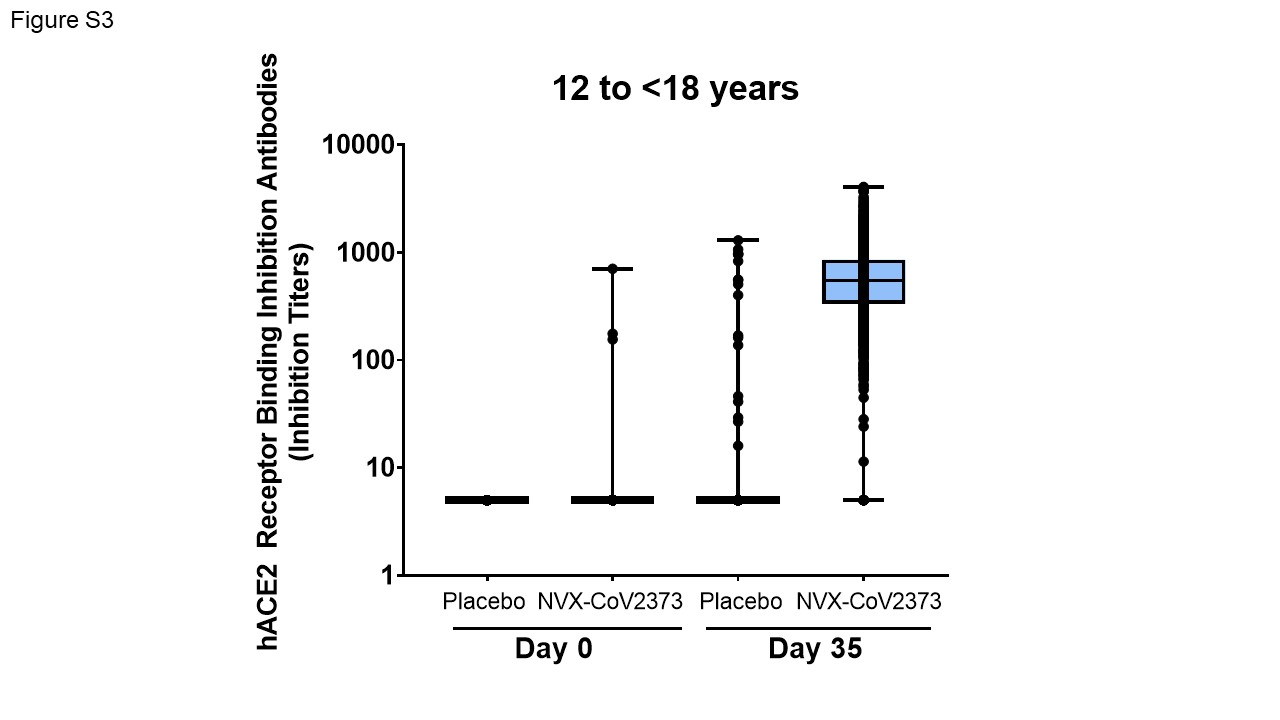

Abbreviations: hACE2 = human angiotensin-converting enzyme 2; LLOQ = lower limit of quantification; NVX‑CoV2373 = 5 µg SARS-CoV-2 rS + 50 µg Matrix-M adjuvant; PP‑IMM = Per-Protocol Immunogenicity; S = spike; SARS-CoV-2 = severe acute respiratory syndrome coronavirus 2; SARS-CoV-2rS = NVX-CoV2373.

Note, titer values less than LLOQ (10 inhibition titers) were replaced by 0.5 × LLOQ.

Data source: validated hACE2 receptor binding inhibition assay conducted by Clinical Immunology laboratory, Novavax.^7^

**Figure S4. Serum IgG Antibody Concentrations to SARS-CoV-2 S Protein From Different Variants in Baseline Serologically Negative/PCR-negative Adolescent Participants 12 to <18 Years of Age (N=40, PP-IMM Analysis Set, *Post Hoc* Analysis, Non-Validated, Fit-for-Purpose Assay)**

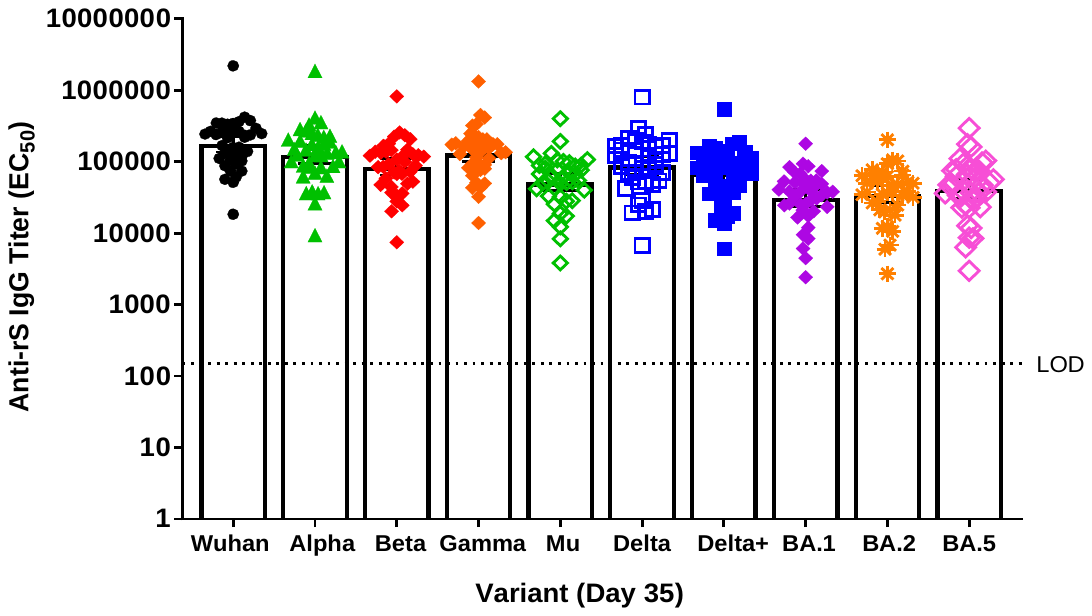

IgG = immunoglobulin G; EC_50_ = half maximal effective concentration; LOD: limit of detection; rS = recombinant spike protein. Data source: fit-for-purpose assay conducted by Vaccine Immunology Laboratory, Novavax.

**Figure S5. hACE2 Inhibition Antibodies to SARS-CoV-2 S Protein From Different Variants in Baseline Serologically Negative/PCR-negative Adolescent Participants 12 to <18 Years of Age (N=40, PP-IMM Analysis Set, *Post Hoc* Analysis, Non-Validated Fit-for-Purpose Assay)**

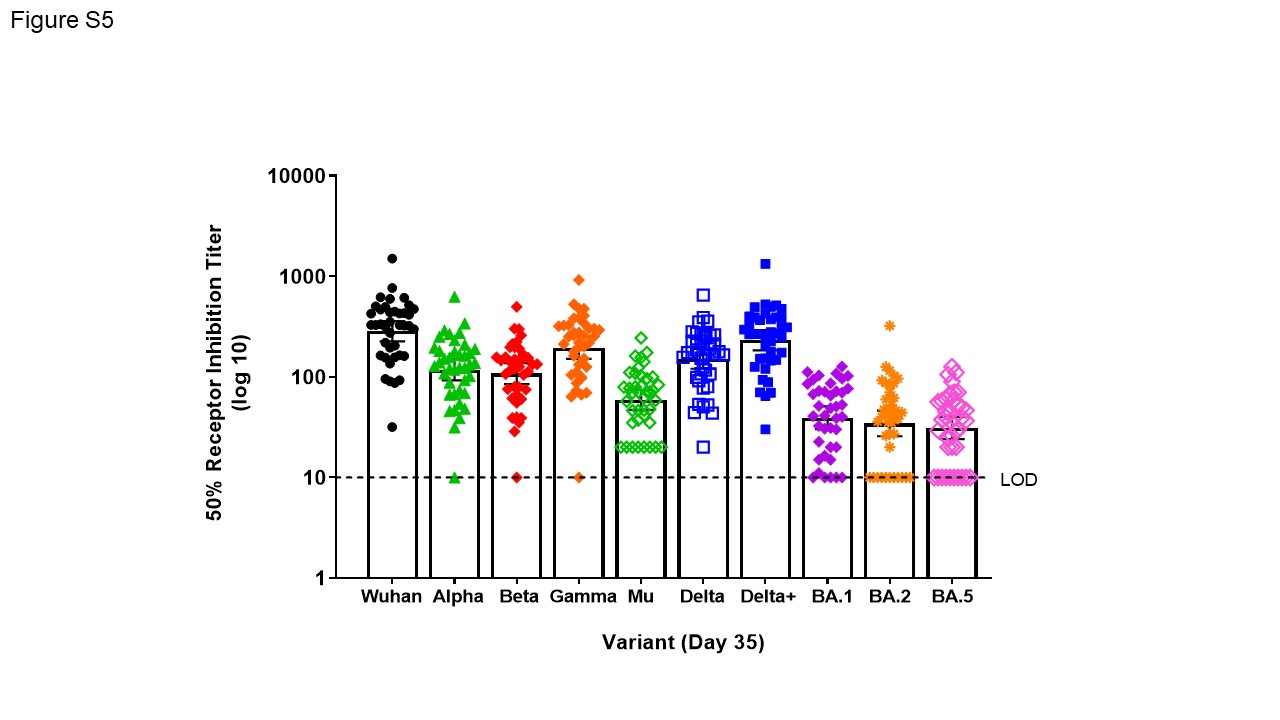

LOD: limit of detection. Data source: fit-for-purpose assay conducted by Vaccine Immunology Laboratory, Novavax.

**Table S5. Vaccine Efficacy Against RT-PCR-Confirmed Symptomatic Mild, Moderate, or Severe Covid-19 at Least 7 Days After Second Vaccination Due to Any SARS-CoV-2 Variant in Adolescent Participants Not Previously Exposed to SARS-CoV-2 (PP-EFF Analysis Set)**

| Parameter | NVX-CoV2373  N = 1205 | Placebo  N = 594 |
| --- | --- | --- |
| Participants with no occurrence of event,* n (%) | 1199 (99.5) | 580 (97.6) |
| Participants with occurrence of event,^†^ n (%) | 6 (0.5) | 14 (2.4) |
| Severity of first occurrence, n (%) | | |
| Mild | 6 (0.5) | 14 (2.4) |
| Moderate | 0 | 0 |
| Severe | 0 | 0 |
| Median surveillance time^‡^ (days) | 64.0 | 63.0 |
| Minimum - maximum | 1-135 | 1-118 |
| Log-linear model using modified Poisson regression^§^ | | |
| Mean disease incidence rate per year in 1000 people | 2.90 | 14.20 |
| 95% CI | 1.31, 6.46 | 8.42, 23.93 |
| Relative risk | 0.20 | |
| 95% CI | 0.08, 0.53 | |
| Vaccine efficacy (%) | 79.54 | |
| 95% CI | 46.83, 92.13 | |
| Cox proportional hazard model (sensitivity analysis)^‖^ | | |
| Vaccine efficacy (%) | 79.39 | |
| 95% CI | 46.34, 92.08 | |

Abbreviations: CI = confidence interval; Covid-19 = coronavirus disease 2019; NVX-CoV2373 = 5 μg SARS-CoV-2 rS with 50 μg Matrix-M™ adjuvant; RT-PCR = reverse transcriptase-polymerase chain reaction; PP-EFF = Per-Protocol Efficacy; SARS-CoV-2 rS = severe acute respiratory syndrome coronavirus 2 recombinant spike protein nanoparticle vaccine; VE = vaccine efficacy.

* Includes participants with RT-PCR-confirmed infection who did not meet mild, moderate, or severe Covid-19 criteria.

^†^	Event = first occurrence of RT-PCR-confirmed mild, moderate, or severe Covid-19 with onset of illness episode from at least 7 days after second vaccination within the surveillance period.

^‡^	Surveillance time was defined as the difference between the date at end of surveillance period (onset of first occurrence of event/ censoring) and date at start of surveillance period (7 days after the second injection) + 1.

^§^	Modified Poisson regression with logarithmic link function, treatment group as fixed effects and robust error variance.^8^

^‖^	Cox-proportional hazard model with Efron’s method for tie handling with vaccine group. Hazard ratio was used to estimate relative risk.

**Table S6.** **Vaccine Efficacy Against RT-PCR-Confirmed Symptomatic Mild, Moderate or Severe Covid-19 at Least 7 Days After Second Vaccination Due to the SARS-CoV-2 Delta Variant in Adolescent Participants Not Previously Exposed to SARS-CoV-2 (PP-EFF Analysis Set)**

| Parameter | NVX-CoV2373  N = 1205 | Placebo  N = 594 |
| --- | --- | --- |
| Participants with no occurrence of event,* n (%) | 1202 (99.8) | 586 (98.7) |
| Participants with occurrence of event,^†^ n (%) | 3 (0.2) | 8 (1.3) |
| Severity of first occurrence, n (%) | | |
| Mild | 3 (0.2) | 8 (1.3) |
| Moderate | 0 | 0 |
| Severe | 0 | 0 |
| Median surveillance time^‡^ (days) | 64.0 | 63.0 |
| Minimum - maximum | 1-135 | 1-118 |
| Log-linear model using modified Poisson regression^§^ | | |
| Mean disease incidence rate per year in 1000 people | 1.45 | 8.07 |
| 95% CI | 0.47, 4.49 | 4.04, 16.11 |
| Relative risk | 0.18 | |
| 95% CI | 0.05, 0.68 | |
| Vaccine efficacy (%) | 82.04 | |
| 95% CI | 32.42, 95.23 | |

Abbreviations: CI = confidence interval; Covid-19 = coronavirus disease 2019; NVX-CoV2373 = 5 μg SARS-CoV-2 rS with 50 μg Matrix-M™ adjuvant; RT-PCR = reverse transcriptase-polymerase chain reaction; PP-EFF = Per-Protocol Efficacy; SARS-CoV-2 rS = severe acute respiratory syndrome coronavirus 2 recombinant spike protein nanoparticle vaccine; VE = vaccine efficacy.

* Includes participants with RT-PCR-confirmed infection who did not meet mild, moderate, or severe Covid-19 criteria due to the Delta variant.

^†^	Event = first occurrence of RT-PCR-confirmed mild, moderate, or severe Covid-19 due to a VOC or VOI with onset of illness episode from at least 7 days after second vaccination within the surveillance period.

^‡^	Surveillance time was defined as the difference between the date at end of surveillance period (onset of first occurrence of event/censoring) and date at start of surveillance period (7 days after the second injection) + 1.

^§^	Modified Poisson regression with logarithmic link function, treatment group as fixed effects and robust error variance.^8^

**Table S7. Overall Summary of Treatment-Emergent Adverse Events Reported Between Start of First Vaccination and Blinded Crossover or Early Withdrawal (Safety Analysis Set)**

| TEAE Category | NVX-CoV2373  N = 1487 | | Placebo  N = 745 | |
| --- | --- | --- | --- | --- |
|  | n (%) | E | n (%) | E |
| Any TEAE | 236 (15.9) | 386 | 116 (15.6) | 178 |
| Any severe TEAE* | 6 (0.4) | 7 | 2 (0.3) | 2 |
| Any treatment-related TEAE * | 43 (2.9) | 62 | 7 (0.9) | 9 |
| Any severe treatment-related TEAE* | 0 | 0 | 0 | 0 |
| Any MAAE | 96 (6.5) | 129 | 51 (6.8) | 70 |
| Any treatment-related MAAE* | 5 (0.3) | 10 | 3 (0.4) | 4 |
| Any serious treatment-related MAAE* | 0 | 0 | 0 | 0 |
| Any serious TEAE | 7 (0.5) | 9 | 2 (0.3) | 2 |
| Any TEAE leading to vaccination discontinuation | 1 (< 0.1) | 1 | 1 (0.1) | 1 |
| Any treatment-related TEAE leading to vaccination discontinuation* | 0 | 0 | 0 | 0 |
| Any TEAE leading to study discontinuation | 0 | 0 | 0 | 0 |
| Any treatment-related TEAE leading to study discontinuation^*^ | 0 | 0 | 0 | 0 |
| Any AESI: PIMMC | 0 | 0 | 0 | 0 |
| Any treatment-related AESI: PIMMC* | 0 | 0 | 0 | 0 |
| Any AESI: relevant to Covid-19 | 0 | 0 | 0 | 0 |
| Any treatment-related AESI: relevant to Covid-19* | 0 | 0 | 0 | 0 |

Abbreviations: AESI = adverse event of special interest; Covid-19 = coronavirus disease 2019; E = number of events at each level of summarization; MAAE = medically attended adverse event; NVX-CoV2373 = 5 μg SARS-CoV-2 rS with 50 μg Matrix-M™ adjuvant; PIMMC = potential immune-mediated medical conditions; SARS-CoV-2 = severe acute respiratory syndrome coronavirus 2; SARS-CoV-2 rS = severe acute respiratory syndrome coronavirus 2 recombinant spike protein nanoparticle vaccine; TEAE = treatment-emergent adverse event.

* Relationship and severity were based on the data reported by site, i.e., missing information was not imputed.

Note: Events indicated as continuing from the reactogenicity period by the clinic site were excluded from this presentation of TEAEs.

Note: At each level of participant summarization, a participant was counted once if the participant reported ≥ 1 events.

**Figure S6. Solicited Local and Systemic Adverse Events by Age Subgroup.**

The percentage of participants vaccinated with NVX-CoV2373 in each age group with solicited local (A) and systemic (B) adverse events during the 7 days after each vaccination is plotted by FDA toxicity grade, either as any grade (mild, moderate, severe, or potentially life-threatening) or as Grade 3+ (severe or potentially life-threatening).^10^

**A**

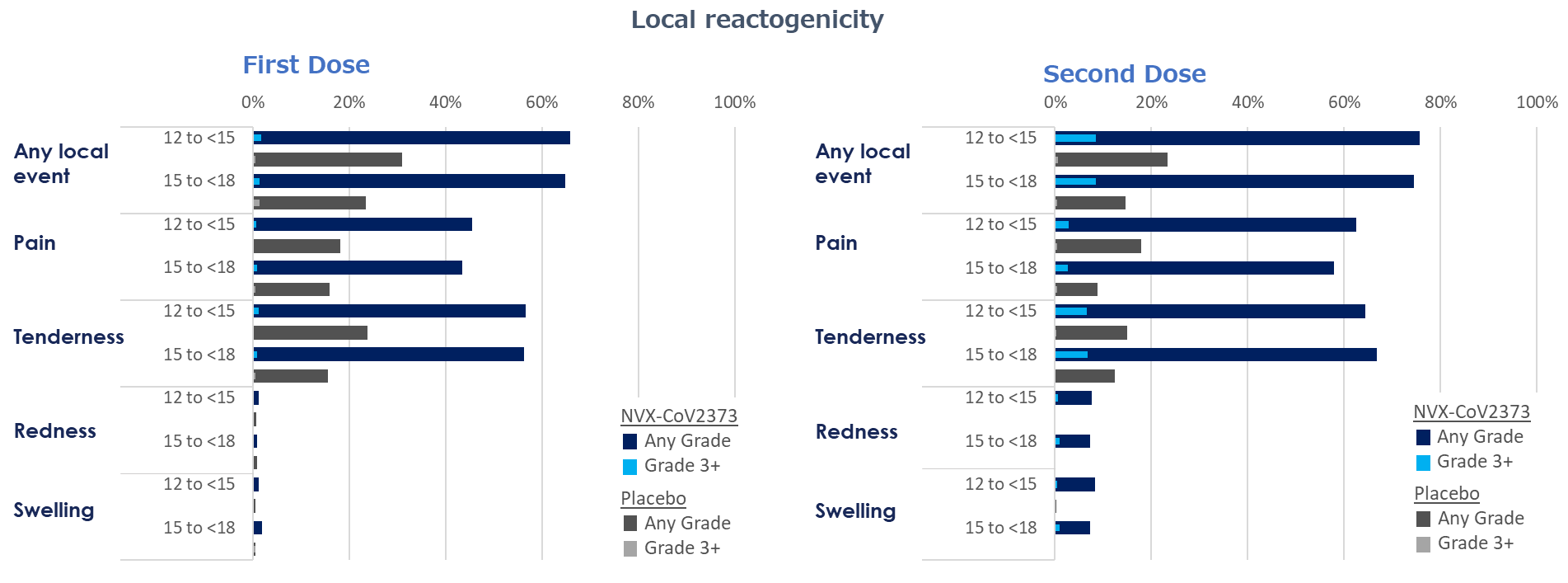

**B**

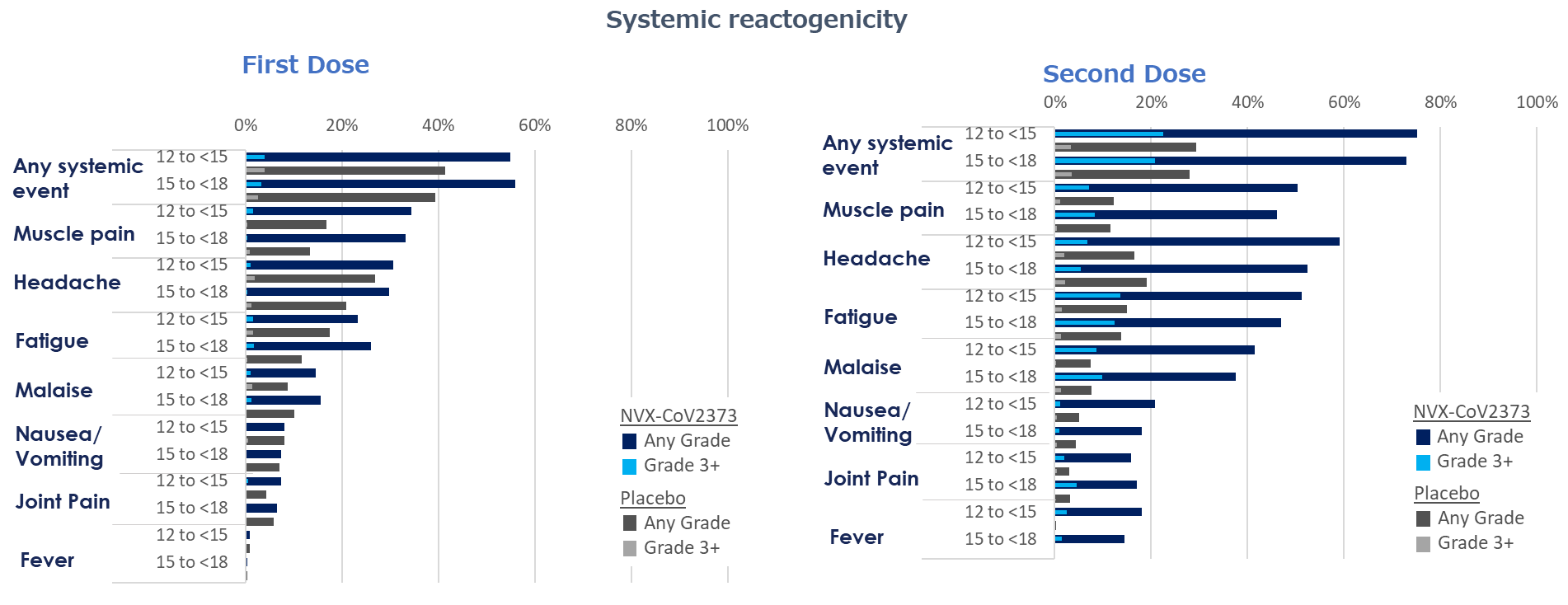

**Table S8. Summary of Solicited Local Adverse Events Within 7 Days After Dose 1 and Dose 2 in All Participants (Safety Analysis Set)**

| Solicited Local Adverse Events | All Participants | |
| --- | --- | --- |
|  | **NVX-CoV2373**  **N = 1487** | **Placebo**  **N = 745** |
| Any local adverse event, N1/N2 | **1448/1394** | **726/686** |
| Dose 1 (any grade) | 948 (65.5) | 207 (28.5) |
| *Grade 3* | 22 (1.5) | 5 (0.7) |
| *Grade 4* | 0 | 0 |
| Dose 2 (any grade) | 1050 (75.3) | 141 (20.6) |
| *Grade* *3* | 118 (8.5) | 4 (0.6) |
| *Grade 4* | 0 | 0 |
| **Any pain, N1/N2** | **1448/1394** | **726/686** |
| Dose 1 (any grade) | 648 (44.6) | 126 (17.4) |
| *Grade* *3* | 10 (0.7) | 2 (0.3) |
| *Grade 4* | 0 | 0 |
| Dose 2 (any grade) | 850 (61.0) | 102 (14.9) |
| *Grade* *3* | 38 (2.7) | 3 (0.4) |
| *Grade 4* | 0 | 0 |
| **Any tenderness, N1/N2** | **1448/1394** | **726/686** |
| Dose 1 (any grade) | 817 (56.4) | 153 (21.1) |
| *Grade* *3* | 16 (1.1) | 2 (0.3) |
| *Grade 4* | 0 | 0 |
| Dose 2 (any grade) | 909 (65.2) | 97 (14.1) |
| *Grade* *3* | 93 (6.7) | 1 (0.1) |
| *Grade 4* | 0 | 0 |
| **Any erythema, N1/N2** | **1448/1394** | **726/686** |
| Dose 1 (any grade) | 15 (1.0) | 5 (0.7) |
| *Grade* *3* | 0 | 0 |
| *Grade 4* | 0 | 0 |
| Dose 2 (any grade) | 104 (7.5) | 0 |
| *Grade* *3* | 10 (0.7) | 0 |
| *Grade 4* | 0 | 0 |
| **Any swelling, N1/N2** | **1448/1394** | **726/686** |
| Dose 1 (any grade) | 20 (1.4) | 3 (0.4) |
| *Grade 3* | 0 | 1 (0.1) |
| *Grade 4* | 0 | 0 |
| Dose 2 (any grade) | 111 (8.0) | 1 (0.1) |
| *Grade 3* | 8 (0.6) | 0 |
| *Grade 4* | 0 | 0 |

Abbreviations: FDA = US Food and Drug Administration; N = number of participants in the Safety Analysis Set following Dose 1/Dose 2; N1 = number of participants in the Safety Analysis Set who received the first dose and completed at least 1 day of the reactogenicity diary; N2 = number of participants in the Safety Analysis Set who received the second dose and completed at least 1 day of the reactogenicity diary; NVX-CoV2373 = 5 μg SARS-CoV-2 rS with 50 μg Matrix-M™ adjuvant; SARS-CoV-2 rS = severe acute respiratory syndrome coronavirus 2 recombinant spike protein nanoparticle vaccine.

Note: Data are presented as number (%) of participants experiencing a solicited event. Percentages were based on n/N1 × 100 and n/N2 × 100. At each level of participant summarization, a participant was counted once if they indicated the event. Any grade pertains to reactions reported at grade ≥ 1. Grading of solicited adverse events was based on FDA Toxicity Grading Scale for Clinical Abnormalities.^10^

**Table S9. Duration (Days) of Solicited Local Adverse Events Within 7 Days After Dose 1 and Dose 2 in All Participants (Safety Analysis Set)**

| Solicited Local Adverse Events | **NVX-CoV2373**  N = 1487 | **Placebo**  N = 745 |
| --- | --- | --- |
| **Pain (# of days ≥ grade 1), N1/N2** | **1448/1394** | **726/686** |
| Dose 1, n | 648 | 126 |
| Median | 2.0 | 1.0 |
| Minimum - maximum | 1 - 7 | 1 - 6 |
| Dose 2, n | 850 | 102 |
| Median | 2.0 | 1.0 |
| Minimum - maximum | 1 - 7 | 1 - 6 |
| **Tenderness (# of days ≥ grade 1), N1/N2** | **1448/1394** | **726/686** |
| Dose 1, n | 817 | 153 |
| Median | 2.0 | 1.0 |
| Minimum - maximum | 1 - 7 | 1 - 7 |
| Dose 2, n | 909 | 97 |
| Median | 2.0 | 1.0 |
| Minimum - maximum | 1 - 7 | 1 - 7 |
| **Erythema (# of days ≥ grade 1), N1/N2** | **1448/1394** | **726/686** |
| Dose 1, n | 15 | 5 |
| Median | 2.0 | 1.0 |
| Minimum - maximum | 1 - 4 | 1 - 1 |
| Dose 2, n | 104 | 0 |
| Median | 2.0 | N/A |
| Minimum - maximum | 1 - 6 | N/A |
| **Swelling (# of days ≥ grade 1), N1/N2** | **1448/1394** | **726/686** |
| Dose 1, n | 20 | 3 |
| Median | 1.0 | 1.0 |
| Minimum - maximum | 1 - 5 | 1 - 2 |
| Dose 2, n | 111 | 1 |
| Median | 2.0 | 1.0 |
| Minimum - maximum | 1 - 7 | 1 - 1 |
| Abbreviations: n = number of participants who reported the solicited event; N = number of participants in the Safety Analysis Set following Dose 1/Dose 2; N1 = number of participants in the Safety Analysis Set who received the first dose and completed at least 1 day of the reactogenicity diary; N2 = number of participants in the Safety Analysis Set who received the second dose and completed at least 1 day of the reactogenicity diary; NVX-CoV2373 = 5 μg SARS-CoV-2 rS with 50 μg Matrix-M™ adjuvant; SARS-CoV-2 rS = severe acute respiratory syndrome coronavirus 2 recombinant spike protein nanoparticle vaccine.  Note: Duration is calculated as the number of days the solicited event was greater than grade 0. n = number of subjects who reported the event. | | |

**Table S10. Summary of Solicited Systemic Adverse Events Within 7 Days After Dose 1 and Dose 2 by Age Group (Safety Analysis Set)**

| Solicited Systemic Adverse Events | All Participants | |
| --- | --- | --- |
|  | **NVX-CoV2373**  **N = 1487** | **Placebo**  **N = 745** |
| **Any solicited systemic TEAE, N1/N2** | **1448/1394** | **726/686** |
| Dose 1 (Grade ≥1) | 800 (55.2) | 296 (40.8) |
| *Grade* *3* | 52 (3.6) | 25 (3.4) |
| *Grade 4* | 2 (0.1) | 0 |
| Dose 2 (Grade ≥1) | 1038 (74.5) | 198 (28.9) |
| *Grade* *3* | 304 (21.8) | 23 (3.4) |
| *Grade 4* | 2 (0.1) | 0 |
| **Headache, N1/N2** | **1448/1394** | **726/686** |
| Dose 1 (Grade ≥1) | 440 (30.4) | 181 (24.9) |
| *Grade* *3* | 13 (0.9) | 12 (1.7) |
| *Grade 4* | 0 | 0 |
| Dose 2 (Grade ≥1) | 793 (56.9) | 119 (17.3) |
| *Grade* *3* | 87 (6.2) | 14 (2.0) |
| *Grade 4* | 1(< 0.1) | 0 |
| **Fatigue, N1/N2** | **1448/1394** | **726/686** |
| Dose 1 (Grade ≥1) | 350 (24.2) | 113 (15.6) |
| *Grade* *3* | 23 (1.6) | 9 (1.2) |
| *Grade 4* | 0 | 0 |
| Dose 2 (Grade ≥1) | 695 (49.9) | 100 (14.6) |
| *Grade* *3* | 185 (13.3) | 10 (1.5) |
| *Grade 4* | 0 | 0 |
| **Malaise, N1/N2** | **1448/1394** | **726/686** |
| Dose 1 (Grade ≥1) | 215 (14.8) | 67 (9.2) |
| *Grade* *3* | 16 (1.1) | 7 (1.0) |
| *Grade 4* | 0 | 0 |
| Dose 2 (Grade ≥1) | 560 (40.2) | 51 (7.4) |
| *Grade 3* | 126 (9.0) | 4 (0.6) |
| *Grade 4* | 0 | 0 |
| **Muscle pain, N1/N2** | **1448/1394** | **726/686** |
| Dose 1 (Grade ≥1) | 492 (34.0) | 114 (15.7) |
| *Grade* *3* | 17 (1.2) | 4 (0.6) |
| *Grade 4* | 0 | 0 |
| Dose 2 (Grade ≥1) | 683 (49.0) | 82 (12.0) |
| *Grade 3* | 104 (7.5) | 6 (0.9) |
| *Grade 4* | 0 | 0 |
| **Joint pain, N1/N2** | **1448/1394** | **726/686** |
| Dose 1 (Grade ≥1) | 102 (7.0) | 35 (4.8) |
| *Grade* *3* | 6 (0.4) | 1 (0.1) |
| *Grade 4* | 0 | 0 |
| Dose 2 (Grade ≥1) | 226 (16.2) | 21 (3.1) |
| *Grade 3* | 40 (2.9) | 2 (0.3) |
| *Grade 4* | 0 | 0 |
| **Fever, N1/N2** | **1448/1394** | **726/686** |
| Dose 1 (Grade ≥1) | 11 (0.8) | 5 (0.7) |
| *Grade* *3* | 1 (< 0.1) | 0 |
| *Grade 4* | 2 (0.1) | 0 |
| Dose 2 (Grade ≥1) | 235 (16.9) | 1 (0.1) |
| *Grade 3* | 31 (2.2) | 0 |
| *Grade 4* | 0 | 0 |
| **Nausea/Vomiting, N1/N2** | **1448/1394** | **726/686** |
| Dose 1 (Grade ≥1) | 113 (7.8) | 56 (7.7) |
| *Grade* *3* | 2 (0.1) | 3 (0.4) |
| *Grade 4* | 0 | 0 |
| Dose 2 (Grade ≥1) | 277 (19.9) | 33 (4.8) |
| *Grade 3* | 14 (1.0) | 3 (0.4) |
| *Grade 4* | 1 (< 0.1) | 0 |

Abbreviations: FDA = US Food and Drug Administration; N = number of participants in the Safety Analysis Set following Dose 1/Dose 2; N1 = number of participants in the Safety Analysis Set who received the first dose and completed at least 1 day of the reactogenicity diary; N2 = number of participants in the Safety Analysis Set who received the second dose and completed at least 1 day of the reactogenicity diary; NVX-CoV2373 = 5 μg SARS-CoV-2 rS with 50 μg Matrix-M™ adjuvant; SARS-CoV-2 rS = severe acute respiratory syndrome coronavirus 2 recombinant spike protein nanoparticle vaccine; TEAE = treatment-emergent adverse event.

Note: Data are presented as number (%) of participants experiencing a solicited event. Percentages were based on n/N1 × 100 and n/N2 × 100. At each level of participant summarization, a participant was counted once if they indicated the event occurred and provided a severity during the reactogenicity period. The highest severity experienced during the reactogenicity period is summarized in this table.

Note: Grading of solicited adverse events was based on FDA Toxicity Grading Scale for Clinical Abnormalities.^10^

**Table S11. Duration (Days) of Solicited Systemic Adverse Events Within 7 Days After Dose 1 and Dose 2 in All Participants (Safety Analysis Set)**

| Solicited Systemic Adverse Events | **NVX-CoV2373**  N = 1487 | **Placebo**  N = 745 |
| --- | --- | --- |
| **Fatigue (# of days ≥ grade 1), N1/N2** | **1448/1394** | **726/686** |
| Dose 1, n | 350 | 113 |
| Median | 1.0 | 1.0 |
| Minimum - maximum | 1-7 | 1-6 |
| Dose 2, n | 695 | 100 |
| Median | 1.0 | 1.0 |
| Minimum - maximum | 1-7 | 1-6 |
| **Fever (# of days ≥ grade 1), N1/N2** | **1448/1394** | **726/686** |
| Dose 1, n | 11 | 5 |
| Median | 1.0 | 1.0 |
| Minimum - maximum | 1-3 | 1-2 |
| Dose 2, n | 235 | 1 |
| Median | 1.0 | 1.0 |
| Minimum - maximum | 1-2 | 1-1 |
| **Headache (# of days ≥ grade 1), N1/N2** | **1448/1394** | **726/686** |
| Dose 1, n | 440 | 181 |
| Median | 1.0 | 1.0 |
| Minimum - maximum | 1-7 | 1-7 |
| Dose 2, n | 793 | 119 |
| Median | 1.0 | 1.0 |
| Minimum - maximum | 1-7 | 1-6 |
| **Joint pain (# of days ≥ grade 1), N1/N2** | **1448/1394** | **726/686** |
| Dose 1, n | 102 | 35 |
| Median | 1.0 | 1.0 |
| Minimum - maximum | 1-6 | 1-5 |
| Dose 2, n | 225 | 21 |
| Median | 1.0 | 1.0 |
| Minimum - maximum | 1-6 | 1-5 |
| **Malaise (# of days ≥ grade 1), N1/N2** | **1448/1394** | **726/686** |
| Dose 1, n | 215 | 67 |
| Median | 1.0 | 1.0 |
| Minimum - maximum | 1-5 | 1-5 |
| Dose 2, n | 560 | 51 |
| Median | 1.0 | 1.0 |
| Minimum - maximum | 1-7 | 1-5 |
| **Muscle pain (# of days ≥ grade 1), N1/N2** | **1448/1394** | **726/686** |
| Dose 1, n | 492 | 114 |
| Median | 1.0 | 1.0 |
| Minimum - maximum | 1-7 | 1-7 |
| Dose 2, n | 683 | 82 |
| Median | 2.0 | 1.0 |
| Minimum - maximum | 1-6 | 1-6 |
| **Nausea/Vomiting (# of days ≥ grade 1), N1/N2** | **1448/1394** | **726/686** |
| Dose 1, n | 113 | 56 |
| Median | 1.0 | 1.0 |
| Minimum - maximum | 1-5 | 1-4 |
| Dose 2, n | 277 | 33 |
| Median | 1.0 | 1.0 |
| Minimum - maximum | 1-7 | 1-6 |
| Abbreviations: n = number of participants who reported the solicited event; N = number of participants in the Safety Analysis Set following Dose 1/Dose 2; N1 = number of participants in the Safety Analysis Set who received the first dose and completed at least 1 day of the reactogenicity diary; N2 = number of participants in the Safety Analysis Set who received the second dose and completed at least 1 day of the reactogenicity diary; NVX-CoV2373 = 5 μg SARS-CoV-2 rS with 50 μg Matrix-M™ adjuvant; SARS-CoV-2 rS = severe acute respiratory syndrome coronavirus 2 recombinant spike protein nanoparticle vaccine.  Note: Duration is calculated as the number of days the solicited event was greater than grade 0. n=number of subjects who reported the event. | | |

**Table S12. Overall Summary of Unsolicited Adverse Events by System Organ Class and Preferred Term Reported Within 49 Days After First Vaccination in at Least 0.5% of All Adolescent Participants in Any Study Vaccine Group by Age Strata (Safety Analysis Set)**

| System Organ Class/ Preferred Term  (MedDRA, Version 24.0) | Participants 12 to < 18 Years | | Participants 12 to < 15 Years | | Participants 15 to < 18 Years | |
| --- | --- | --- | --- | --- | --- | --- |
|  | NVX-CoV2373  N = 1478 | Placebo  N = 745 | NVX-CoV2373  N = 998 | Placebo  N = 500 | NVX-CoV2373  N = 489 | Placebo  N = 245 |
|  | n (%) | n (%) | n (%) | n (%) | n (%) | n (%) |
| **Any system organ class** | **234 (15.7)** | **115 (15.4)** | **174 (17.4)** | **82 (16.4)** | **60 (12.3)** | **33 (13.5)** |
| **Infections and infestations** | **61 (4.1)** | **41 (5.5)** | **43 (4.3)** | **30 (6.0)** | **18 (3.7)** | **11 (4.5)** |
| Upper respiratory tract infection | 10 (0.7) | 14 (1.9) | 9 (0.9) | 11 (2.2) | 1 (0.2) | 3 (1.2) |
| Viral infection | 10 (0.7) | 6 (0.8) | 4 (0.4) | 3 (0.6) | 6 (1.2) | 3 (1.2) |
| Nasopharyngitis | 6 (0.4) | 5 (0.7) | 4 (0.4) | 5 (1.0) | 2 (0.4) | 0 |
| **Respiratory, thoracic and mediastinal disorders** | **42 (2.8)** | **25 (3.4)** | **33 (3.3)** | **22 (4.4)** | **9 (1.8)** | **3 (1.2)** |
| Nasal congestion | 21 (1.4) | 10 (1.3) | 17 (1.7) | 8 (1.6) | 4 (0.8) | 2 (0.8) |
| Cough | 18 (1.2) | 6 (0.8) | 16 (1.6) | 5 (1.0) | 2 (0.4) | 1 (0.4) |
| Oropharyngeal pain | 16 (1.1) | 13 (1.7) | 12 (1.2) | 10 (2.0) | 4 (0.8) | 3 (1.2) |
| Rhinorrhea | 6 (0.4) | 8 (1.1) | 5 (0.5) | 8 (1.6) | 1 (0.2) | 0 |
| **General disorders and administration site conditions** | **34 (2.3)** | **7 (0.9)** | **21 (2.1)** | **5 (1.0)** | **13 (2.7)** | **2 (0.8)** |
| Fatigue | 7 (0.5) | 0 | 4 (0.4) | 0 | 3 (0.6) | 0 |
| **Injury, poisoning and procedural complications** | **38 (2.6)** | **15 (2.0)** | **27 (2.7)** | **8 (1.6)** | **11 (2.2)** | **7 (2.9)** |
| Ligament sprain | 3 (0.2) | 3 (0.4) | 3 (0.3) | 1 (0.2) | 0 | 2 (0.8) |
| Skin laceration | 1 (< 0.1) | 5 (0.7) | 1 (0.1) | 4 (0.8) | 0 | 1 (0.4) |
| **Nervous system disorders** | **24 (1.6)** | **14 (1.9)** | **15 (1.5)** | **10 (2.0)** | **9 (1.8)** | **4 (1.6)** |
| Headache | 12 (0.8) | 8 (1.1) | 9 (0.9) | 6 (1.2) | 3 (0.6) | 2 (0.8) |
| **Gastrointestinal disorders** | **27 (1.8)** | **18 (2.4)** | **17 (1.7)** | **14 (2.8)** | **10 (2.0)** | **4 (1.6)** |
| Nausea | 8 (0.5) | 5 (0.7) | 4 (0.4) | 5 (1.0) | 4 (0.8) | 0 |
| Diarrhea | 8 (0.5) | 6 (0.8) | 6 (0.6) | 6 (1.2) | 2 (0.4) | 0 |
| Vomiting | 5 (0.3) | 3 (0.4) | 3 (0.3) | 5 (1.0) | 2 (0.4) | 0 |
| **Psychiatric disorders** | **22 (1.5)** | **9 (1.2)** | **15 (1.5)** | **6 (1.2)** | **7 (1.4)** | **3 (1.2)** |
| Attention deficit hyperactivity disorder | 8 (0.5) | 2 (0.3) | 5 (0.5) | 2 (0.4) | 3 (0.6) | 0 |
| Anxiety | 2 (0.1) | 2 (0.3) | 1 (0.1) | 0 | 1 (0.2) | 2 (0.8) |
| **Musculoskeletal and connective tissue disorders** | **18 (1.2)** | **2 (0.3)** | **14 (1.4)** | **0** | **4 (0.8)** | **2 (0.8)** |
| Arthralgia | 3 (0.2) | 0 | 3 (0.3) | 0 | 0 | 0 |
| **Skin and subcutaneous tissue disorders** | **21 (1.4)** | **6 (0.8)** | **17 (1.7)** | **5 (1.0)** | **4 (0.8)** | **1 (0.4)** |
| Rash | 7 (0.5) | 2 (0.3) | 6 (0.6) | 2 (0.4) | 1 (0.2) | 0 |
| **Blood and lymphatic system disorders** | **11 (0.7)** | **0** | **9 (0.9)** | **0** | **2 (0.4)** | **0** |
| Lymphadenopathy | 10 (0.7) | 0 | 8 (0.8) | 0 | 2 (0.4) | 0 |
| **Ear and labyrinth disorders** | **4 (0.3)** | **2 (0.3)** | **4 (0.4)** | **0** | **0** | **2 (0.8)** |
| **Eye disorders** | **7 (0.5)** | **1 (0.1)** | **6 (0.6)** | **0** | **1 (0.2)** | **1 (0.4)** |

Abbreviations: MedDRA = Medical Dictionary for Regulatory Activities; NVX-CoV2373 = 5 μg SARS-CoV-2 rS with 50 μg Matrix-M™ adjuvant; SARS-CoV-2 = severe acute respiratory syndrome coronavirus 2; SARS-CoV-2 rS = severe acute respiratory syndrome coronavirus 2 recombinant spike protein nanoparticle vaccine.

**Table S13. Overall Summary of Unsolicited Serious Adverse Events From Start of First Vaccination to Blinded Crossover Dose in All Adolescent Participants in Any Study Vaccine Group by Age Strata (Safety Analysis Set)**

| System Organ Class/ Preferred Term  (MedDRA, Version 24.0) | Participants 12 to < 18 Years | | Participants 12 to < 15 Years | | Participants 15 to < 18 Years | |
| --- | --- | --- | --- | --- | --- | --- |
|  | NVX-CoV2373  N = 1487 | Placebo  N = 745 | NVX-CoV2373  N = 998 | Placebo  N = 500 | NVX-CoV2373  N = 489 | Placebo  N = 245 |
|  | n (%) | n (%) | n (%) | n (%) | n (%) | n (%) |
| **Any system organ class** | **7 (0.5)** | **2 (0.3)** | **5 (0.5)** | **1 (0.2)** | **2 (0.4)** | **1 (0.4)** |
| **Injury, poisoning and procedural complications** | **3 (0.2)** | **0** | **2 (0.2)** | **0** | **1 (0.2)** | **0** |
| Intentional overdose | 2 (0.1) | 0 | 1 (0.1) | 0 | 1 (0.2) | 0 |
| Splenic rupture | 1 (< 0.1) | 0 | 1 (0.1) | 0 | 0 | 0 |
| **Infections and infestations** | **2 (0.1)** | **1 (0.1)** | **1 (0.1)** | **0** | **1 (0.2)** | **1 (0.4)** |
| Gastroenteritis norovirus | 1 (< 0.1) | 0 | 0 | 0 | 1 (0.2) | 0 |
| Localized infection | 1 (< 0.1) | 0 | 1 (0.1) | 0 | 0 | 0 |
| Peritonsillar abscess | 0 | 1 (0.1) | 0 | 0 | 0 | 1 (0.4) |
| **Nervous system disorders** | **2 (0.1)** | **0** | **1 (0.1)** | **0** | **1 (0.2)** | **0** |
| Juvenile myoclonic epilepsy | 1 (< 0.1) | 0 | 1 (0.1) | 0 | 0 | 0 |
| Seizure | 1 (< 0.1) | 0 | 0 | 0 | 1 (0.2) | 0 |
| **Psychiatric disorders** | **2 (0.1)** | **1 (0.1)** | **2 (0.2)** | **1 (0.2)** | **0** | **0** |
| Aggression | 1 (< 0.1) | 0 | 1 (0.1) | 0 | 0 | 0 |
| Suicide attempt | 1 (< 0.1) | 0 | 1 (0.1) | 0 | 0 | 0 |
| Mental status changes | 0 | 1 (0.1) | 0 | 1 (0.2) | 0 | 0 |

Abbreviations: MedDRA = Medical Dictionary for Regulatory Activities; NVX-CoV2373 = 5 μg SARS-CoV-2 rS with 50 μg Matrix-M™ adjuvant; SARS-CoV-2 = severe acute respiratory syndrome coronavirus 2; SARS-CoV-2 rS = severe acute respiratory syndrome coronavirus 2 recombinant spike protein nanoparticle vaccine.
